# Parent-of-Origin inference and its role in the genetic architecture of complex traits: evidence from ∼265,000 individuals

**DOI:** 10.1101/2024.12.03.24318392

**Authors:** Robin J. Hofmeister, Théo Cavinato, Roya Karimi, Adriaan van der Graaf, Fanny-Dhelia Pajuste, Jaanika Kronberg, Nele Taba, Estonian Biobank research team, Reedik Mägi, Marc Vaudel, Simone Rubinacci, Stefan Johansson, Lili Milani, Olivier Delaneau, Zoltán Kutalik

## Abstract

Parent-of-origin effects (POEs) occur when the impact of a genetic variant depends on its parental origin. Traditionally linked to genomic imprinting, these effects are believed to have evolved from parental conflict over resource allocation to offspring, which results in opposing parental genetic influences. Despite their potential importance, POEs remain heavily understudied in complex traits, largely due to the lack of parental genomes.

Here, we present a multi-step approach to infer the parent-of-origin of alleles without parental genomes, leveraging inter-chromosomal phasing, mitochondrial and chromosome X data, and sibling-based crossover inference. Applied to the UK Biobank (discovery cohort) and Estonian Biobank (replication cohort), this scalable approach enabled parent-of-origin inference for up to 221,062 individuals, representing the largest dataset of its kind.

GWAS scans in the UK Biobank for more than 60 complex traits and over 2,400 protein levels contrasting maternal and paternal effects identified over 30 novel POEs and confirmed more than 50% of testable known associations. Notably, approximately half of our POEs exhibited a bi-polar pattern, where maternal and paternal alleles exert conflicting effects. These effects were particularly prevalent for traits related to growth (e.g., IGF-1, height, fat-free mass) and metabolism (e.g., type 2 diabetes, triglycerides, glucose). Replication in the Estonian Biobank and in 45,402 offspring from the Norwegian Mother, Father and Child Cohort Study validated over 75% of testable associations.

Overall, our findings shed new light on the influence of POEs on diverse complex traits and align with the parental conflict hypothesis, providing compelling evidence for this understudied evolutionary phenomenon.

## Introduction

Genome-wide association studies (GWAS) have traditionally focused on identifying additive genetic effects, where the phenotypic impact depends on the number of copies of a given allele. This approach assumes that alleles inherited from the mother and father contribute equally to the examined trait. However, some sequence variants can have distinct phenotypic effects depending on whether they are maternally or paternally inherited, a phenomenon known as parent-of-origin effects (POEs). Traditionally, POEs have been linked to genomic imprinting, where only one parental gene is expressed depending on its parental origin. This selective expression is believed to have evolved under the parental conflict hypothesis, which suggests opposing evolutionary pressures on maternal and paternal alleles over resource allocation to offspring: paternally inherited alleles are believed to enhance offspring growth even at the cost of maternal resources, favoring paternal genetic fitness, while maternally inherited alleles conserve resources for future reproduction^1^. This dynamic can result in opposite parental effects at imprinted loci, particularly for traits related to growth, metabolism, and energy storage. However, since most studies have identified only isolated instances of POEs without a systematic evaluation of their abundance, there is only scarce examples to support the conflict hypothesis across a wide array of traits.

While the conflict hypothesis provides a compelling framework for understanding genomic imprinting, the observation of POEs in non-imprinted genes raises the possibility of more complex inheritance patterns influencing certain complex traits^2,3^. Alternatively, some of these effects might also reflect environmental or parental rearing influences rather than strictly genetic mechanisms. This broader perspective underscores the importance of extending investigations beyond classical imprinted regions to open avenues to explore new mechanisms leading to POEs.

Studying POEs at a genome-wide scale requires large cohorts with parent-of-origin (PofO) information. Traditional approaches rely on extensive collections of parent-offspring trios or duos or detailed genealogical data to identify relatives as surrogate parents^4,5^. However, large-scale biobanks often lack both substantial parental genomic data and comprehensive genealogies^6^. Our previous work introduced a method leveraging chromosome X sharing to identify maternal relatives without genealogical information^3^, which opened new research opportunities and significantly increased the sample size. However, this method is limited to male participants, prohibiting sex-specific POE detection, constraining sample size and hence reducing statistical power.

To overcome these challenges, we developed a novel multi-step approach for inferring the PofO of sequence variants in many additional pedigree situations. First, we partition the genome into maternal and paternal haplotypes through statistical inter-chromosomal phasing, using inferred surrogate parents. To determine the PofO of these haplotypes, we integrate several predictors: beyond leveraging chromosome X data for males, we also use mitochondrial DNA whole-genome sequencing (WGS). In addition, we infer crossover positions in sibling pairs and assign their PofO using sex-specific recombination maps^7,8^ (Figure 1). Using these predictors, our approach probabilistically determines the PofO of variants, maximizing sample size while minimizing misclassification errors inherent to strict parent assignments used in our prior study^3^.

**Fig. 1.**
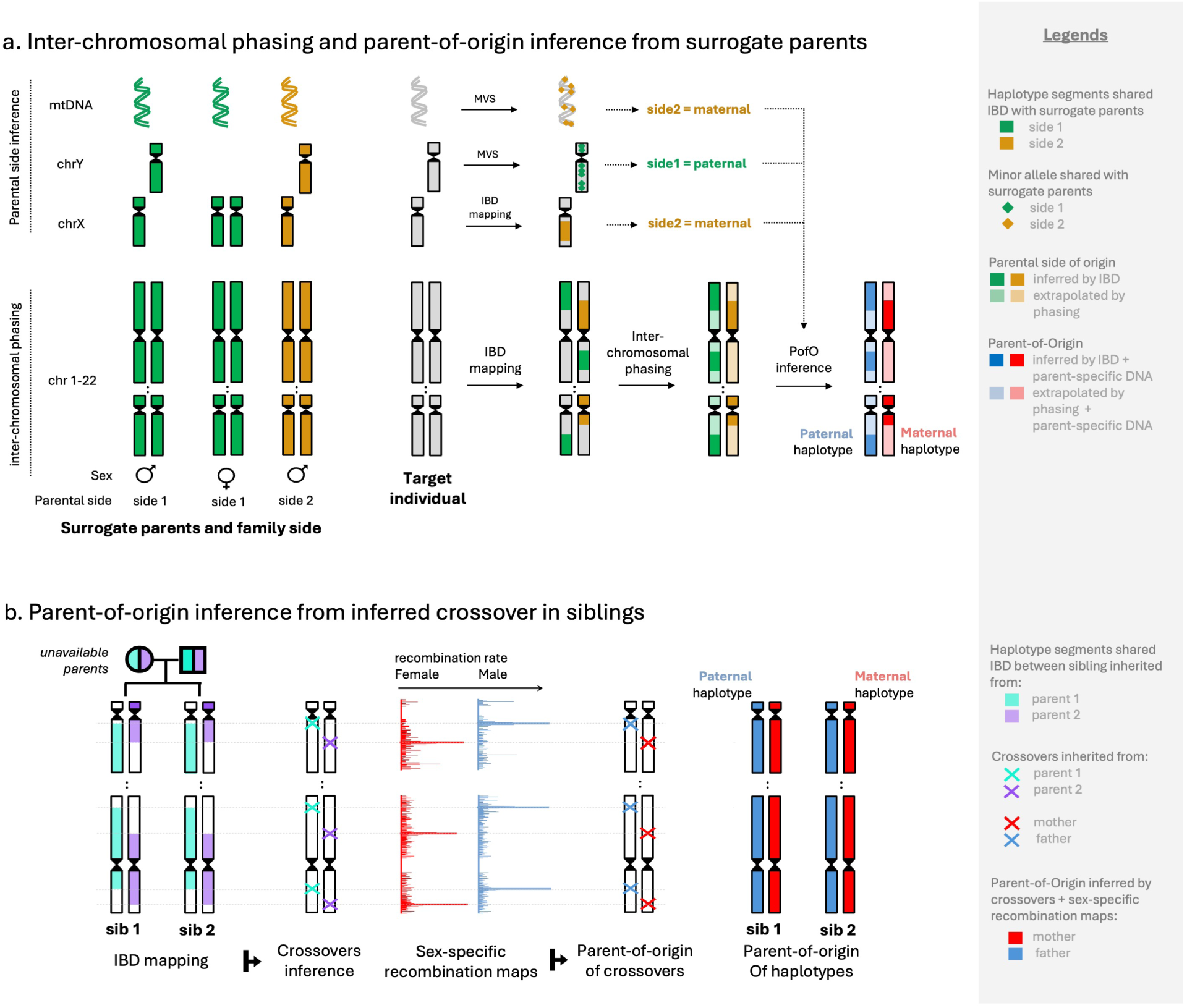
Inter-chromosomal phasing and parent-of-origin methods overview. **a**) Inter-chromosomal phasing and parent-of-origin inference. We initially clustered relatives (2*^nd^*, 3*^rd^* and 4*^th^* degree) into surrogate parent groups, segregating the relatives into “one family side” (green) *vs* “the other family side” (orange). We then combined two steps to infer the PofO of a focal individual: on the one hand, we leveraged IBD sharing with surrogate parent groups to perform inter-chromosomal phasing. For instance, green IBD tracks were forced to align on the first haplotype, simultaneously correcting for intra-chromosomal phasing errors; on the other hand, we examined genetic similarities on chromosome X, Y and mtDNA between the focal individual and its surrogate parents, to assign surrogate parent groups to a parental side. Finally, we deduce the PofO of inter-chromosomally phased haplotypes from the parental side of the surrogate parent in IBD (see Methods for details). **b**) To determine the PofO of siblings, we first inferred crossover events using IBD haplotypes. We then overlapped inferred crossovers with sex-specific genetic map to derive the likelihood of a crossover originating from the mother or the father. This subsequently allowed us to deduce the PofO of the haplotype carrying the crossover.

We initially applied this method to the UK Biobank, successfully inferring the PofO for 123,716 individuals at over 7.6 million variant sites — a five-fold increase in sample size compared to our previous work^3^, which served as the discovery cohort. For replication, we implemented our approach in the Estonian Biobank, inferring PofO information for 97,346 individuals, and furthermore we used 45,402 offspring (with parental genotypes) from the Norwegian Mother, Father and Child Cohort Study. This combined dataset of 266,464 individuals enabled a systematic investigation of POEs across a broad range of human traits, including anthropometric characteristics, blood biomarkers, diseases, and protein levels. Our analysis identified over 30 novel POEs, particularly on traits tied to resource allocation, such as growth-related and metabolic traits, as well as protein levels. Many of these effects aligned with the parental conflict hypothesis, primarily through bi-polar effects, where maternal and paternal alleles exert opposing influences. These findings highlight the genetic and evolutionary significance of POEs in shaping complex traits and diseases, offering new insights into their broader biological roles.

## Results

We developed a comprehensive, multi-step strategy to infer the PofO of sequence variants in large-scale biobanks. A brief summary of the methods is outlined below, followed by a detailed analysis of POEs.

### Summary of the methods

#### Surrogate parent-based inter-chromosomal phasing

We used kinship estimates, age and sex to identify parent-offspring trios, duos and siblings. For individuals with more distant relatives, we utilized a clustering approach to segregate relatives by parental sides^3^, thereby classifying them into “one family side” versus “the other family side”. This could be done for a total of 274,525 UK Biobank participants (Supplementary Figure 1). In subsequent sections, we refer to those family groups as “surrogate parents”. Within this subset, 2,141 individuals also had directly available parental genomes. This group served as a “validation cohort” for assessing the accuracy of our method by comparing surrogate parent-based inferences against true parental genotypes (Supplementary Figure 2). We then used identity-by-descent (IBD) information from surrogate parents to conduct inter-chromosomal phasing, effectively identifying sets of variants inherited together from the same parent across different chromosomes (Figure 1A). This phasing method was applied to 274,525 UK Biobank participants, achieving an average inter-chromosomal phasing accuracy of 99% (Supplementary Figure 3). In subsequent analyses, we refer to this dataset as “inter-chromosomally phased”.

#### Parent-of-origin determination for 123,716 UK Biobank individuals

We leveraged our inter-chromosomally phased data to infer the PofO of each complete parental haplotype set simultaneously. We used two key approaches depending on the available surrogate parents.

First, for second- to fourth-degree relatives, the PofO was determined by identifying whether the surrogate parent was on the maternal or paternal side. Once the PofO of a single haplotype segment was established through its IBD status with the used surrogate parents, this assignment could be confidently extended to the entire parental haplotype set. To determine whether the used surrogate parents are on the maternal or on the paternal side, we leveraged chromosome X genotype data and mtDNA whole-genome sequence data, while also demonstrating the potential value of including chromosome Y whole-genome sequences (Figure 1A, see Methods). Chromosome X is maternally inherited in males, and mtDNA is maternally inherited in both sexes, allowing us to identify maternal relationships (Supplementary Figures 4-8). Although chromosome Y data can reliably identify paternal relationships (Supplementary Figure 9), we found this approach to be less cost-effective and therefore chose not to implement it broadly. By analyzing IBD sharing with surrogate parents on these specialized chromosomes, we were able to determine whether the surrogate parent (and, consequently, all IBD segments shared on other chromosomes) belonged to the maternal or paternal side.

For siblings, who cannot be assigned exclusively to a single parental side since they inherit genomes from both parents, we applied a different strategy. Specifically, we inferred crossover positions in siblings and probabilistically assigned them a PofO using sex-specific recombination maps^7,8^ (Figure 1B). For this approach, we prioritized inter-chromosomally phased data that improved accuracy by aggregating crossovers events across chromosomes (see methods, Supplementary Figures 10-12).

We combined these different kinds of PofO predictors (see Methods) with inference made from available parental genomes to obtain PofO estimates for a set of 286,666 individuals. Notably, over 40% of these individuals (N=123,716) had an estimated PofO probability between 0.99 and 1 (Figure 13A,B). For 31,008 individuals, multiple predictors were available, and these consistently indicated the same PofO in over 94% of cases (N=29,154 individuals), further reinforcing the reliability of these assignments. Using the validation cohort (where the PofO of the alleles is known) we estimated the accuracy of our inference to be 97.94% at heterozygous sites (Figure 13C). Notably, the majority of individuals (*>* 80%) showed accuracy greater than 99%. This high level of accuracy demonstrates the reliability of our method for determining the PofO of alleles across the genome.

Next, we used the resulting PofO of alleles to perform PofO specific association scans on 59 selected complex traits (Supplementary Table 1) and 2,414 proteins. We selected complex traits primarily from biomarkers and morphological measures to assess POEs across a diverse range of phenotypes while including traits associated with resource allocation, such as growth-related traits (e.g. height and fat-free mass), metabolic traits (e.g., glucose, lipids, and basal metabolic rate), and traits related to energy storage (e.g. regional fat percentages and BMI), allowing us to test the conflict hypothesis. Our analysis employed three main approaches: (i) we first examined imprinted regions with established imprinting potential to detect loci most likely to exhibit POEs, (ii) we then focused on regions exhibiting additive associations with the trait (referred to as “additively associated regions” in subsequent sections), and (iii) we finally scanned for POEs genome-wide to identify novel POEs without prior assumptions.

### Replicating known POE associations

We first replicated POEs previously reported in the literature^3,4,5,9,10,11,12,13^, both to validate known POEs and to validate our PofO assignment. No consensus method is currently used to identify POEs and many studies simply claim POEs if the effect is significant (at an arbitrary threshold) when the allele came from one parent, but not significantly associated when it came from the other parent. Therefore, we first pre-filtered claimed POEs by rigorously testing for a differential paternal *vs* maternal alleles effect and kept only published hits where the differential test P-value *P_D_* was lower than 10^−3^, to ensure more robust evidence for POE.

We identified 23 known POEs meeting this criterion, of which 22 could be tested for replication in our cohort (same variant-trait pair). We successfully replicated 12 of them (54.5%) at *P_D_ <* 0.05*/*22. Interestingly, of the 8 POEs reported for birth weight, none were replicated in our cohort. For the other traits, we successfully replicated *>* 85% (12*/*14) POEs, with the same parental effect and the same direction of effect (Supplementary Table 2).

### Parent-of-origin association within imprinted regions

To identify POEs within imprinted regions, we restricted our POE analysis to variants located within a 500kb window of known imprinted regions, which are more likely to harbor POEs (Supplementary Table 3)^5^. We applied this approach to the 59 selected traits (Supplementary Table 1) and identified a total of 27 POEs with significant (0.05/16,574, see Methods) POE difference P-value *P_D_* (Table 1).

**Table 1.** Significant Parent-of-origin identified in this study. chromosome; POS: genetic position (hg19); SNP ID: variant rs id; A0: reference allele; A1: assessed allele; EQ: A1 allele frequency. BETA, SE and P denote effect sizes, standard errors and P-values; PAT, MAT, DIFF DD denote paternal, maternal, differential and additive tests. TRAIT: phenotype name; *: POE indentified the imprinted region-focused approach that are also robust when correcting for the number of phenotype (see Methods); (c) denote POE identified in a conditional analysis. For these, the conditional estimates are ed. For these, reported coefficients are those from the conditional analysis; SCAN indicate the approach (I printed region-focus, A for additively associated region-focused, G for genome-wide scan); bold ID: bi-polar

| CHR | POS | SNP ID | A0 | A1 | A1FREQ | BETA PAT | SE PAT | P PAT | BETA MAT | SE MAT | P MAT | BETA DIFF | SE DIFF | P DIFF | BETA ADD | SE ADD | P ADD | TRAIT | STATUS | Locus | SCAN |
| --- | --- | --- | --- | --- | --- | --- | --- | --- | --- | --- | --- | --- | --- | --- | --- | --- | --- | --- | --- | --- | --- |
| 11 | 1914139 | rs576603 | C | T | 0.37 | -0.027 | 0.004 | 8.53E-11 | 0.005 | 0.004 | 2.03E-01 | -0.033 | 0.006 | 3.86E-08 | -0.011 | 0.003 | 2.12E-04 | Standing height* (c) | novel | H19,IGF2 | I |
| 11 | 2040272 | **rs77708343** | A | G | 0.039 | -0.055 | 0.011 | 2.21E-07 | 0.034 | 0.010 | 1.07E-03 | -0.088 | 0.015 | 2.18E-09 | -0.010 | 0.007 | 1.92E-01 | Standing height* | known | H19, IGF2 | I |
| 11 | 2813322 | rs143840904 | C | T | 0.019 | -0.023 | 0.015 | 1.37E-01 | -0.183 | 0.016 | 9.13E-32 | 0.152 | 0.021 | 5.21E-13 | -0.100 | 0.011 | 2.56E-20 | Standing height* | known | KCNQ1 | I/A/G |
| 11 | 2040272 | **rs77708343** | A | G | 0.040 | -0.048 | 0.011 | 1.25E-05 | 0.035 | 0.011 | 1.33E-03 | -0.083 | 0.015 | 7.53E-08 | -0.006 | 0.008 | 4.34E-01 | Basal metabolic rate | novel | H19, IGF2 | I |
| 11 | 2040272 | **rs77708343** | A | G | 0.040 | -0.052 | 0.011 | 1.28E-06 | 0.033 | 0.011 | 2.06E-03 | -0.085 | 0.015 | 1.80E-08 | -0.009 | 0.008 | 2.29E-01 | Leg fat-free mass* | novel | H19, IGF2 | I |
| 11 | 2040272 | **rs77708343** | A | G | 0.040 | -0.044 | 0.010 | 3.08E-05 | 0.033 | 0.010 | 1.23E-03 | -0.077 | 0.015 | 1.63E-07 | -0.005 | 0.007 | 5.32E-01 | Whole body water mass | novel | H19, IGF2 | I |
| 11 | 2041348 | **rs78507815** | G | T | 0.053 | -0.031 | 0.009 | 7.03E-04 | 0.033 | 0.009 | 1.86E-04 | -0.065 | 0.013 | 3.61E-07 | 0.002 | 0.006 | 7.68E-01 | Trunk fat-free mass | novel | H19, IGF2 | I |
| 11 | 1920285 | rs4264135 | A | G | 0.293 | -0.033 | 0.006 | 1.83E-07 | 0.014 | 0.006 | 3.31E-02 | -0.046 | 0.009 | 2.22E-07 | -0.010 | 0.005 | 2.90E-02 | Urate | novel | H19, IGF2 | I |
| 11 | 1998031 | **rs170102** | G | A | 0.346 | -0.041 | 0.006 | 5.96E-11 | 0.026 | 0.006 | 3.19E-05 | -0.067 | 0.009 | 5.98E-14 | -0.007 | 0.004 | 9.47E-02 | Cystatin C* | novel | H19, IGF2 | I/G |
| 11 | 2003944 | **rs217215** | T | G | 0.656 | 0.030 | 0.006 | 2.53E-07 | -0.024 | 0.006 | 2.68E-05 | 0.054 | 0.008 | 4.94E-11 | 0.003 | 0.004 | 5.04E-01 | Creatinine* | novel | H19, IGF2 | I/G |
| 11 | 1703564 | **rs4417225** | C | T | 0.426 | 0.029 | 0.008 | 9.26E-05 | -0.028 | 0.008 | 2.04E-04 | 0.057 | 0.011 | 7.40E-08 | 0.001 | 0.005 | 8.85E-01 | Glucose | novel (tags known T2D) | H19, IGF2 | I |
| 11 | 1702929 | **rs10838787** | G | A | 0.425 | 0.136 | 0.027 | 3.88E-07 | -0.091 | 0.027 | 7.55E-04 | 0.228 | 0.038 | 1.87E-09 | 0.023 | 0.019 | 2.26E-01 | Type 2 diabetes* | known | H19, IGF2 | I |
| 11 | 1702929 | **rs10838787** | G | A | 0.425 | 0.050 | 0.007 | 2.34E-14 | -0.028 | 0.007 | 1.80E-05 | 0.078 | 0.009 | 2.77E-17 | 0.011 | 0.005 | 1.76E-02 | Glycated haemoglobin* | novel (tags known T2D) | H19, IGF2 | I/G |
| 11 | 2858295 | rs2299620 | C | T | 0.036 | -0.011 | 0.018 | 5.22E-01 | -0.129 | 0.017 | 1.50E-13 | 0.116 | 0.024 | 2.23E-06 | -0.072 | 0.013 | 9.05E-09 | Glycated haemoglobin | novel (tags known T2D) | KCNQ1 | I |
| 7 | 130009312 | **rs10239342** | T | G | 0.309 | 0.022 | 0.007 | 1.36E-03 | -0.024 | 0.007 | 4.15E-04 | 0.045 | 0.009 | 1.87E-06 | -0.001 | 0.005 | 8.17E-01 | SHBG | novel | KLF14, MEST, CPA4 | I |
| 7 | 130016470 | **rs62471721** | G | A | 0.351 | -0.023 | 0.007 | 9.70E-04 | 0.033 | 0.007 | 1.57E-06 | -0.057 | 0.010 | 6.51E-09 | 0.005 | 0.005 | 2.85E-01 | Triglycerides* | novel | KLF14, MEST, CPA4 | I |
| 7 | 130017940 | **rs4731690** | A | G | 0.503 | 0.019 | 0.006 | 1.97E-03 | -0.022 | 0.006 | 2.76E-04 | 0.041 | 0.009 | 1.92E-06 | -0.002 | 0.004 | 7.01E-01 | HDL cholesterol | novel | MEST, CPA4, | I |
| 7 | 130463192 | rs6467315 | C | G | 0.509 | 0.001 | 0.006 | 9.02E-01 | -0.042 | 0.006 | 5.81E-11 | 0.043 | 0.009 | 2.20E-06 | -0.021 | 0.005 | 5.72E-06 | Hip circumference | novel (tags known T2D) | KLF14, CPA4 | I |
| 7 | 130400698 | rs3847104 | G | A | 0.877796 | -0.016 | 0.010 | 1.01E-01 | 0.056 | 0.010 | 1.94E-08 | -0.072 | 0.014 | 3.81E-07 | 0.020 | 0.007 | 4.62E-03 | Hip circumference (c) | novel | KFL14, CPA4 | I |
| 20 | 57216538 | **rs80116540** | A | G | 0.075 | 0.035 | 0.010 | 4.38E-04 | -0.043 | 0.010 | 2.04E-05 | 0.078 | 0.014 | 3.39E-08 | -0.004 | 0.007 | 6.09E-01 | Arm fat percentage* | novel | GNAS | I |
| 20 | 57216538 | **rs80116540** | A | G | 0.075 | 0.037 | 0.010 | 2.65E-04 | -0.037 | 0.010 | 2.27E-04 | 0.074 | 0.014 | 2.06E-07 | 0.000 | 0.007 | 9.90E-01 | Body fat percentage | novel | GNAS | I |
| 20 | 57226079 | **rs6026426** | C | G | 0.070 | 0.045 | 0.012 | 3.08E-04 | -0.040 | 0.012 | 1.37E-03 | 0.085 | 0.018 | 1.29E-06 | 0.003 | 0.009 | 7.64E-01 | Trunk fat percentage | novel | GNAS | I |
| 20 | 57484934 | **rs3730173** | C | T | 0.052 | 0.036 | 0.010 | 3.18E-04 | -0.035 | 0.010 | 3.95E-04 | 0.071 | 0.014 | 5.32E-07 | 0.000 | 0.007 | 9.95E-01 | Leg fat percentage | novel | GNAS | I |
| 6 | 144274210 | rs12528876 | T | C | 0.171 | -0.036 | 0.009 | 2.58E-05 | 0.021 | 0.009 | 1.60E-02 | -0.057 | 0.012 | 2.96E-06 | -0.008 | 0.006 | 1.96E-01 | IGF-1 | novel | PLAGL1 | I |
| 16 | 3412861 | rs7188903 | A | G | 0.703 | -0.012 | 0.007 | 1.03E-01 | 0.041 | 0.007 | 5.35E-08 | -0.053 | 0.011 | 6.14E-07 | 0.014 | 0.005 | 7.24E-03 | IGF-1 | novel | ZNF597, NAA60 | I |
| 15 | 25983333 | **rs146982369** | C | G | 0.014 | 0.101 | 0.032 | 1.88E-03 | -0.106 | 0.032 | 9.99E-04 | 0.208 | 0.044 | 2.59E-06 | -0.003 | 0.023 | 8.82E-01 | Total protein | novel | ATP10A | I |
| 14 | 101185187 | rs59228823 | G | C | 0.245 | -0.010 | 0.008 | 2.04E-01 | -0.092 | 0.008 | 3.87E-34 | 0.082 | 0.011 | 1.92E-14 | -0.051 | 0.005 | 2.36E-21 | Platelet count* | known | MEG3 | I/A/G |
| 3 | 169482335 | rs2293607 | T | C | 0.245 | -0.122 | 0.008 | 2.26E-55 | -0.068 | 0.008 | 5.36E-18 | -0.055 | 0.011 | 5.70E-07 | -0.095 | 0.006 | 3.34E-66 | TS ratio | novel | TERC | A |
| 4 | 164012901 | rs11100479 | C | T | 0.778 | 0.079 | 0.008 | 1.82E-22 | 0.022 | 0.008 | 6.61E-03 | 0.057 | 0.012 | 6.25E-07 | 0.050 | 0.006 | 1.46E-18 | TS ratio | novel | NAF1 | A |
| 5 | 1285974 | rs7705526 | C | A | 0.323 | 0.038 | 0.007 | 2.09E-07 | 0.106 | 0.007 | 1.27E-47 | -0.068 | 0.010 | 5.82E-11 | 0.072 | 0.005 | 5.78E-44 | TS ratio | known | TERT | A/G |

A systematic evaluation of the lead POE variants across traits revealed two key patterns. First, traits related to growth and metabolism were particularly over-represented among pleiotropic loci, notably on chromosomes 7, 11, and 20 (Figure 2A), consistent with the expected roles of imprinted genes under the conflict theory. Second, the majority of these POEs exhibited opposite parental effects on the examined trait, i.e the trait is increased by the allele inherited from one parent, but decreased when the allele is inherited from the other parent (Figure 2B). These opposite parental effects align with a pattern of bi-polar dominance^14,15^, emphasizing the distinct and opposing contributions of each parent’s allele to the trait. Notably, such effects are often missed by traditional additive GWAS, where these parental effects cancel out at heterozygotes (Supplementary Figure 15).

**Fig. 2.**
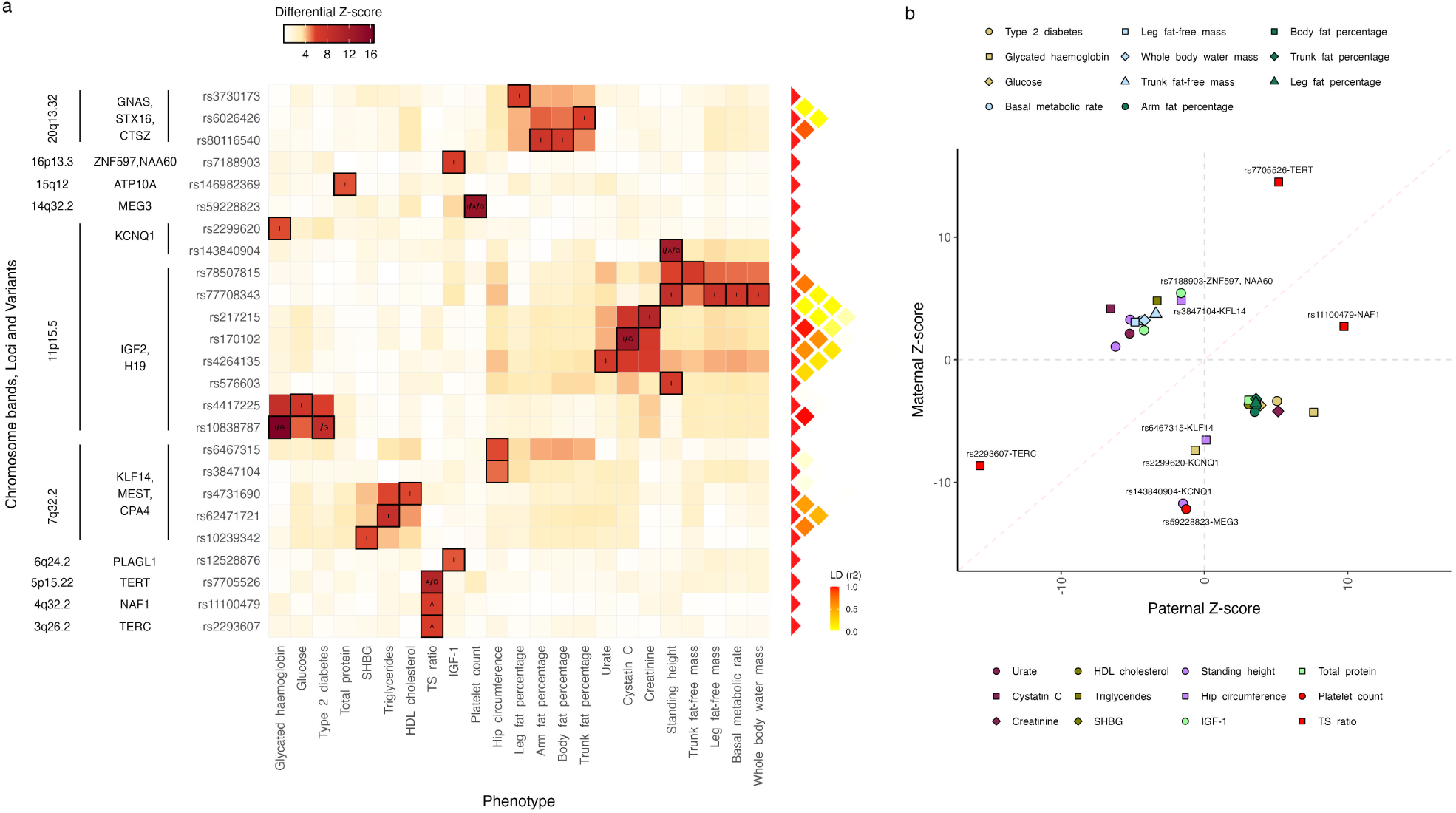
Significant parent-of-origin effects. **a**) Heatmap summarizing the differential Z-scores for all significant POEs identified in this study. Rows correspond to genetic variants ordered by genetic position and annotated with the genes and chromosome bands, while columns represent phenotypes. The color intensity represents the magnitude of the differential Z-scores, with darker shades indicating stronger differential effects. Cells with black rectangles indicate significant POEs identified in this SNP-trait pair and reported in Table 1. Black annotations within cells specify the type of approach that led to the identification of the POE: “I” for imprinted region-focused analysis, “A” for additively associated region-focused analysis, “G” for genome-wide analysis. The LD heatmap on the right shows linkage disequilibrium (LD; *r*^2^) between the variants. **b**) Scatter plot illustrating the parental Z-scores for all significant POEs identified in this study and reported in Table 1. Each point represents a significant POE, colored and shaped by phenotype. The dashed red line represents the line of equality, and dashed gray lines represent zero values for paternal and maternal Z-scores, respectively. Labeled points correspond to POE classified as paternal or maternal effects. Unlabeled points correspond to bi-polar POEs.

In the next sections, we described the POE associations identified using our imprinted region-focused approach across several regions and traits, which exemplifies the diversity and pleiotropy observed in our POE analysis.

#### A complex pattern of bi-polar and maternal dominant effect on standing height at 11p15.5

We detected three independent loci associated with standing height in a parent-of-origin dependent fashion at the *H19* /*IGF2* imprinted region (Table 1, Figure 2, Figure 3). Two of these, rs143840904 and rs77708343, are in high LD (*r*^2^ *>* 0.6) with variants that were associated with height in previous POE studies^9,12,13^ (see replication section, Figure 3A-F). At the same locus, the conditional analysis allowed us to discover an independent and novel POE of rs576603, located more than 120kb away and not in LD with both previously reported lead SNPs (*r*^2^ *<* 0.003 with rs77708343 and rs147239461, respectively). The paternal T allele of this variant is associated with decreased height, while the maternal one had no effect (*β̂_P_* = −0.027, *β̂_M_* = 0.005, *P_P_* = 8.53 × 10^−11^, *P_M_* = 0.2, *P_D_* = 3.86 × 10^−8^, Figure 3G, Supplementary Figure 16). The variant is a splicing QTL (sQTL) for the maternally expressed *H19* gene in *>*25 GTEx tissues and also acts as an eQTL for the paternally expressed gene *IGF2* in tibial artery^16^. Further conditional analyses showed that all three variants were independently associated with standing height (*P_D_c__* = 3.86 × 10^−8^, 3.88 × 10^−10^ and 5.8×10^−13^ for rs576603, rs77708343 and rs143840904, respectively).

**Fig. 3.**
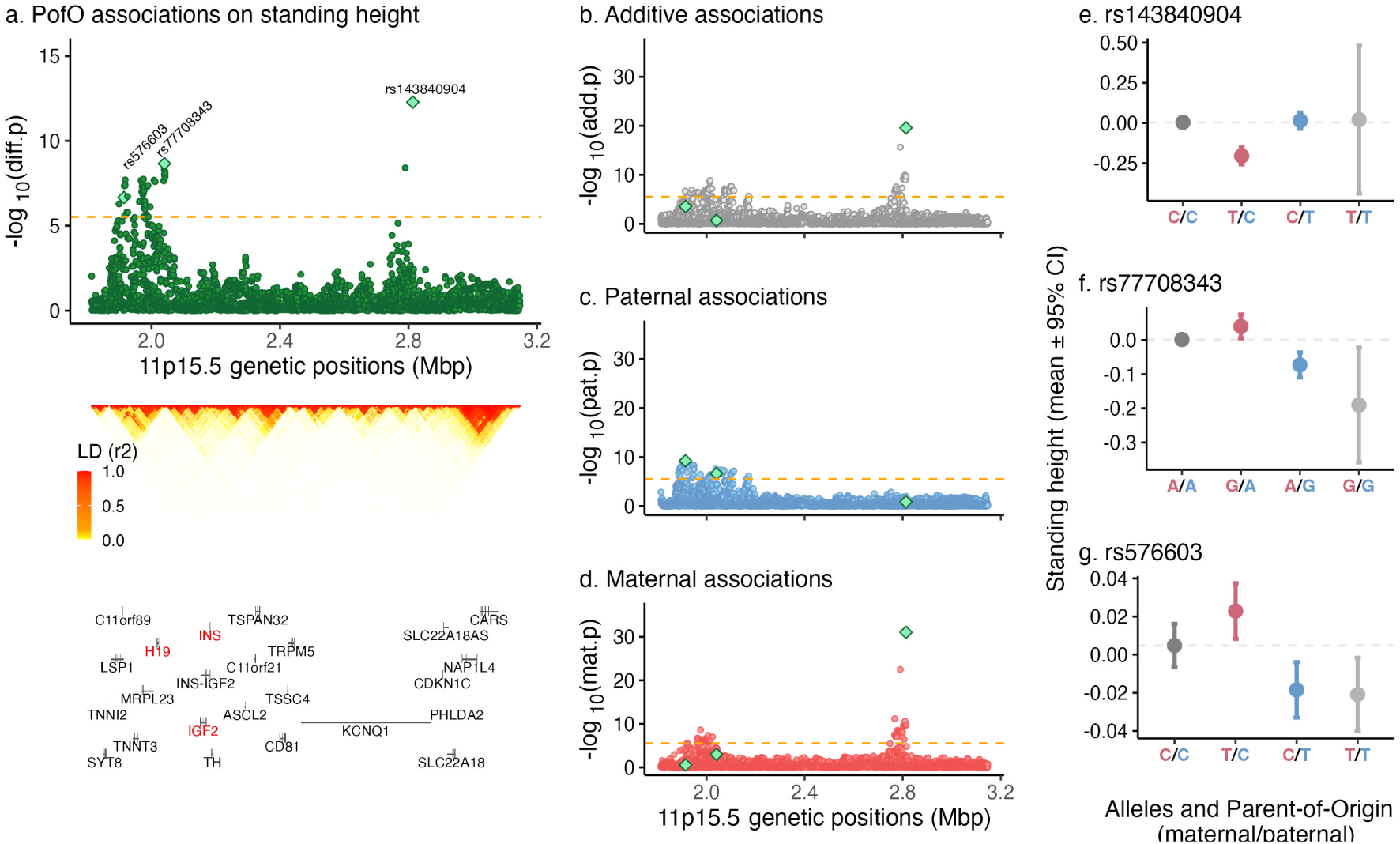
Parent-of-Origin associations on standing height. **a**) Differential GWAS (top) shows the association strength (*−log*_10_(*p − value*), y-axis) against the genomic position (x-axis). Each point represents a genetic variant. Diamonds indicate variants with independent POE on standing height found in this study (see Supplementary Figure 16 for the conditional analysis). Orange dashed line represents the significance threshold used in this study when focusing on imprinted regions (3.1 *×* 10^−06^). Linkage Disequilibrium (LD) pattern (middle) ranges from LD=0 (white) to LD=1 (red). Gene positions (bottom) along the 11p15.5 imprinted region (x-axis). Horizontal lines show the gene start and end positions. Vertical lines show the exons start and end positions. Gene names are shown below the corresponding gene coordinates. **b-d**) Additive (b, grey), Paternal (c, blue) and Maternal (d, red) associations on standing height shown as association strength (*−log*_10_(*p − value*), y-axis) against the genomic position (x-axis). Green diamonds indicate POEs selected in the differential scan. Orange dashed lines represent the significance threshold used in this study when focusing on imprinted regions (3.1 *×* 10^−06^). **e-g**) Genotypes and PofO of alleles (x-axis) effects on standing height (y-axis). Red and blue alleles indicate maternal and paternal alleles, respectively. Dots show mean values, error bars show 95% confidence intervals. Grey dashed lines represent the mean value of individuals carrying no alternative allele (i.e., genotype 0). Dots and bars colors indicate maternal heterozygotes (red), paternal heterozygotes (blue), and homozygotes (dark and light grey).

To understand how these loci can differentially impact various growth parameters, we evaluated these variants for POEs on sitting height. For rs143840904 and rs77708343, the associations showed similar (and significant) trends for sitting height (see Supplementary Table 4). Interestingly, while the significance and effect sizes (in SD) of rs143840904 decreased for sitting height, the association of rs77708343 remained relatively stable, indicating that it may contribute equally to trunk size as it does to overall height. On the other hand, rs576603 did not associate with sitting height in a similar fashion, suggesting that this variant likely contributes to femoral length, rather than to trunk size.

#### Novel bi-polar dominant effects on metabolic traits at 11p15.5

At the same locus, we also uncovered novel bi-polar POEs on metabolic rate and body composition traits (Table 1, Figure 2). Specifically, the paternal G allele of the standing height-associated SNP rs77708343 (see above) was also associated with a decrease in metabolic rate, while the maternal G allele showed an increase (*β̂_P_* = −0.048, *β̂_M_* = 0.035, *P_P_* = 1.2 × 10, *P_M_*= 1.3 × 10, *P_D_*= 7.5 × 10). Given that basal metabolic rate is estimated from height and weight, we further examined the effects of rs77708343 on weight, and found the paternal G allele associated with decreased, and the maternal G allele associated with increased weight (*β̂_P_* = −0.064, *β̂_M_* = 0.031, *P_P_* = 1.3 × 10^−5^, *P_M_* = 3.1 × 10^−2^, *P_D_* = 3.8 × 10).

Due to the correlation between height and weight, we further explored associations of rs77708343 with additional morphological traits, including waist and hip circumference, as well as fat, fat-free and water mass measurements for legs, arms, trunk and whole body. The analysis revealed that rs77708343 influenced waist circumference, fat-free mass and whole body water mass, while no significant effect was observed on hip circumference or fat percentage. Specifically, the paternal G allele was associated with decreased and the maternal G allele with increased trait levels (*β̂_P_* = −0.056, −0.052, −0.032, −0.038 and −0.043, *β̂_M_* = 0.039, 0.032, 0.030, 0.034 and 0.033, *P_P_* = 1.0 × 10^−3^, 1.2 × 10^−6^, 1.8 × 10^−3^, 1.9 × 10^−4^ and 1.0 × 10^−5^, *P_M_* = 1.7 × 10^−2^, 2.0 × 10^−3^, 3.1 × 10^−3^, 8.0 × 10^−4^ and 1.2 × 10^−3^, *P_D_* = 7.0 × 10^−5^, 1.8 × 10^−8^, 1.6 × 10^−5^, 5.5 × 10^−7^ and 1.6 × 10^−7^ for waist circumference, leg, arm, and trunk fat-free mass, and whole-body water mass, respectively). Notably, leg and trunk fat-free mass, and whole body water mass also met our significance threshold specific to imprinted regions at this locus (Table 1).

#### Distinct bi-polar dominant and maternal effects on blood glucose biomarkers and type 2 diabetes at the 11p15.5 region

We identified novel associations with type 2 diabetes (T2D), HbA1c and glucose levels at two variants in high LD (*r*^2^ *>* 0.99), rs10838787 and rs4417225 (*P_D_* = 1.8×10^−9^, 2.7×10^−17^ and 7.4 × 10^-8^ , respectively, Table 1, Figure 2). Specifically, we found the paternal A allele of rs10838787 to be associated with increased T2D prevalence, while the maternal A allele appears to be protective (*β̂_P_* = 0.136, *β̂_M_* = −0.09, *P_P_* = 3.8 × 10^−7^, *P_M_* = 7.4*e* × 10^−4^, *P_D_* = 1.8 × 10^-9^ ). We also found bi-polar dominant effects on HbA1c and glucose, with paternal alleles associated with increased, and maternal alleles associated with decreased trait levels (*β̂_P_* = 0.05 and 0.029, *β̂_M_* = −0.028 and −0.028, *P_P_* = 2.3×10^−14^ and 9.2×10^−5^, *P_M_* = 1.8 × 10^-5^ and 2.0 × 10^-4^ for HbA1c and glucose, respectively, Figures 4A). Although these variants are novel, they are in high LD (*r*^2^ *>* 0.9) with a variant previously associated with T2D in a parent-of-origin manner^5^.

**Fig. 4.**
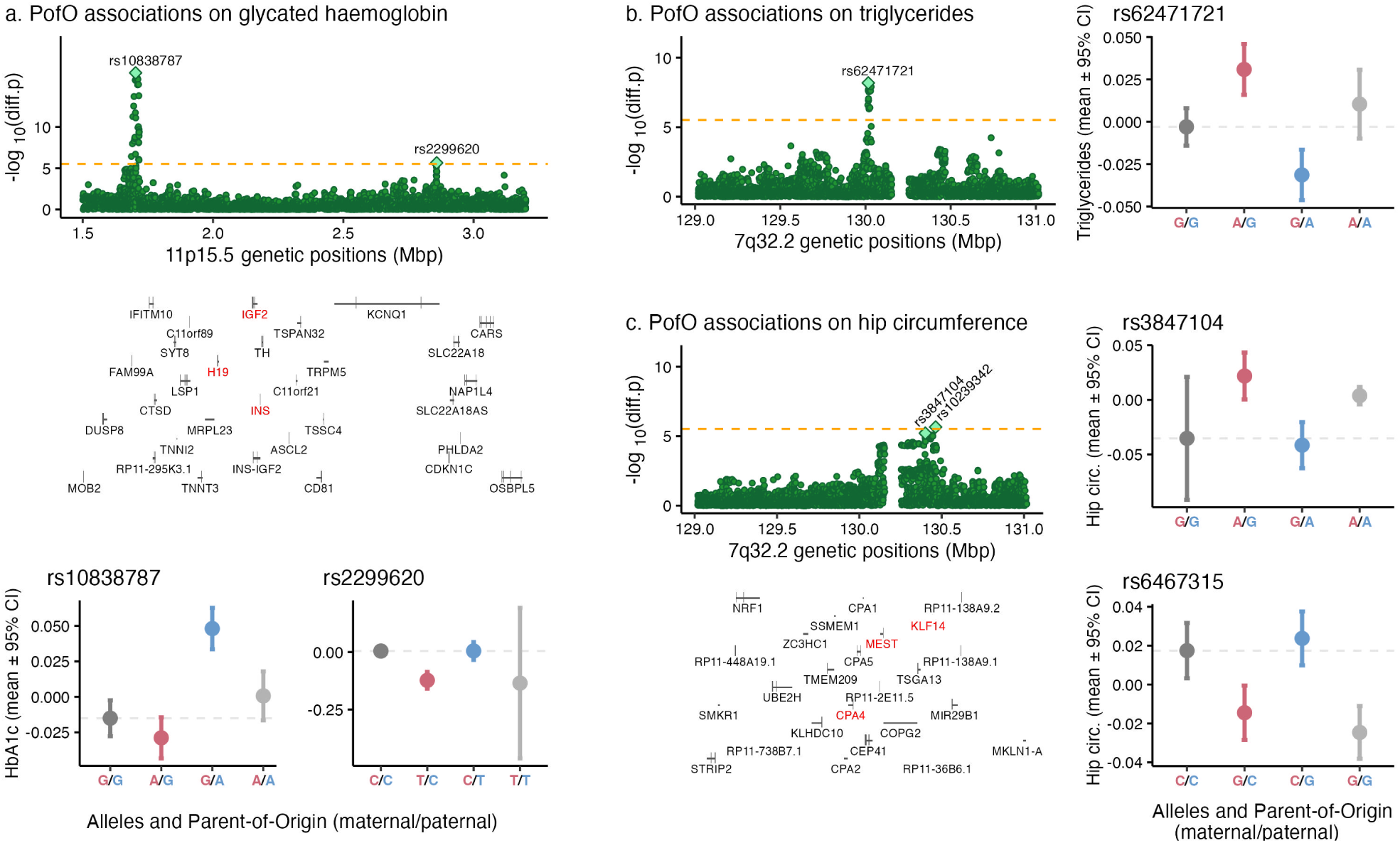
Parent-of-Origin associations at 11p15.5 and 7q32.2. Differential GWAS show the association strength (*−log*_10_(*p − value*), y-axis) against the genomic position (x-axis) on **a**) HbA1c, **b**) triglycerides, and **c**) hip circumference. Each point represents a genetic variant. Diamonds indicate variants with independent POE found in this study and listed in Table 1. Orange dashed line represents the significance threshold used in this study when focusing on imprinted regions (3.1 *×* 10^−06^). For **a**) and **c**), gene positions are shown along the genomic positions (x-axis). Horizontal lines show the gene start and end positions. Vertical lines show the exons start and end positions. Gene names are shown below the corresponding gene coordinates. Gene positions for **c**) are aligned to those in **b**). The independent lead POE variants are represented within each GWAS plot (light green diamonds). For each independent POE, we also show the genotypes and PofO of alleles (x-axis) effects on trait level (y-axis). Red and blue alleles indicate maternal and paternal alleles, respectively. Dots show mean values, error bars show 95% confidence intervals. Grey dashed lines represent the mean value of individuals carrying no alternative allele (i.e., genotype 0). Dots and bars colors indicate maternal heterozygotes (red), paternal heterozygotes (blue), and homozygotes (dark and light grey).

We identified a novel, independent POE of rs2299620 on HbA1c (*P_D_*= 2.2 × 10^−6^), a variant located 1.1 Mb away and not in LD with the previously discussed variants (*r*^2^ *<* 0.01, Table 1, Figures 4A). The maternal T allele was associated with decreased HbA1c levels (*β̂_M_* = −0.12, *P_M_* = 1.3 × 10^−13^), while the paternal T allele has no detectable effect. Here again, while being novel for HbA1c, the variant is in high LD (*r*^2^ = 0.5) with rs2237892, a variant previously associated with T2D^5^. Since HbA1c is a known proxy for T2D^17^, we also examined the POE of rs2299620 on T2D. Although not reaching our conventional significance threshold, we observed a moderate protective effect of the maternal T allele (*β̂_M_* = −0.36, *β̂_P_*= −0.01, *P_M_* = 3.2 × 10, *P_P_* = 0.42, *P_D_* = 2.2 × 10^-3^ ). To disentangle the observed signal at this locus, we performed a conditional analysis for HbA1c, which revealed that the two SNPs are not independently associated with HbA1c. SNP rs2299620 emerged as the lead variant driving the observed HbA1c association at this locus (*P_D_c__* 0.37 for rs2299620 and rs2237892, respectively). = 8.1 × 10^−5^ and 0.37 for rs2299620 and rs2237892, respectively).

#### Novel bi-polar dominant effects on renal function indicators at 11p15.5

We identified three novel POEs on cystatin C, creatinine and urate levels (*P_D_* = 5.9 × 10^−14^, 4.9×10^−11^ and 2.2×10^−7^, respectively, Table 1, Figure 2) at a locus located over 300kb away and not in LD with other known POEs on blood biomarkers (*r*^2^ *<* 0.001). We observed bi-polar effects of lead SNPs rs170102 and rs217215 (*r*^2^ *>* 0.98) for both cystatin C and creatinine, respectively. Specifically, we found the paternal A allele of rs170102 associated with decreased, but the maternal A allele associated with increased cystine C levels (*β̂_P_* = −0.041, *β̂_M_* = 0.026, *P_P_* = 5.9×10^−11^, *P_M_* = 3.1×10^−5^). We also found the paternal G allele of rs217215 associated with increased, but the maternal G allele associated with decreased creatinine levels (*β̂_P_* = 0.03, *β̂_M_* = −0.024, *P_P_* = 2.5 × 10^−7^, *P_M_* = 2.6 × 10^−5^). For urate, we found the G allele of rs4264135 associated with decreased urate levels when paternally inherited, and observed no effect when maternally inherited (*β̂_P_* = −0.033, *β̂_M_* = 0.014, *P_P_* = 1.8 × 10^−7^, *P_M_* = 0.033).

Given the extensive pleiotropy observed at 11p15.5, including POEs on urate, cystatin C, creatinine, standing height, and metabolic traits (Figure 2), we further performed co-localization analysis within a 1Mb region surrounding these signals (Supplementary Figure 17A). This analysis identified two distinct clusters of traits likely driven by two distinct causal variants. The first cluster included cystatin C and creatinine, which exhibited a high posterior probability of sharing a causal variant (*H*_4_ *>* 0.92). The second cluster comprised standing height and metabolic traits, which also showed high probability to share the causal variant (*H*_4_ *>* 0.92). In contrast, urate showed only moderate co-localization probabilities with all other traits, with *H*_4_ ranging from 0.27 to 0.47 (Supplementary Figure 17B). These findings highlight the complexity of genetic regulation at 11p15.5 and its diverse impact on multiple phenotypes.

#### 7q32.2 revealed novel bi-polar dominant effects on lipid-related profiles

We identified novel POEs of rs62471721, rs4731690 and rs10239342 on triglycerides (TG), HDL-cholesterol (HDL-C) and sex hormone-binding globulin (SHBG) levels, respectively (*P_D_* = 6.5 × 10^−9^, 1.9 × 10^−6^ and 1.8 × 10^−6^, respectively, Table 1). These variants are in moderate to high LD (*r*^2^ *>* 0.4) with each other, but are not in LD with other previously reported POE associations (*r*^2^ *<* 0.001). These novel POEs revealed clear bi-polar dominant effects, where parental alleles influence traits in opposite directions (Figure 2).

We found the maternal A allele of rs62471721, which is an eQTL for the imprinted genes *KLF14*, *MEST*, *COPG2* and *CPA4* in adipose tissue^16^, to be associated with increased, and the paternal A allele with decreased TG levels (*β̂_P_*= −0.023, *β̂_M_* = 0.033, *P_P_*= 9.7 × 10^−4^, *P_M_* = 1.5 × 10^-6^ , Figure 4B).

We also identified a bi-polar dominant POE of rs4731690 on HDL-cholesterol levels, a variant also influencing *MEST*, *COPG2* and *CPA4* expression levels in adipose tissue^16^. Specifically, we found the paternal G allele to be associated with increased, but the maternal G allele associated with decreased HDL-C levels (*β̂_P_* = 0.019, *β̂_M_* = −0.022, *P_P_* = 1.9 × 10^−3^, *P_M_* = 2.7 × 10).

Lastly, we found a bi-polar dominant POE of rs10239342, also eQTL for imprinted genes *KLF14*, *MEST* and *COPG2* ^16^, on SHBG levels, with the paternal G allele associated with increased, but the maternal G allele associated with decreased SHBG level (*β̂_P_* = 0.022, *β̂_M_*= −0.024, *P_P_* = 1.3 × 10, *P_M_* = 4.1 × 10).

We evaluated the independence of these effects using co-localization analysis, which found that all three traits (HDL-C, TG and SHBG) likely share the same causal POE variant (posterior probability *H*_4_ *>* 0.99 for HDL-TG and SHBG-TG, *H*_4_ = 0.75 for HDL-SHBG).

#### Novel maternal effects on hip circumference at 7q32.2

We discovered a novel POE of rs6467315 on hip circumference, a variant eQTL for the maternally expressed gene *KLF14* in multiple tissues, located within the 7q32.2 imprinted region (Table 1, Figure 4C). We found the maternal G allele of rs6467315 associated with increased hip circumference and we observed no effect of the paternal G allele (*β̂_M_* = −0.042, *β̂_P_* = 0.001, *P_M_* = 5.8 × 10^−11^, *P_P_* = 0.9, *P_D_* = 2.2 × 10^−6^). The variant is not in LD with the previous POEs we reported on TG, HDL-C and SHBG above (*r*^2^ *<* 0.001), but is in high LD (*r*^2^ = 0.96) with other variants exhibiting maternal effects on TG, HDL-C, and T2D reported by previous studies (see replication section, Supplementary Table 2)^4,5^. Consistently, we found moderate associations when testing rs6467315 for POEs on these traits (*P_D_* = 6.9 × 10^−3^, 8.9 × 10^−4^ and 4.6 × 10^−4^ for T2D, TG and HDL-C, respectively), with maternal effects aligning to those previously reported. For example, we found the maternal G allele of rs6467315 to be associated with increased T2D prevalence, while the paternal G allele had no effect (*β̂_M_* = 0.088, *β̂_P_* = −0.010, *P_M_*= 6.6 × 10^−4^, *P_P_* = 0.69). Also at 7q32.2, the conditional analysis allowed us to discover a novel POE, rs3847104, independent of rs6467315 (*r*^2^ = 0.012), located only 62kb away from it. The variant was also not in LD with previous POEs we reported on TG, HDL-C and SHBG above (*r*^2^ *<* 0.001). We found the maternal A allele of rs3847104 associated with an increase in hip circumference and did not observe an effect of the paternal G allele (*β̂_M_c__* = 0.05, *β̂_P_c__* = −0.016, *P_M_c__* = 1.9 × 10^−08^, *P_P_* = 0.1, *P_D_c__* = 3.8 × 10^−7^, Figures 4C, Supplementary Figure 18).

#### A novel locus with bi-polar dominant effects on body fat composition at 20q13.32

We identified a novel POE of rs80116540 on body fat percentage within the 20q13.32 imprinted region. The paternal G allele was associated with increased in body fat percentage, while the maternal G allele showed the opposite effect (*β̂_P_* = 0.037, *β̂_M_* = −0.037, *P_P_* = 2.6 × 10^−4^, *P_M_* = 2.2 × 10^−4^, *P_D_* = 2.0 × 10^−7^). To assess which parts of body fat this effect is driven by, we examined its POE on arm-, leg-, and trunk fat percentages. Significant POEs were observed for all these traits (*β̂_P_* = 0.025, 0.035, and 0.042; *β̂_M_* = −0.024, −0.042, and −0.039; *P_P_* = 2.8 × 10^−3^, 4.3 × 10^−4^, and 3.6 × 10^−4^; *P_M_* = 3.5 × 10^−3^, 2.0 × 10^−5^, and 1.0 × 10^−3^; *P_D_* = 2.9 × 10^−5^, 3.4 × 10^−8^, and 1.3 × 10^−6^ for leg, arm, and trunk fat percentages, respectively). Notably, these traits met the significance threshold specific to imprinted regions at this locus, though some exhibited different lead variants (Table 1). These lead variants were identified as either sQTLs for *STX16* or *CTSZ* in adipose tissue^16^ or eQTLs for the *GNAS* imprinted gene in blood^18^, underscoring their potential functional relevance in the observed POEs.

#### Novel and distinct maternal and paternal effects on IGF-1 at 6q24.2 and 16p13.3

We identified a novel POE of rs12528876 on IGF-1 levels at the 6q24.2 locus, located in an intron of the *PLAGL1* gene (*P_D_* = 2.9 × 10^−6^, Table 1). Specifically, we found the paternal C allele of rs12528876 to be associated with decreased IGF-1 levels and the maternal allele having no effect (*β̂_P_* = −0.036, *β̂_M_* = 0.021, *P_P_* = 2.5 × 10^−5^, *P_M_* = 0.016).

We identified a second independent and novel POE of rs7188903 on IGF-1 at 16p13.3 (*P_D_* = 6.1 × 10^−7^, Table 1), which is an eQTL in blood for two maternally expressed genes, *ZNF597* and *NAA60*. We found the maternal G allele of rs7188903 to be associated with increased IGF-1 levels and the paternal allele having no effect (*β̂_M_* = 0.041, *β̂_P_* = −0.012, *P_M_* = 5.3 × 10, *P_P_* = 0.1).

#### Novel bi-polar dominant effect on total protein levels at 15q12

We identified a novel POE of rs146982369 on total blood protein levels within the 15q12 imprinted region, located in the intro of *ATP10A* (*P_D_* = 2.5 × 10^−6^, Table 1). We found the paternal G allele of rs146982369 associated with an increase in blood protein levels, while the maternal G allele has the opposite effect (*β̂_P_* = 0.101, *β̂_M_* = −0.106, *P_P_* = 1.8 × 10^−3^, *P_M_* = 9.9 × 10).

#### A known maternal effect on platelet count at 14q32.2

Lastly, we identified a POE of rs59228823 on platelet count, an association that was already reported in our previous study^3^. This variant has been mapped to the *MEG3* maternally expressed gene, and is in high LD (*r*^2^ *>* 0.55) with rs12881545, a known maternal eQTL of *MEG3* ^19^.

### Parent-of-origin specificity of additively associated regions

Another way to reduce multiple testing burden in a meaningful way is to restrict our POE analysis to variants exhibiting an additive effect on the examined traits, as captured by traditional GWAS. For simplicity, we refer to these as “additively associated regions” in subsequent sections. To identify novel POEs within additively associated regions, we first performed additive association testing (ignoring the PofO of the alleles) for 59 selected traits (Supplementary Table 1) and identified 1,812 significant (additive P-value *<* 1 × 10^−9^, see Methods) LD-pruned additive associations. We then sought to detect whether some of these associations may involve POEs. To this end, we applied a differential POE GWAS scan (see Methods) restricted to these SNP-trait pairs. This approach is geared towards the identification of POEs where the maternal and paternal effects are not in opposite direction and hence even the simple additive effect is non-zero. While this strategy allows us to detect only a subset of POE scenarios (Supplementary Figure 15), it also drastically reduces the multiple testing burden, setting the significance threshold to 0.05*/*1812 = 2.75 × 10^−5^ (see Methods).

This analysis identified six significant POEs (Table 1, Figure 2). Of these, two were novel associations, both with telomere length (Figure 5). First, the C allele of rs2293607 on chromosome 3, located within the *TERC* gene (exon of noncoding transcript), was found to be associated with decreased telomere length when inherited from either parent, although the effect is significantly stronger when inherited paternally (*β̂_P_* = −0.12, *β̂_M_* = −0.068, *P_P_* = 2.2 × 10, *P_M_* = 5.3 × 10, *P_D_* = 5.7 × 10^-7^ , Figure 5A,B). Furthermore, the T allele of rs11100479 on chromosome 4, an eQTL for the gene *NAF1*, was associated with an increase in telomere length when inherited from the father. We also observed a moderate effect of the maternal T allele, albeit the effect of the paternal T allele was significantly stronger (*β̂_P_* = 0.078, *β̂_M_* = 0.022, *P_P_* = 1.8 × 10^−22^, *P_M_* = 6.6 × 10^−3^, *P_D_* = 6.25 × 10^−7^, Figure 5A,C).

**Fig. 5.**
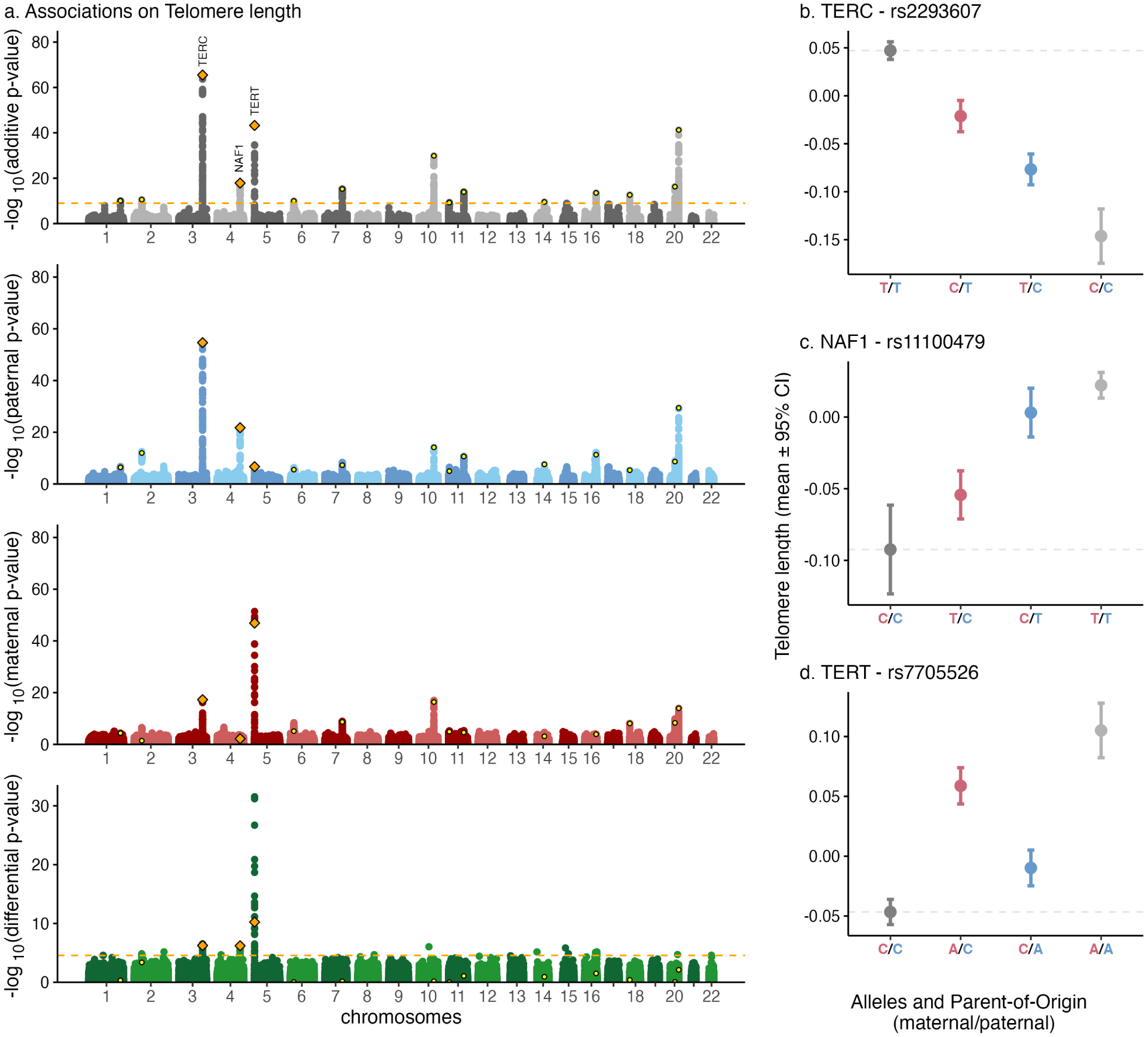
Parent-of-origin associations on telomere length. **a**) Genome-wide associations for additive (grey), paternal (blue), maternal (red), and differential (green) effects represented as association strength (*−log*_10_(*p − value*), y-axis) against the genomic position (x-axis). Each point represents a genetic variant. Yellow dots indicate additive genome-wide significant variants (those then tested for POE). Orange diamonds indicate variants exhibiting significant POE among the lead additive effect variants. Orange dashed lines indicate significant thresholds (1 *×* 10*^−^*^9^ for the additive effects and 3.1 *×* 10*^−^*^6^ for POEs). **b-d**) Genotypes and PofO of alleles (x-axis) effects on telomere length (y-axis). Red and blue alleles indicate maternal and paternal alleles, respectively. Dots show mean values, and error bars show 95% confidence intervals. Grey dashed lines represent the mean value of individuals carrying no alternative allele (i.e., genotype 0).

Two additional associations were found in moderate to low LD with previously reported variants. First, at the 5p15.33 locus, the POE of rs7705526 on telomere length exhibited moderate LD (*r*^2^ = 0.27) with rs2735940, a variant previously implicated in telomere length in our earlier work^20^. This reflects the limitations of the additively associated regions approach, which assesses the POE specificity of the lead additive variant but may not always pinpoint the true lead POE variant. Interestingly, rs2735940 was also identified as the lead POE variant for telomere length in the genome-wide analysis presented below (Figure 5A,D). Second, at the 11p15.5 region, we identified maternal effect of rs2237895 on HbA1c. This variant was in low LD (*r*^2^ = 0.025) with rs2299620, previously implicated in our imprinted region-focused analysis. Conditional analysis suggested that neither variant met the significance threshold independently (*P_Dc_* = 3.5 × 10^−4^ and 4.3 × 10^−5^ for rs2237895 and rs2299620, respectively), therefore we cannot distinguish between these two signals The remaining two associations - with standing height at 11p15.5 and with platelet count at 14q32.2 - were already identified in our imprinted region-focused analysis (see Table 1).

In less stringent analysis, where the testing burden is reduced by evaluating each trait individually, we identified six additional suggestively significant POEs (Supplementary Table 5), two of which (both novel) with IGF-1. The maternal T allele of the first lead variant rs17145750, intronic to *MLXIPL*, was associated with increased IGF-1 levels, and we observed no effect from the paternal T allele (*β̂_P_* = 0.015, *β̂_M_* = 0.063, *P_P_* = 0.09, *P_M_* = 9.07 × 10^−13^, *P_D_* = 1.18 × 10^−4^). The second association is located within the 11p15.5 imprinted region. The lead variant rs3213217, intronic to *IGF2*, demonstrated asymmetric polar effects, with both maternal and paternal T alleles associated with increased IGF-1 levels, albeit to different extents (*β̂_P_* = 0.096, *β̂_M_* = 0.058, *P_P_* = 1.4×10^−34^, *P_M_* = 2.0×10^−13^ *P_D_* = 5.7 × 10^−4^). While these findings did not meet the stringent significance threshold when focusing on additive regions, they highlight potential POEs with moderate effects.

### Parent-of-origin association genome-wide

We finally conducted a genome-wide scan for POEs across the 59 selected traits (Supplementary Table 1), identifying 6 significant POE associations (differential P-value *P_D_ <* 1 × 10^−9^, see Methods, Table 1, Figure 2, Supplementary Table 6A). However, this approach did not reveal any additional independent associations beyond those identified through the targeted strategies presented above.

By reducing the testing burden and analyzing each trait independently, we identified 11 additional suggestively significant POEs (differential P-value *P_D_ <* 5 × 10^−8^). Of these, 4 overlapped with findings from the targeted strategies (for triglycerides, T2D, leg fat-free mass, and arm fat percentage), while the remaining 7 were novel (Supplementary Table 6B). For example, we found a novel suggestive POE of rs145976519 on birth weight. This variant, located at 1p34.2, lies outside known imprinted regions and shows no additive association with birth weight. The C allele was associated with decreased birth weight when inherited maternally, while the paternal C allele increased it. Beyond its role as an eQTL for *CAP1* and *PPT1*, rs145976519 is located approximately 130kb from *ZMPSTE24* and modulates the expression of its divergent transcript^18^. Notably, *ZMPSTE24* has been implicated in neonatal pathologies, including intrauterine growth retardation and prematurity.

### Parent-of-origin effects in early life

#### Comparative childhood traits at age 10 in the UK Biobank

We examined the timing of POEs on complex traits to assess whether these effects manifest in early childhood or are specific to adult traits. For this purpose, we tested three loci associated with standing height for significant POEs on “comparative body height at age 10” and seven loci linked to obesity-related traits for POEs on “birth weight” and “comparative body size at age 10.” All three loci associated with adult standing height also exhibited significant POEs on “comparative body height at age 10” (significance threshold = 0.05*/*(3 + 7 × 2) = 2.9 × 10^-3^ , Supplementary Table 7). Specifically, the paternal T allele of rs576603 and the paternal G allele of rs77708343, both near *IGF2*, were associated with decreased body height at age 10, while the maternal alleles of these SNPs led to an increase. Additionally, the maternal T allele of rs143840904 (intronic to *KCNQ1*) was associated with a decrease in body height at age 10, whereas the paternal T allele had no significant effect. Notably, these observed effects on body height at age 10 align with those seen in adult standing height, suggesting that POEs at these loci may influence early growth trajectories with long-lasting effects.

#### Early-life BMI dynamics in the MoBa cohort

To further explore these findings, we analyzed BMI in early life using data from the Norwegian Mother, Father and Child Cohort Study (MoBa)^21^. We used 45,402 children with both parents and longitudinal BMI measurements from 6 weeks to 8 years of age^22^. In this cohort, we tested the seven obesity-associated loci we found in the UK Biobank for association with BMI at 11 time points (Supplementary Table 8). We identified seven significant POEs (*P_D_ <* 1.67 × 10^−3^, see Methods), all involving the SNP rs6467315. Specifically, we found the maternal G allele of rs6467315 associated with increased BMI during infancy (6 weeks to 2 years, *β̂_M_* = 0.057 to 0.089, *P_M_* = 5.9 × 10^−4^ to 9.4 × 10^−7^), while the paternal allele showed no effect. The maternal effect was strongest at 6 months and gradually attenuated, becoming undetectable by the age of 3 years (Supplementary Table 9, Supplementary Figure 19). This attenuation likely explains the absence of a detectable effect at age 10 in the UK Biobank cohort.

Interestingly, the maternal G allele had an opposite direction of effect on adult traits, where it decreased both hip circumference (see above, Table 1) and BMI (*β̂_M_* = −0.02, *β̂_P_* = 0.006, *P_M_*= 2.6 × 10, *P_P_*= 0.33, *P_D_* = 1.0 × 10^-3^ ). We performed co-localization analysis, confirming that the same causal variant likely drives both traits at this locus (posterior probability H4 *>* 0.96).

These findings align with earlier research on rs287621 (*r*^2^=0.36 with rs6467315), which reported similar opposing maternal effects on BMI across infancy and adulthood^22^. Consistently, we also find a moderate maternal effect of this variant on hip circumference, with the maternal T allele decreasing hip circumference, and no observable effect of the paternal C allele (*β̂_M_* = −0.035, *β̂_P_* = −0.003, *P_M_* = 9.3 × 10^−7^, *P_P_* = 0.61, *P_D_* = 1.8 × 10^−3^). We, however, found no POE of this variant on adult BMI in the UK Biobank cohort (*β̂_M_* = 0.016, *β̂_P_* = 0.001, *P_M_* = 0.023, *P_P_* = 0.82, *P_D_* = 0.14).

To disentangle the observed signal at this locus, we performed a conditional analysis for both adult hip circumference and BMI, which revealed that our lead POE variant, rs6467315, is the one driving the observed associations at this locus (hip circumference *P_D**c**_* = 3.6 × 10^−3^ and 0.84; BMI *P_Dc_* = 9.7 × 10^−3^ and 0.49, for rs6467315 and rs287621, respectively).

We further confirmed that the observed maternal effect on early-life BMI was not attributable to maternal untransmitted alleles (Supplementary Figure 19, Supplementary Table 9), ruling out maternal rearing effects as the underlying mechanism and consistent with imprinting effects.

### Evidence of sex-specific bi-polar POE on glucose levels

Recognizing that complex traits often exhibit sex-specific variation^23,24^, we investigated whether POEs also display sex-specific patterns. To explore this, we conducted POE analyses separately for males and females across the 30 POE associations identified in this study and listed in Table 1.

We identified one significant sex-specific difference, for the POE of rs4417225 (near *H19* /*IGF2*) on glucose levels (Supplementary Table 10, Supplementary Figure 20). The POE was significant in male individuals (*P_Dmales_* = 4.7 × 10^−9^), but not in female individuals (*P_Dfemales_* = 0.19). Specifically, the T allele of rs4417225 was associated with increased glucose levels in men when paternally inherited, and with decreased levels when maternally inherited (*β̂_P_* = 0.045, *β̂_M_* = −0.040, *P_P_* = 1.0^−4^, *P_M_* = 1.3 × 10^−5^), while we observed no effect of neither paternal nor maternal T alleles in women (*P_P_* = 0.3, *P_M_* = 0.38). Additionally, we formally tested for significant sex differences in POE by comparing male and female GWAS effect sizes using a Z-score approach, applying a significance threshold of *P_Z_ <* 0.05*/*30 = 1.67 × 10^−3^ (see Methods). This analysis confirmed a significant sex-specific POE for glucose levels (*P_Z_D__* = 1.3 × 10^−3^, Supplementary Table 10). Interestingly, although this locus is also associated with HbA1c and type 2 diabetes risk in a POE fashion, no sex-specific effects were detected for these traits (Supplementary Table 10).

### Parent-of-Origin SNP heritability for complex traits

We used Linkage Disequilibrium Score Regression (LDSC) to estimate SNP heritability (h²), which represents the proportion of phenotypic variability explained by Single Nucleotide Polymorphisms (SNPs). Instead of the traditional SNP-based h² computation that considers additive effects, we estimated *h*^2^ separately for the maternal effects, the paternal effects, and for the parental differential effects at heterozygous sites (as derived from our maternal, paternal and differential GWAS scans, respectively; see Methods).

Our genome-wide estimates reveal that for most traits, the paternal and maternal haplotypes contribute similarly to SNP-based heritability (Figure 6A). Next, we focused our analysis within imprinted regions. While the paternal and maternal contributions remained balanced for most traits, we observed several interesting trait-specific patterns (Figure 6B): for arm fat percentage and type 2 diabetes, we found a trend for larger maternal *vs* paternal SNP heritability, while we found larger point estimates for paternal SNP heritability for birth weight, glucose and basophil count.

**Fig. 6.**
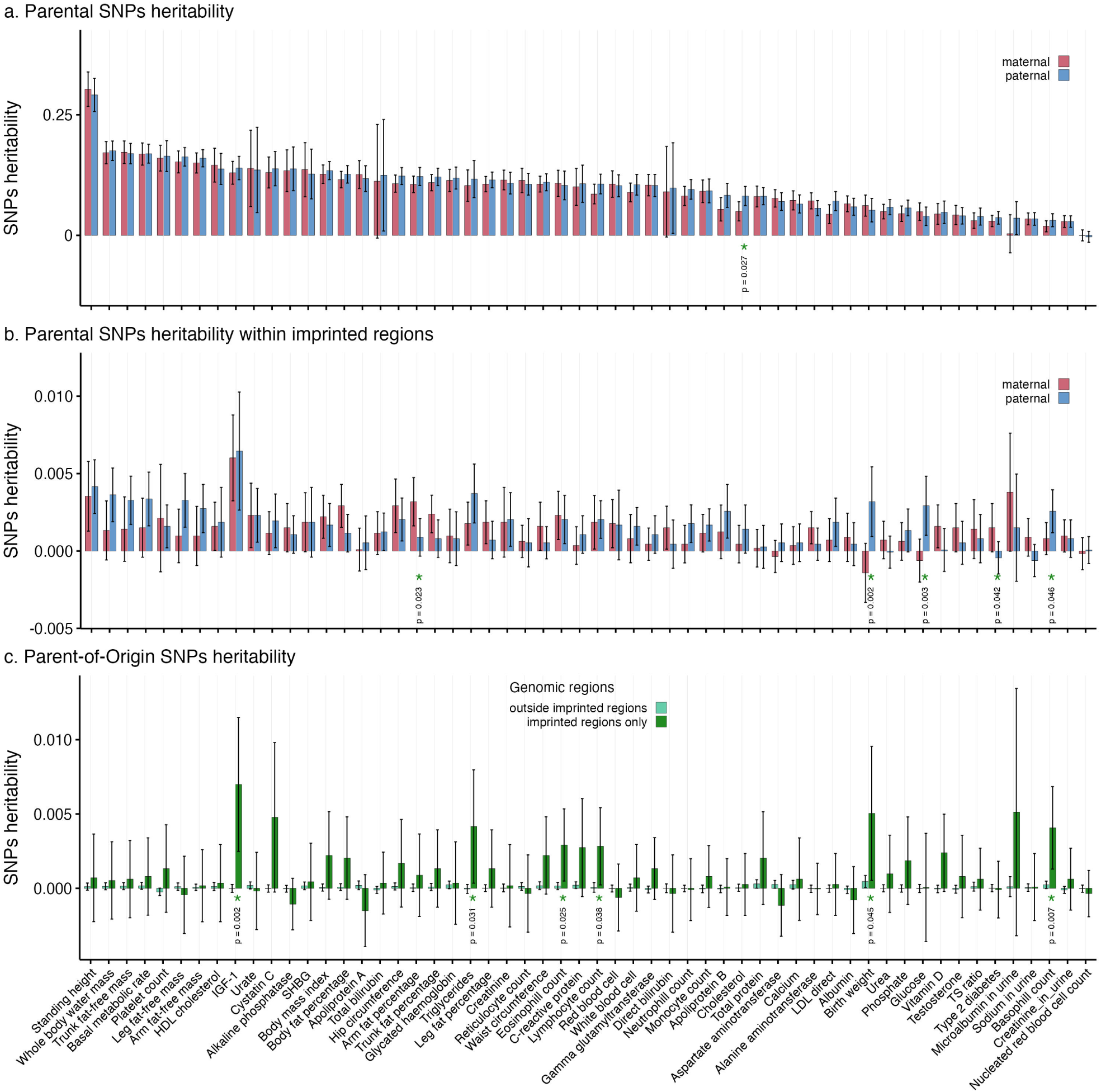
SNPs heritability. SNPs heritability (*h*^2^, y-axis) across 56 traits tested (x-axis). Paternal (blue) and maternal (red) *h*^2^ was estimated (**a**) genome-wide and (**b**) within imprinted regions. Stars and p-values indicate nominal significant differences between paternal and maternal *h*^2^. **c**) *h*^2^ was estimated within and outside imprinted regions. Stars and p-values indicate nominal significant differences between 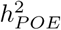 within imprinted regions *vs* outside imprinted regions. Three out of the 59 selected traits were not represented here due to disproportionate standard errors: lipoprotein A, rheumatoid factor, and oestradiol.

Finally, we estimated POE SNP heritability (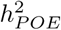) within and outside imprinted regions. Our findings revealed that six traits exhibited significantly higher 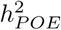 within imprinted regions compared to non-imprinted regions (Figure 6C), namely for IGF-1, triglycerides, eosinophil count, lymphocyte count, birth weight and basophil count. Reassuringly, no traits showed significantly greater 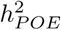 outside known imprinted regions, hinting that a disproportionately large POE heritability is due to genomic imprinting.

### Parent-of-Origin Protein Quantitative Trait Loci

In addition to our investigation of POEs on complex traits, we explored the UK Biobank plasma proteomics data to identify parent-of-origin protein quantitative trait loci (POE- pQTLs). Since protein levels widely correlate with transcript abundance, we hypothesized that imprinting mechanisms acting on gene expression levels may detectably propagate to protein levels. Our analysis focused on two main objectives: first, determining whether variants exhibiting POEs on complex traits identified in this study also exhibit POE- pQTL effects on their associated genes, and second, assessing whether previously established pQTLs^25^ exhibit associations in a PofO fashion.

#### Parent-of-Origin pQTLs at known POEs

Across the genes associated with variants exhibiting POEs on complex traits identified in this study (Table 1), two genes, *CPA4* and *GNAS*, had available protein quantification data. To investigate potential POEs at the protein level, we tested eight variant-protein pairs: rs10239342, rs62471721, rs4731690, rs6467315 and rs3847104 with *CPA4*, and rs80116540, rs6026426, and rs3730173 with *GNAS*. Among these, we identified one significant (0.05/8) POE-pQTL for *CPA4*. Specifically, the G allele of rs4731690, previously identified as exerting a bi-polar effect on HDL-C, was found to increase *CPA4* protein levels only when paternally inherited (*β̂_M_* = −0.01, *β̂_P_* = 0.08, *P_M_* = 0.58, *P_P_* = 6.3×10^−4^, *P_D_* = 5.7×10^−3^). In contrast, no significant POE-pQTLs were observed for *GNAS*.

Co-localization analysis revealed that the observed POE signals for HDL-C and *CPA4* are distinct (posterior probability *H*_4_ = 0.04).

#### Novel Parent-of-Origin pQTLs among additive pQTLs

Beyond exploring POE-pQTLs at variants regulating complex traits in a parent-of-origin fashion, we also evaluated a set of established additive pQTLs^25^ for potential POEs. From the 14,287 lead pQTL associations spanning 2,414 proteins reported in a previous study^25^, we could directly test 10,611 associations in our dataset, where the same SNP and protein data were available, and we used proxy variants selected based on LD (*r*^2^ *>* 0.8) for the remaining ones. We identified four significant (*P_D_ <* 0.05*/*14285 = 3.5 × 10^−6^) POE associations, with namely *DLK1*, *CPA4*, *ADAM23* and *PER3* protein levels (Table 2, Supplementary Figure 21).

**Table 2.** Significant Parent-of-Origin protein QTLs. chromosome; GENPOS: genetic position (hg19); ID: variant rs id; A0: reference allele; A1: assessed allele; EQ: A1 allele frequency. BETA, SE and P denote effect sizes, standard errors and P-values; PAT, MAT, DIFF DD denote paternal, maternal, differential and additive tests. PHENAME: phenotype name.

| CHR | POS | SNP ID | A0 | A1 | A1FREQ | BETA PAT | SE PAT | P PAT | BETA MAT | SE MAT | P MAT | BETA DIFF | SE DIFF | P DIFF | BETA ADD | SE ADD | P ADD | TRAIT | STATUS |
| --- | --- | --- | --- | --- | --- | --- | --- | --- | --- | --- | --- | --- | --- | --- | --- | --- | --- | --- | --- |
| 14 | 101176212 | rs12881545 | G | C | 0.668 | 0.826 | 0.023 | 2.26E-274 | 0.187 | 0.023 | 1.24E-15 | 0.642 | 0.033 | 3.03E-84 | 0.505 | 0.016 | 2.34E-206 | DLK1 | primary pQTL and POE-pQTL |
| 7 | 129938598 | rs34587586 | G | T | 0.404 | -0.434 | 0.024 | 2.77E-71 | -0.634 | 0.024 | 4.44E-149 | 0.201 | 0.034 | 5.60E-09 | -0.532 | 0.017 | 1.53E-210 | CPA4 | primary pQTL |
| 7 | 129940561 | rs12538292 | G | A | 0.327 | -0.432 | 0.025 | 4.85E-65 | -0.667 | 0.025 | 2.32E-151 | 0.238 | 0.036 | 3.51E-11 | -0.548 | 0.018 | 7.76E-205 | CPA4 | primary POE-pQTL |
| 2 | 207311349 | rs72933203 | T | A | 0.104 | 1.017 | 0.035 | 1.45E-190 | 0.723 | 0.034 | 9.02E-102 | 0.293 | 0.048 | 1.29E-09 | 0.865 | 0.024 | 1.01E-281 | ADAM23 | primary pQTL |
| 2 | 207413625 | rs13429599 | G | A | 0.270 | 0.719 | 0.024 | 5.93E-204 | 0.457 | 0.024 | 9.30E-84 | 0.263 | 0.034 | 4.34E-15 | 0.576 | 0.017 | 4.18E-267 | ADAM23 | primary POE-pQTL |
| 1 | 7914177 | rs228647 | G | A | 0.570 | 0.506 | 0.023 | 6.87E-110 | 0.241 | 0.023 | 4.13E-26 | 0.264 | 0.032 | 1.95E-16 | 0.375 | 0.016 | 7.50E-120 | PER3 | primary pQTL |
| 1 | 7856346 | rs228682 | T | C | 0.401 | 0.540 | 0.023 | 4.34E-121 | 0.241 | 0.023 | 1.09E-25 | 0.297 | 0.032 | 5.41E-20 | 0.395 | 0.016 | 4.32E-128 | PER3 | primary POE-pQTL |

Two of these associations involved known imprinted genes: *DLK1* and *CPA4*. Specifically, we found the paternal C allele of rs12881545 associated with higher *DLK1* protein levels compared to the maternal C allele, consistent with the paternal expression of the *DLK1* gene (*β̂_P_* = 0.82, *β̂_M_* = 0.18, *P_P_* = 2.2 × 10^−274^, *P_M_* = 1.2 × 10^−15^, *P_D_* = 3.0 × 10^−84^, Table 2, Supplementary Figure 21A). For *DLK1*, the lead pQTL reported in the previous study was also our lead POE-pQTL. In addition, we found the maternal T allele of rs34587586 associated with lower *CPA4* protein levels compared to the paternal T allele, consistent with the maternal expression of the *CPA4* gene (*β̂_M_* = −0.63, *β̂_P_* = −0.43, *P_M_* = 4.4 × 10^−149^, *P_P_* = 2.7 × 10, *P_D_* = 5.6 × 10^-9^ ). For *CPA4*, the lead pQTL reported in the previous study was not our lead POE-pQTL: we found a stronger POE association with *CPA4* protein levels for another variant, rs12538292, in high LD with rs34587586 (*r*^2^ = 0.77). Specifically, we found the maternal A allele of rs12538292 associated with lower *CPA4* protein levels compared to the paternal one (*β̂_M_* = −0.66, *β̂_P_* = −0.43, *P_M_* = 2.3 × 10^−151^, *P_P_* = 4.8 × 10^−65^, *P_D_* = 3.5 × 10^−11^, Table 2, Supplementary Figure 21B).

We found two POE associations where genes were located outside known imprinted regions: for *ADAM23* and *PER3* protein levels. Firstly, we found the paternal A allele of rs72933203 associated with higher *ADAM23* protein levels compared to the maternal one (*β̂_P_* = 1.01, *β̂_M_* = 0.72, *P_P_* = 1.4 × 10^−190^, *P_M_* = 9.0 × 10^−102^, *P_D_* = 1.2 × 10^−9^). For *ADAM23*, the lead pQTL reported in the previous study was not our lead POE-pQTL: we found a stronger POE association with *ADAM23* for another variant, rs13429599, located over 100 kb away from rs72933203 and in low LD with it (*r*^2^ = 0.046). For this variant, we found the paternal A allele associated with higher *ADAM23* protein levels compared to the maternal one (*β̂_P_* = 0.71, *β̂_M_* = 0.45, *P_P_* = 5.9 × 10^−204^, *P_M_* = 9.3 × 10^−84^, *P_D_* = 4.3 × 10^−15^, Table 2, Supplementary Figure 21C). In addition, we found the paternal A allele of rs228647 to be associated with higher *PER3* protein levels compared to the maternal A allele (*β̂_P_* = 0.50, *β̂_M_* = 0.24, *P_P_* = 6.8×10^−110^, *P_M_* = 4.1×10^−26^, *P_D_* = 1.9×10^−16^). Again for *ADAM23*, the lead pQTL reported in the previous study was not our lead POE-pQTL: we found a stronger POE association with *PER3* for another variant, rs228682, in moderate LD with rs228647 (*r*^2^ = 0.3). Here again, we found the paternal C allele of rs228682 associated with higher protein levels compared to the maternal C allele (*β̂_P_* = 0.54, *β̂_M_* = 0.24, *P_P_* = 4.3 × 10^−121^, *P_M_* = 1.0 × 10^−25^, *P_D_* = 5.4 × 10^−20^, Table 2, Supplementary Figure 21D).

### Replication analyses in the Estonian Biobank

To replicate the POEs identified in our study, we leveraged the high level of relatedness within the Estonian Biobank (EstBB) cohort^26^. Our analysis followed a similar approach to that applied to the UK Biobank cohort.

Among the 211,259 EstBB participants, we identified 14,063 parent-offspring trios, 37,686 parent-offspring duos, over 55,000 individuals with at least one sibling and more than 150,000 individuals with more distant relatives. Notably, 95% of individuals had at least one close relative (up to the fourth degree), which were then clustered in surrogate parent groups, allowing us to leverage inter-chromosomal statistical phasing via shared IBD haplotype segments for almost the entire cohort. This extensive relatedness in EstBB enabled an inter-chromosomal phasing error rate of only 0.56%, compared to 1% in the UK Biobank (Supplementary Figure 22).

For individuals without directly available parental genomes, we inferred the PofO of parental haplotype sets by analyzing chromosome X inheritance in male individuals (Supplementary Figure 23) and by using crossover inference combined with sex-specific recombination maps for sibling pairs (Supplementary Figures 24 and 25). Using this approach, we successfully inferred PofO of haplotypes in a total of 113,921 EstBB individuals, of which more than 85% (N=97,346) had a probability greater than 0.99, and achieving an average error rate at heterozygous sites of just 0.89%, compared to 2.06% in the UK Biobank (Supplementary Figure 26).

Due to the limited overlap of phenotypes in the EstBB cohort, we were able to test 7 POEs identified in our study using the exact same variants. For 7 additional associations, we utilized proxy variants selected based on the highest LD available in the dataset. Out of these 14 associations, we successfully replicated 10 (*P_D_ <* 0.05*/*14 = 3.35 × 10^−3^), with two additional associations at nominal significance (Supplementary Table 11).

## Discussion

We introduce a novel multi-step approach to infer the parent-of-origin (PofO) of haplotypes, providing substantial improvements over existing methods. This innovation enables the production of the largest cohort to date with PofO information, paving the way for discoveries of parent-of-origin effects (POEs) at an unprecedented scale across complex traits, diseases, and protein levels. In the scope of this study, we initially applied this approach on the UK Biobank, inferring the PofO for 123,716 individuals. For replication, we first used our approach on the Estonian Biobank, resulting in 97,346 individuals with PofO information. To further enhance the replication and validation of our findings, we also included an additional 45,402 offspring from the Norwegian Mother, Father, and Child Cohort Study (MoBa). These combined 266,464 individuals with PofO information allowed us to systematically investigate and replicate POEs accross a broad range of human traits. Notably, we identify over 30 novel POEs, underscoring the power of this approach to uncover previously undetectable genetic effects on human traits.

### Advantages and limitations of the methodology

Our novel approach addresses significant limitations of traditional methods that rely on parental genomes, known genealogies, or chromosome X sharing in male individuals, thereby limiting scalability and applicability. By contrast, our method enables PofO inference without these constraints, greatly enhancing sample sizes and improving the detection of POEs. Instead of relying exclusively on chromosome X sharing in males, we also leverage mtDNA data from whole-genome sequencing (WGS). This allows us to infer PofO in female individuals, increasing the overall sample size and allowing us to examine sex differences in POEs. However, this approach is limited to cohorts with available WGS data. To overcome this limitation, we introduced a sibling-based crossover inference method, which provides a promising alternative for datasets derived from SNP arrays alone. This strategy allows extending PofO inference to a broader range of close relatives.

Despite its advantages, our method has several limitations. The accuracy of PofO inference depends on the availability of haplotype segments shared IBD across chromosomes, which in turn is influenced by cohort structure and data availability. Sample sizes vary across chromosomes, with fewer individuals available for smaller chromosomes where detecting IBD segments is more challenging. This variability restricts the analysis power, particularly in cohorts with limited data on related individuals. This limitation is evident when comparing inferences between the UK Biobank and the Estonian Biobank. While the UK Biobank includes an average of 1.6 relatives per individual, the Estonian Biobank includes approximately 12 relatives per individual. This difference results in larger sample sizes per chromosome and reduces error rates by half in the Estonian Biobank.

Our method is also highly dependent on the degree of relatedness among individuals. Higher accuracy and call rate is achieved with closer relatives, as seen with mtDNA minor variant sharing (MVS), where second-degree relatives can be use to predict both paternal and maternal side, versus accurately predicting only maternal side when using more distant relatives. Consequently, biobanks enriched with closer familial relationships, such as the Estonian

Biobank, provide a larger proportion of the cohort with accurate PofO inference. Indeed, approximately half of the Estonian Biobank cohort had PofO probabilities exceeding 0.99, compared to only about a quarter of the UK Biobank cohort.

Another potential limitation that was previously discussed^20^ relates to consanguinity within populations. Higher levels of consanguinity could lead to individuals sharing chromosome X or mtDNA from both sides of the family, potentially introducing errors in PofO assignment. However, our observations in the Estonian Biobank do not support this hypothesis. Despite significantly higher average Runs-of-Homozygosity in the Estonian Biobank compared to the UK Biobank, we observed lower error rates and better PofO inference coverage in the Estonian cohort. This is likely due to the larger number of close relatives per individual and our rigorous clustering approach for relatives. We excluded individuals for whom relatives could not be clustered into two distinct groups or for whom group clustering was ambiguous. Together, these measures ensure the robustness of our multi-step approach, even in populations with high consanguinity.

A further weakness of our approach is that we cannot distinguish between parental rearing and true imprinting effects. The effect of a maternal transmitted allele may emerge through changing maternal behavior impacting how a child is raised (starting from intra-uterine effects to child education), or it may exert its effect directly in the offspring via suppressed gene expression. This weakness can be addressed in the future by inferring untransmitted parental effects and comparing them to that of the transmitted one. We could examine this in the MoBa cohort, where the availability of a large number of parental genomes allowed us to infer parental untransmitted alleles. We observed effects on BMI from maternally transmitted alleles, but no effects from either paternally transmitted alleles or maternally untransmitted alleles, suggesting that maternal rearing effects are unlikely to be the underlying mechanism and aligning more closely with true imprinting effects.

### Parent-of-origin on complex traits revealed a novel asymmetric pattern

The detection power of the POE analysis is comparable to that of additive GWAS: The expected test statistic for additive GWAS is 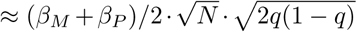. On the other hand, only heterozygous samples are informative to detect POEs, hence its test statistic is 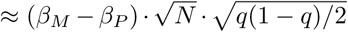, where *β̂_M_* and *β̂_P_* are the per allele maternal and paternal effect size, respectively. In classical POEs scenario, when a SNP has an effect only when it comes from one of the parents (e.g. *β̂_P_* = 0), the test statistics become equal. Note that in these situations, an additive effect should also be detectable. The fact that we detect much less POEs is therefore likely driven by the lack of classical patterns (i.e, strict paternal or maternal effect). Thus, besides the genome-wide scan, we explored two additional strategies to boost power: focus on (i) SNPs that show an additive association (since apart from the scenario of *β̂_M_* = −*β̂_P_*, the additive effect will not be zero); (ii) imprinted regions (where POEs are more likely). These filters reduce multiple testing burden and hence lead to milder P-value thresholds and can improve statistical power.

These analyses uncovered a wide range of POEs, offering insights into previously uncharacterized genetic mechanisms. Our additively associated regions-focused scan revealed two novel POEs on telomere length, the first associated with the *TERC* gene, and the second with the *NAF1* gene, both well-known regulators of telomere biology^27,28,29^. Although these associations do not fall within imprinted regions, they align with earlier hypotheses suggesting that telomere length regulation may involve imprinting mechanisms^30,31,32^. The identification of these loci broadens our understanding of telomere length regulation, suggesting that genomic imprinting mechanisms may operate beyond established imprinted regions, warranting further investigation. Notably, these POEs exhibit a pattern distinct from the current classification of POEs, termed here “asymmetric polar” parental effects: both parental alleles affect the trait in the same direction, but with significantly different magnitudes of effect (Supplementary Figure 15). This novel POE pattern may relate to the concept of incomplete imprinting, where both parental alleles are expressed but at different levels.

We also explored POEs within additively associated regions with a less stringent approach, analyzing each trait independently to reduce the multiple testing burden. Although these findings have higher false discovery rate, they suggest novel associations worth further investigation. For example, two POEs were identified on IGF-1 levels, each supported by biological evidence. The first lead variant is located within *MLXIPL*, a gene involved in glucose metabolism and type 2 diabetes. The second is located within the *IGF2* gene, which supports a role for the locus in insulin regulation pathways.

### Regulatory mechanism behind bi-polar dominant effects

Our imprinted region-focused scan underscored the complexity and prevalence of bi-polar POEs, where maternal and paternal alleles influence a trait in opposite directions. These effects were observed across a wide range of traits, including SHBG, triglycerides, HDL-C, HbA1c, glucose, standing height, cystatin C, creatinine, basal metabolic rate, leg and trunk fat-free mass, whole-body water mass, total protein, and various measures of fat percentage. Notably, while bi-polar effects have been reported previously, their mechanisms have often remained unclear. Our findings offer new insights that help clarify the genetic basis of these intriguing effects.

At the 7q32.2 imprinted region, we uncovered novel bi-polar POEs on triglycerides, HDL-C, and SHBG. These associations are particularly notable in the context of bi-polar dominance, as the lead variants act as eQTLs for both maternally expressed genes (*KLF14* and *CPA4*) and the paternally expressed *MEST*. The pleiotropic nature of these variants provides a compelling explanation for the observed bi-polar effects, with opposing parental influences arising from their simultaneous impact on genes exhibiting opposite imprinting patterns. This interplay between maternally and paternally expressed genes exemplifies the intricate regulatory mechanisms underlying bi-polar POEs.

Additionally, we uncovered examples of bi-polar POEs that are not directly explained by both paternal and maternal imprinted genes, suggesting the involvement of more complex regulatory networks. This includes for example bi-polar effects on glucose levels and type 2 diabetes at 11p15.5, within the *H19* /*IGF2* region.

### Multi-trait integration reveals broader parental contributions

One of the key advantages of our method is its scalability and applicability to biobank-scale datasets, providing access to a vast array of phenotypes. Such phenotype-rich biobanks allow in-depth POE pheWAS follow-up, which can pinpoint pleiotropic mechanisms underlying multi-trait associations and opens new avenues to follow-up, such as colocalization and Mendelian Randomization to distinguish vertical and horizontal pleiotropy.

For example, we identified POEs associated with standing height and basal metabolic rate at 11p15.5. To better understand the genetic signal at this locus, we explored its associations with several related traits, including weight, waist and hip circumference, and detailed body composition measures such as fat, fat-free, and water mass. Interestingly, while the lead variant showed no significant association with fat percentage across different body regions (arm, leg, trunk, or whole body), it was strongly associated with fat-free traits, such as fat-free mass and water mass. This pattern suggests that the genetic signal at 11p15.5 is more likely driven by height and fat-free body mass, which are closely related to lean tissue and skeletal structure, rather than by weight alone.

Another illustrative example comes from the 11p15.5 locus, where we identified POEs on cystatin C, creatinine, and urate levels. Both cystatin C and creatinine are well-established markers of kidney function, and colocalization analyses provided strong evidence for a shared genetic basis for these traits at this locus, likely related to renal filtration capacity. In contrast, urate, a byproduct of purine metabolism that is also filtered by the kidneys, exhibited lower colocalization probabilities with cystatin C and creatinine. This finding suggests that urate regulation, while partly overlapping with GFR-related traits, likely involves additional distinct pathways. Together, these three POEs link this locus to renal functions.

Lastly, we observed POEs on triglycerides (TG), HDL-C, and SHBG at the 7q32.2 region. These traits are established indicators of metabolic health and are closely tied to insulin resistance and metabolic syndrome. Insulin resistance typically raises TG levels while lowering HDL-C and SHBG levels, a pattern that aligns with the observed POEs. Despite the known associations of these traits with T2D risk, no significant associations were detected between the identified variants and T2D, pointing to independent regulatory mechanisms, likely specific to lipid and hormonal pathways.

### Evolutionary insights supporting the parental conflict hypothesis

Analyzing POEs at a biobank scale enables us to move beyond investigating contrasting parental effects on the same trait at a single locus, such as bi-polar effects. While we can explore contrasting effects on the same trait across different loci, a more comprehensive approach leverages the vast range of available phenotypes to examine contrasting parental effects across both traits and loci. This broader perspective allows us to identify overarching patterns and gain deeper insights into the evolutionary dynamics of imprinted genes.

For example, IGF-1 levels exhibited contrasting POEs at two distinct loci. At 6q24.2, we identified a paternal effect decreasing IGF-1 level driven by a variant within the paternally expressed gene *PLAGL1*, which is implicated in transient neonatal diabetes mellitus^33,34^. The role of *PLAGL1* in glucose metabolism and insulin sensitivity^35^ underscores the importance of this locus in regulating IGF-1 levels and metabolic pathways. At 16p13.3, we observed a maternal effect increasing IGF-1 levels driven by a variant that acts as eQTL for two maternally expressed genes, *ZNF597* and *NAA60*, as well as for *TRAP1*, a gene linked to mitochondrial function and metformin use^36^. Together, these findings illustrate how contrasting maternal and paternal effects at different loci collectively shape the genetic architecture of IGF-1 levels, offering valuable insights into the broader evolutionary dynamics underlying imprinted loci.

Similarly, for standing height, we identified contrasting parental effect at two independent POE loci at 11p15.5. We observed a paternal effect of rs576603, and a maternal effect of rs143840904, located 900 kb away. This pattern also exemplifies how parental effects can differ at separate loci for the same trait, reflecting a complex interplay of genetic influences. Notably, such findings align with the parental conflict theory, which posits that paternal alleles are selected to enhance resource extraction from the mother, promoting offspring growth and increasing the father’s genetic success. In this framework, paternally expressed imprinted genes often drive growth and resource acquisition. In contrast, maternally inherited alleles are selected to conserve maternal resources for future reproduction and offspring survival, with maternally expressed genes acting as growth suppressors to counterbalance paternal effects. At the genetic association level, this dynamic is reflected by differential paternal and maternal effects on traits related to resource allocation, often across different loci and in opposing directions, exemplifying bi-polar patterns. This pattern is evident at other loci too, for traits related to resource allocation: at 7q32.2 for SHBG, triglycerides and HDL-cholesterol; at 11p15.5 for T2D, HbA1c, glucose and height, but also for basal metabolic rate, whole body water mass, leg and trunk fat-free masses, and creatinine and cystatine C ; at 15q12 for total protein levels, and finally at 20q13.32 for fat percentage in the arms, legs, trunk, and whole body. These findings underscore the role of imprinted genes in balancing maternal and paternal interests across traits linked to resource allocation, fat storage, and energy usage. Their strong presence among our findings (across multiple loci and traits) may support the conflict hypothesis and likely represent footprints of the evolutionary forces shaping the genetic architecture of complex traits.

### Differential parental contributions to SNP heritability

To examine genome-wide parental contributions to complex traits, as well as those within imprinted regions, we estimated SNP heritability separately for paternal, maternal, and POEs. This analysis revealed notable differences in the contribution of maternal and paternal haplotypes to certain traits, particularly those related to resource allocation, in line with the parental conflict hypothesis. For example, type 2 diabetes and arm fat percentage exhibited higher SNP heritability from maternal alleles. Conversely, traits such as birth weight, glucose levels, and basophil count showed higher heritability from paternal haplotypes. These differences likely reflect distinct parental contributions to biological processes associated with growth, metabolic regulation, and immune function. Interestingly, some traits, such as IGF-1 and triglycerides, exhibited no significant differences between maternal and paternal heritability estimates but showed substantial PofO-specific heritability (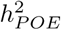).

This suggests that maternal and paternal-specific genetic contributions to these traits are distributed differently across the genome.

### Protein QTLs, a new frontier for POEs

We further leveraged protein quantification data both to validate POEs at the protein level and to assess POEs at established protein QTLs (pQTLs). Although the available protein panel did not cover many of the genes located at our complex trait POE-variant, we identified a significant POE-pQTL for *CPA4* when testing variants initially linked to triglycerides, HDL-C, and SHBG. While this could initially suggest a causal relationship between *CPA4*, the lead variants, and these traits, further analysis revealed a much more significant POE-pQTL located approximately 75 kb away from the POE-variant and in moderate linkage disequilibrium (*r*^2^ *>* 0.05). This finding indicates that the detected POEs on triglycerides, HDL-C, and SHBG are unlikely to be driven by *CPA4*, with the moderate significance observed at the POE variant likely arising from linkage disequilibrium with the true causal variant. Instead, other genes at this locus, such as *KLF14* and *MEST*, are more plausible candidates due to their established eQTL associations with the lead variants. In addition, the POE-variant exhibited a paternal effect on *CPA4* protein level, despite the gene being maternally expressed. This likely reflect influence of paternally expressed genes via shared protein network, such as *MEST* ^37^. While this finding does not conclusively identify the gene responsible for the observed POEs, it allows us to refine our candidate list, excluding *CPA4* and prioritizing other loci for further study.

Beyond testing complex trait POE-variant for POE-pQTLs, we also evaluated PofO specificity at established pQTLs^25^ and identified four significant POE-pQTLs. Two of these were located within known imprinted regions and showed parental effects consistent with the parental expression of the respective genes. For instance, the protein product of the paternally expressed gene *DLK1* was predominantly associated with paternally inherited alleles, though a moderate maternal association was also observed. This biparental influence likely reflects the interconnected nature of imprinted gene networks, where maternally expressed genes can indirectly modulate the expression or activity of paternally expressed genes. For example, the imprinting regulation of *DLK1* is also dependent on two differentially methylated regions (DMRs), including the *MEG3* DMR^38^, a maternally expressed gene. This highlights the complex epigenetic mechanisms that drive *DLK1* expression and its imprinting pattern. In contrast, the protein product of *CPA4* exhibited a strong maternal association, consistent with its preferential maternal expression^39^. Interestingly, a significant - but moderate - paternal association was also observed for *CPA4*, potentially reflecting network-level interactions with paternally expressed genes such as *MEST*, which may modulate *CPA4* protein levels through shared pathways^37^.

The remaining two POE-pQTLs were identified for genes outside known imprinted regions: *ADAM23* and *PER3*, both of which exhibited paternal effects, albeit demonstrating “asymmetric polar” effect. Although *ADAM23* is not currently classified as imprinted, it is located near two paternally expressed genes, *ZDBF2* and *GPR1*, and a recent study reported a paternal bias in its expression^40^, suggesting incomplete imprinting. Similarly, *PER3* has been documented to show paternal-biased expression^41^, consistent with the paternal effect observed in our study. These findings strengthen the case for these genes as candidates for incomplete or context-dependent imprinting and point to broader PofO specific mechanisms that extend beyond imprinting and involve rearing effects.

Interestingly, no significant POE-pQTLs were detected for several other known imprinted genes. This absence may indicate that protein levels do not always reflect RNA expression due to complex feedback mechanisms that could mask detectable PofO associations at the protein level. These results underscore the importance of complementary transcriptomic analyses to capture the full spectrum of POEs. Recent studies focusing on parental-specific eQTLs in parent-offspring trios have successfully validated known imprinted genes and identified novel candidates, emphasizing the value of integrating multi-omics approaches^19^.

### Replication analyses support our novel findings

Lastly, we validated several of our findings in the Estonian Biobank (EstBB), a cohort with an enrichment of familial relatedness, though limited in phenotype overlap with our detected POEs. Among the ten associations replicated in the EstBB, six were novel associations, while four corresponded to previously reported loci or variants in high linkage disequilibrium with known associations. Specifically, we confirmed our newly identified bi-polar effect of rs62471721 on triglycerides, rs4731690 on HDL cholesterol, and rs4264135 on creatinine. Additionally, we also validated our novel bi-polar effects of rs4417225 on glucose levels and rs10838787 on HbA1c. While the latter locus had been previously associated with T2D, its link to glucose-related traits provides further, albeit less novel, insights. Notably, among the replicated associations were bi-polar effects driven by variants acting as eQTLs for distinct genes with contrasting parental expression (for HDL cholesterol and triglycerides). This finding further supports the hypothesis that pleiotropic effects may offer a mechanistic explanation for bi-polar dominant phenomena. Not all POEs identified in the UKBB were replicated in the EstBB, which could reflect differences in statistical power, cohort characteristics, or environmental factors. For instance, while two of the POEs on standing height at 11p15.5 were successfully replicated, the third association at the same locus was not. This discrepancy could arise from cohort-specific imprinting or parental nurturing effects.

Interestingly, a POE not replicated in the Estonian cohort — the maternal effect on hip circumference — was validated in the MoBa cohort, where it showed a significant effect on infant BMI. This effect, which operates in opposite directions during infancy and adulthood, highlights the importance of examining POEs at different stages of life to capture the dynamic nature of genetic imprinting.

### Summary and future directions

This study, along with its replication efforts, provides significant insights into the genetic and evolutionary mechanisms underlying parent-of-origin effects, with several key discoveries highlighted below.

First, we established the largest cohort to date with parent-of-origin information, enabling the most comprehensive analysis of POEs across diverse traits, diseases, and protein levels. By employing a focused analytical approach to reduce the multiple testing burden, we identified POEs that might not have achieved genome-wide significance in a traditional genome-wide scan.

Second, our findings highlight the prevalence of bi-polar POEs, which we discovered to be far more common than previously recognized at imprinted loci. We propose a plausible explanation for these effects in cases where lead variants act as eQTLs for both maternally and paternally expressed genes, such as *KLF14* and *MEST* at the 7q32.2 locus. These findings underscore the intricate regulatory dynamics at imprinted loci and their broader evolutionary significance.

Third, we characterized a novel type of POE, termed “asymmetric polar” effects, where both parental alleles influence a trait in the same direction, but with one allele exerting a much stronger effect. This sheds light on potential mechanisms linked to incomplete imprinting, as exemplified by loci such as *ADAM23* and *PER3*, and offers a new dimension to our understanding of imprinting biology.

Fourth, while not replicated in the Estonian Biobank, we report the first sex-specific POE observed on glucose levels. This finding highlights the potential complexity of sex-dependent imprinting, though we acknowledge that cohort-specific factors, such as differences in sample collection or sex-specific participation patterns, may influence the result^42^.

Fifth, many of our results, both at the single-locus level (e.g., POE-GWAS hits) and the multi-trait level (e.g., cross-trait POE-GWAS analyses and heritability estimates), strongly align with the parental conflict hypothesis. This is exemplified by the prevalence of bi-polar effects on traits related to resource allocation, where maternal and paternal alleles exert opposing influences. Our findings provide robust empirical support for this hypothesis, particularly in traits where opposing parental pressures manifest clearly.

Lastly, our validation efforts in both the Estonian Biobank and the MoBa cohort demonstrate that while some POEs are robustly replicated, others require larger sample sizes to achieve significance, such as the sex-specific POE on glucose. Moving forward, expanding sample sizes through meta-analyses across multiple familial biobanks, such as Finngen^43^ or HUNT^44^, will be critical. Such efforts will be pivotal in uncovering the full genetic architecture of POEs and understanding their implications for complex traits.

## Methods

### UK Biobank genotype processing

We used the UK Biobank Axiom Array data provided in PLINK format^45^ and converted it to VCF format using PLINK v1.90b5^46^. We then used the UK biobank SNPs QC file (UK Biobank resource 1955) to keep only variants used for the official phasing of the original UK Biobank data release^45^, resulting in 670,741 variant sites across the 22 autosomes and 16,601 variant sites on chromosome X. We then used the SHAPEIT5 phase common tool^20^ with default parameters and no filter on allele frequency to perform an initial phasing of autosomes. For chromosome X, we proceed as described in the official phasing report of the UK Biobank whole genome sequencing data, interim release of 200,031 samples^47^. Briefly, we removed pseudoautosomal regions (PAR), identified males as genetically determined, forced reference allele homozygosity at heterozygotes sites in males, and finally we provided the list of male individuals as input in the *–haploid* option of SHAPEIT5. Female individuals were phased as autosomes.

### Close relative clustering

We use pairwise kinship estimates computed using the KING software^48^ to identify related individuals up to the 4*^th^* degree in the UK Biobank cohort. We identified parent-offspring duos and trios as having kinship between 0.1767 and 0.3535, IBS0 below 0.0012 and age difference greater than 15 years^3,45^, resulting in 1,071 trios and 4,136 duos (Supplementary Figures 1A-B). We used Mendel error rate to ensure the accuracy of the identified parent-offspring trios (Supplementary Figure 1C).

We next identified sibling pairs as those with kinship between 0.1767 and 0.3535, IBS0 above 0.0012 and age difference smaller than 15 years^3^, resulting in 22,751 pairs for 41,661 unique individuals (Supplementary Figures 1A-B).

For individuals with more distant relatives (up to 4*^th^*degree), we utilized a clustering approach to segregate relatives by parental sides using the *igraph* package in R, as done previously^3^. Through this method, we identified 274,525 UK Biobank participants whose relatives could be clustered into parental groups based on their relatedness (Supplementary Figure 1D). In our analysis we refer to those close relatives as “surrogate parents”.

We finally identified individuals who have both available parental genomes and inferred surrogate parents, which are utilized for validating our method. This allows us to perform the entire analysis using surrogate parents and excluding parental genomes, effectively simulating the remaining individuals without available parental genomes, and later reintroduce the parental genomes solely for validation and accuracy estimation. In subsequent sections we refer to those as the “validation cohort”. A total of 2,141 individuals met these criteria, resulting in 3,160 target-relative pairs (Supplementary Figure 2A). For each target-relative pair, we used KING pairwise kinship estimates between the target’s relative and the target’s parents to assign the relative to a parental side (Supplementary Figure 2B). This validation cohort was subsequently used to assess the accuracy of our inferences and to derive probabilities for PofO assignments.

### Inter-chromosomal phasing from available parental genomes

For individuals with available parental genomes (1,071 trios and 4,136 duos), we performed inter-chromosomal phasing using the *–pedigree* option of SHAPEIT5^20^. This method requires a three-column pedigree file listing offspring, fathers, and mothers. In cases where one parent is unavailable, the missing parent is indicated by *”NA”*. This procedure results in phased genotype data (i.e, haplotypes). The order of the haplotypes correspond to the order of parents in the pedigree file (here, first haplotypes are paternally inherited; second haplotypes are maternally inherited). Consequently, this method allows direct inference of the PofO of haplotypes from statistical phasing when parental genomes are available. The key advantage of using pedigree-based statistical phasing over traditional Mendelian logic is its ability to infer the PofO of alleles even when both parents are heterozygous. This is accomplished by leveraging haplotype information from the broader population, providing more robust and accurate results, such as done in traditional statistical phasing. In this study, the PofO inferred from parental genomes serves as the ground truth. This ground truth is critical for validating the accuracy of PofO inference derived from surrogate parents in the validation cohort (see subsequent sections).

### Inter-chromosomal phasing from surrogate parents

Identity-by-descent (IBD) mapping is a powerful approach for identifying haplotype segments co-inherited from a common ancestor among pairs of relatives. By analyzing haplotype segments shared IBD with the same set of surrogate parents across the 22 autosomes for a given target individual, we can determine which haplotype segments are inherited from the same parent. This enables the construction of partial parental haplotype sets, which can then be used to perform inter-chromosomal phasing—segregating haplotypes inherited from each parent across the genome. This approach goes beyond traditional phasing methods, which are limited to resolving haplotypes within individual chromosomes (intra-chromosomal phasing).

For 274,525 individuals with identified surrogate parents, we used the THORIN tool^3^ to map haplotype segments shared IBD between the target individuals and their surrogate parents. These IBD-shared segments served as the foundation for both intra- and inter-chromosomal phasing, building on prior methodologies^3^. The key principle here is that haplotypes shared IBD with the same surrogate parents originate from the same parent. Consequently, these haplotypes should consistently appear on the same parental haplotype across autosomes. In practice, we filtered for shared haplotype segments longer than 3 centimorgans (cM), which we incorporated into a scaffold file. This scaffold was then used as input for the SHAPEIT5 phase common tool. This step allowed us to refine and re-estimate haplotypes from genotype data, while simultaneously correcting intra-chromosomal phasing switch errors and performing inter-chromosomal phasing by assigning all haplotypes shared with the same surrogate parents to the same parental haplotype (e.g., first or second) across all 22 autosomes.

This approach was applied to 274,525 individuals from the UK Biobank. Accuracy assessments (Supplementary Figure 3) demonstrated the robustness and reliability of our interchromosomal phasing method in segregating parental haplotype sets with high precision.

### Surrogate parents’ parental side determination

#### Chromosome X

Identity-by-descent (IBD) mapping on chromosome X has been shown as an accurate approach to identify maternal relatives for male target individuals^3^. Here, we build upon this previous approach to probabilistically infer close relatives’ parental side. To do so, we utilized the THORIN tool to map IBD segments between a target individual and its surrogate parents. When a target individual has several surrogate parents from the same family side, we combined them into a single surrogate parent set, which allow the THORIN tool to merge overlapping IBD segment from different relatives within the same set. We then retained only the largest IBD segment (size computed in Morgans) between the target and each surrogate parent set. Unlike the previous approach that only determine maternal relationships using a strict threshold in the chromosome X IBD^3^, we probabilistically determined the surrogate parents’ parental side. We used the validation cohort to derive probabilities of a surrogate parents set being on the paternal or maternal side based on the length of the chr X IBD haplotype segment shared with the target individual. This method effectively maximizes the number of surrogate parents with determined parental status.

Let *l* be the length in cM of the longest haplotype segment shared between the chrX of a focal target and that of its surrogate parent(s). In addition, we define *l_i_*as the length of chrX haplotype sharing of an individual *i* with its surrogate parents, in the training set. Let *I_pat_* refer to the set of *N_pat_* target individuals with paternal relatives and *I_mat_*to those *N_mat_* targets with maternal relatives in the training set. We derive the probability of the surrogate parent of a target individual (in the test set), sharing *l* cM chrX with its surrogate parent, being on the paternal and maternal side as follows:

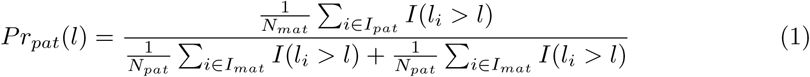

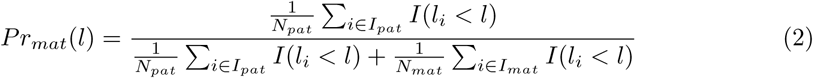

where

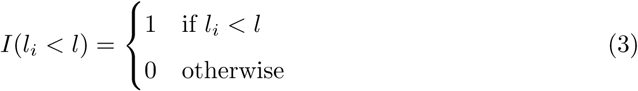

We initially applied this approach on the male individuals subset of the validation cohort (N=857). We found that all surrogate fathers share IBD haplotype segments smaller than 11.3 cM, and 99% share segments smaller than 3 cM (Supplementary Figure 4A). We then evaluated the accuracy of our probabilistic assignment. To do so, we proceeded with a “Leave One Sample Out” approach: we removed a given individual from the training set and predicted its paternal and maternal sides by deriving probabilities using the *N* − 1 individuals of training set (Supplementary Figure 4B). We repeated this for the *N* individuals of the training set. To compute accuracy at a given maternal probability *p*, we defined true positives (TP) as the number of surrogate mothers with probability *> p* and false positive (FP) as the number of surrogate fathers with probability *> p*. We then computed accuracy as *TP/*(*TP* + *FP*) (Supplementary Figure 4C).

Applying this method to the UK Biobank male individuals with available surrogate parents and unknown parental assignment, we were able to probabilistically assign parental sides to surrogate parents for a total of 115,027 male individuals. Notably, 34% of these individuals (N=39,100) achieved the highest possible parental assignment probability, indicating a robust and reliable classification (Supplementary Figure 4D).

#### Mitochondrial DNA

Mitochondrial DNA (mtDNA), which is exclusively inherited from the mother, serves as a valuable tool for identifying maternal relatives. However, the UK Biobank Axiom array data proved inadequate for this purpose due to the limited availability of genotyped variants (N=265). To overcome this limitation, we used the whole-genome sequencing GraphTyper cram file available on the UK Biobank Research and Analysis Platform (RAP) for 500,000 individuals. We called variants from the mtDNA whole-genome sequencing (WGS) cram files for 274,525 UK Biobank individuals (those with inter-chromosomal phasing) and their surrogate parents using the MitoHPC software^49^ with default parameters.

We then aimed at determining a surrogate parent’s parental side using genetic similarities on the mtDNA. However, typical IBD mapping software are derived from the Li & Stephens Hidden Markov Model, which excels at modeling the human recombinant genomes architecture by accounting for mutation and recombination between haplotypes. Due to mtDNA’s lack of recombination and its higher mutation rate compared to autosomes, these software proved unsuitable. As a result, we adopted an alternative approach to evaluate non-recombinant DNA sharing between pairs of individuals. This approach is inspired from the Jaccard index and termed here Minor Variant Sharing (MVS). It involves evaluating the proportion of shared minor alleles between relative pairs. This allows us to overcome the limitations of traditional IBD mapping software for mtDNA by basing our analysis on IBS (Identity-By-State) rather than IBD.

Let *V_t_* be the set of variants in the target individual. Similarly, let *V_r_* be the set of variant in the target’s relative. Let 0 be the value of a major allele, and 1 be the value of a minor allele. Thus, we compute mtDNA MVS as:

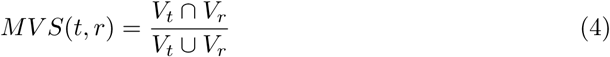

Using the validation cohort, we found that maternal relatives exhibit a higher average MVS compared to paternal relatives (Supplementary Figures 5-7A). The effectiveness of this metric depends on the degree of relatedness and is particularly precise for 2*^nd^*degree relatives. For these relatives, MVS values strongly correlate with the probability of a maternal relationships. However, for more distant relatives (due to the possible presence of an intermediary male relative in the genetic lineage disrupting mtDNA inheritance from the maternal side), low MVS values do not translate to paternal relationship.

To predict the parental side of relatives from the mtDNA MVS, we adopted a perfectly analogous procedure to the one outlined for chrX sharing whereby we replaced chrX IBD segment length with the mtDNA MVS value. To evaluate the accuracy of this approach, we stratified target samples by their degree of relatedness to their closest relatives in the cohort (Supplementary figures 5-7C).

This methodology was applied to predict relative parental side for 19,022 2*^nd^* degree, 114,965 3*^rd^* degree, and 312,447 4*^th^* degree target-relative pairs (Supplementary Figures 5D-7D). For targets with multiple relatives of the same degree, we retained only the relative with the highest predicted accuracy, resulting in 17,625 2*^nd^* degree, 95,151 3*^rd^* degree, and 204,924 4*^th^* degree target-relative pairs. Notably, 69,580 of these pairs were assigned to a parental side with a probability greater than 0.99 (Supplementary Figure 8A). Additionally, parental side assignments were supported by multiple relatives for 9,948 individuals (Supplementary Figure 8B). Together, this indicates a reliable parental assignment from mtDNA MVS predictions.

#### Chromosome Y

Chromosome Y sharing between male relatives can indicate paternal lineage. Due to chromosome Y lack of recombination outside PAR regions, we did not employ IBD mapping. Instead, we calculated chromosome Y MVS, similar to our approach with mtDNA. Similarly, the effectiveness of this method depends heavily on the degree of relatedness, as an intermediary female in the genetic lineage interrupts chromosome Y transmission. We used chromosome Y whole-genome sequencing DRAGEN pVCFs files available on the UK Biobank RAP to compute MVS for male target-male relative pairs as described above for mtDNA. We stratified chromosome Y variants by variant type (SNV and Indels) and by Minor Allele Frequency (MAF). Since this can be done only for male target-relative pairs, we are practically limited to using only 3,907 2*^nd^*, 24,342 3*^rd^* and 66,124 4*^th^* degree relative pairs (72,730 unique total target individuals). To explore the potential of this approach, we analyzed 543 male target-male relative pairs from the validation cohort. We found that rare MVS on chromosome Y (MAF *<*1%) is a reliable predictor of both paternal and maternal sides for all 2*^nd^* degree relatives, a good predictor of paternal side for approximately 13% of 3*^rd^* degree relatives, but is inaccurate for 4*^th^* degree relatives (Supplementary Figure 9). However, since 2*^nd^* degree relatives can already be assigned parental sides with high accuracy using mtDNA, the practical utility of Chromosome Y MVS lies primarily in resolving ambiguous PofO assignments among 3*^rd^* degree relatives. Among 3*^rd^* degree relatives, 11,525 individuals had PofO probabilities below 0.95 when combining Chromosome X and mtDNA predictions. Based on the prediction rate of 13% for Chromosome Y MVS, this method could add approximately 1,500 individuals with improved parental assignments. While this novel approach effectively determines paternal lineage, its limited contribution to sample size—relative to the computational effort and cost of implementing it at scale on the UK Biobank RAP—led us to conclude that a broader application of this method was not justified. Despite its proven accuracy, we opted not to implement Chromosome Y MVS for all 3*^rd^* degree target-relative pairs.

### Parent-of-Origin determination

#### From close relatives

We integrated inter-chromosomal phasing with inferred parental side information from relatives to determine the PofO for each parental haplotype set. Specifically, a haplotype set was classified as maternally inherited if its haplotypes shared IBD segments with maternal relatives, and paternally inherited if they shared IBD segments with paternal relatives. When the assignment was possible for only one of the two haplotypes in a target individual, we assumed the absence of uniparental disomy and inferred the parental origin of the second haplotype by exclusion.

#### From crossovers to PoO inference in siblings

By analyzing IBD segments shared by siblings, we can infer crossover (CO) locations in parental haplotypes. Together with sex-specific genetic recombination maps^7^, Qiao *et al.*^8^ have demonstrated that we can estimate the likelihood of a set of COs originating from either the mother or father, allowing us to determine the PofO of the haplotype carrying this set of COs. This method is reliable exclusively for sibling pairs, as they guarantee that the detected CO events occurred in the parents. With more distant relatives, the uncertainty increases significantly. For instance, CO events identified between second-degree relatives could have occurred in the parents or grandparents, resulting in a mix of male-specific and female-specific CO events on the same haplotype.

We identified haplotypes shared IBD between sibling pairs using the THORIN tool. We kept only haplotype segments larger than 3cM. We used IBD segment breakends as CO positions. However, we are very unlikely to pinpoint the exact CO positions using this approach. First, because we are restricted to genotyped markers. Second, due to the nature of the Li & Stephens Hidden Markov Model used to map IBD segments in the THORIN tool which does not transition at homozygous sites - we identify COs at heterozygotes only. Therefore, we used a 1,000 base pair (bp) window around each CO position to increase chances of including the true position.

We then used recombination maps to extrapolate the probability of a CO occurring within this window based on the genetic distance in Morgans between the start and end positions of the window. We repeated this process for both the male-specific and the female-specific recombination maps for all the CO events identified on the same haplotype.

Let *D_f_* (*p*) be the distance between the CO position *p* and the nearest female-specific recombination map position in Morgan and *D_m_*(*p*) the distance to the nearest male-specific recombination map position. To determine the most likely parent giving rise to an observed CO at position *p*, we can compute the difference in Morgan between male and female recombination probability:

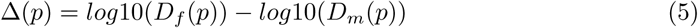

Assuming perfect intra-chromosomal phasing, we can deduce that all CO identified on a given haplotype are inherited from the same parent. We can therefore aggregate Morgan differences for the *n*_0_ and *n*_1_ CO positions of haplotype 0 and haplotype 1, respectively, and compute a global score per chromosome using both haplotypes as:

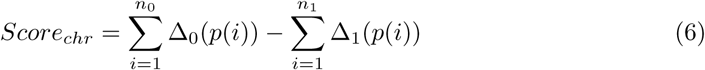

This approach can be sensitive to misidentified crossovers, which may arise from various sources, such as inaccuracies in IBD segment boundaries due to errors in the IBD mapper, genotyping errors, or consanguinity. These errors can inflate or deflate the global score. To mitigate this and remove confounders, we compared the observed number of CO *N_obs_*(*CO*) events to the expected number *N_exp_*(*CO*). *N_exp_*(*CO*) depends on the chromosome length in Morgan *l_M_*. Since we aggregate crossovers across both siblings first and second haplotypes, we can observe COs that occurred on four haplotypes, and thus *N_exp_*(*CO*) = 4*l_M_*. However, CO positions shared between siblings are counted only once, therefore, *N_exp_*(*CO*) represents the theoretical maxima, and the number COs that is possible to identify is within the range [0, 4*l_M_*], where 0 corresponds to siblings in IBD2 or IBD0 for the entire chromosome. In practice, since artifacts from the IBD mapper can inflate this number and go beyond 4*l_M_*, we evaluated the distribution of observed CO for each chromosome. Individuals strongly deviating from this distribution (more than 10 times the interquartile range) are removed. The accuracy of determining the PofO of haplotypes depends on the number of COs analyzed, which varies with chromosome length (in Morgans) (Supplementary Figure 10A). Using intra-chromosomal phasing, we could only determine the PofO for COs within individual chromosomes by computing *Score_chr_*, requiring separate calculations for each chromosome. In contrast, inter-chromosomal phasing enabled us to group COs occurring across the entire genome on the same parental haplotype set, allowing us to infer the PofO for the entire set at once. This enabled the computation of a genome-wide score, thereby increasing the available genomic length and number of COs (Supplementary Figure 10B):

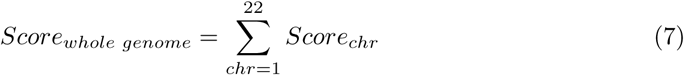

We initially assigned the PofO of haplotype using this score as follows: a negative score means that the first haplotype of the individual is paternally inherited, and a positive score means that it is maternally inherited. We evaluated the efficiency of our approach using a subset of the validation cohort that included 88 individuals with at least one sibling and available inter-chromosomal phasing from close relatives. Since inter-chromosomal phasing might not be available for all chromosomes, we simulated varying conditions by altering the number of chromosomes analyzed per individual, resulting in different numbers of inferred COs depending on the genomic length analyzed (Supplementary Figure 10B). We found that inter-chromosomal phasing significantly increases accuracy compared to single chromosome analysis (Supplementary Figures 10C-E). Moreover, the PofO of COs could be assigned for 96.4% of individuals using inter-chromosomal phasing, versus only 51.3% with intrachromosomal phasing (Supplementary Figure 10F).

To avoid relying on strict threshold, we utilized the validation cohort to derive probabilistic determinations of haplotypes’ parental origin using inferred COs from inter-chromosomally phased data. Let us consider a score *S_t_* for the target individual *t* that has been computed using a specific number of inter-chromosomally phased chromosomes. Taken altogether, these chromosomes have a genomic length in Morgan *l_t_*. Since the number of observed COs *N_obs_*(*CO*) depends on *l_t_* and that *S_t_* depends on *N_obs_*(*CO*), we define *I_pat_*(*t*) as the set of individuals of the validation cohort for which *H*_0_ is paternally inherited and for which *l* is within the window (*l_t_* −3*, l_t_* +3), and *I_mat_*(*t*) as the set of individuals of the validation cohort for which *H*_0_ is maternally inherited and for which *l* is within the window (*l_t_*− 3*, l_t_* + 3). The cardinality of these sets are denoted by *N_pat_*(*t*) and *N_mat_*(*t*), respectively. This allows us to compare our obtained score to a subset of the validation cohort that had a similar available genomic length as *l_t_*.

Since we assigned negative scores to paternal side and positive scores to maternal side, we can then define the probability of paternal origin of a genome-wide haplotype of target sample *t* as follows

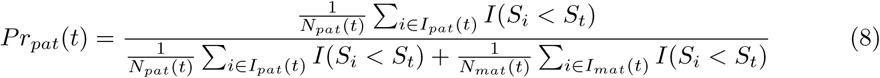

We evaluated the accuracy of this approach using the validation cohort. We simulated varying conditions by altering the number of chromosomes analyzed per individual, ranging from only 1 chromosome used, to the full set of inter-chromosomally phased chromosomes for each individual. We observed better accuracy for scores strongly deviating from zero (Supplementary Figure 11A). In addition, we observed that larger available genomic lengths (i.e, more chromosomes) resulted in less errors, and that no errors were detected when using more than 10 Morgan (Supplementary Figure 11B). Notably, when using the full set of available inter-chromosomally phased chromosomes per individual in the validation cohort (i.e the most realistic scenario), we achieved 100% accuracy (Supplementary Figures 11A-B).

We applied this approach on 26,635 individuals with available inter-chromosomal phasing and at least one sibling. The specific configurations of our sample set (genomic length and number of COs) matched the highest accuracy (Supplementary Figures 11C,D).

Since inter-chromosomal phasing is not available for all siblings, we additionally use 14,695 individuals without inter-chromosomal phasing available and at least one sibling. We applied the single-chromosome approach using strict threshold at scores of −2 and 2 to determine the PofO of each target haplotypes separately, and assessed the different parameters of this approach (Supplementary Figure 12).

#### Accuracy of combined parent-of-origin predictors

A given individual can have several predictors of its haplotype PofO: chromosome X IBD, mtDNA MVS from different relatives, and sib-score. For each individual, we kept the predictor with the probability yielding the highest accuracy at determining paternal or maternal side. As a result, we could unambiguously determine the relatives’ parental side for 99.1% of the target individuals (N=267,831). For ambiguous cases (N=2,491), we then prioritize predictors in the order indicated by overall accuracy of the predictor: chromosome X, sibscore, MVS for second-degree relatives. We obtained 209 individuals with parental sides undetermined because of conflicting mtDNA-based PoO predictions from different (third- and fourth-degree) relatives in the same surrogate parent cluster. In addition, we also had 14,596 individuals with PofO inferred from single-chromosome sibling-score. By removing the validation cohort and combining with individuals having PofO inferred from available parental genomes, we assembled a final set of 286,666 individuals. We finally compared our PofO determination to the one obtained from parental genomes at each heterozygous site of the validation cohort, revealing an accuracy of 97.94% (Supplementary Figure 13).

### Parental haplotypes imputation and encoding

To infer the PofO for untyped alleles, we used haploid imputation as done previously^3^ to separately impute each parental haplotype using the *–out-ap-field* option of IMPUTE5 v1.2.1^50^ and the Haplotype Reference Consortium as a reference panel. This provides an AP field indicating imputed paternal and maternal allele dosages. We then kept only variants with an INFO score greater than 0.8 and Minor Allele Frequency (MAF) greater than 1%. We encoded parental haplotype in separate files. For each target and at each variant, we weighted parental allele imputed dosage by the parental assignment probability. Let us consider an imputed variant *v* for a target individual *t* with haploid imputed dosages *AP_mat_*(*t, v*) and *AP_pat_*(*t, v*) for maternal and paternal haplotype, respectively. Let us also consider the parental assignment probability *p_t_*for the target *t* given by combined predictors. We re-encoded our data as:

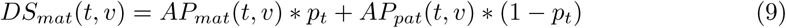

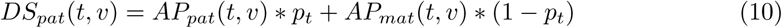

From this, we adapted a haploid genotype posterior probability field *GP* (probability triplets for the three possible genotypes) as:

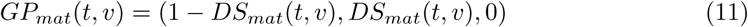

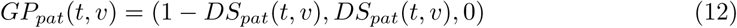

Finally, we also combined maternal and paternal alleles at heterozygous only to directly test for differential effect (all homozygous sites are encoded as missing):

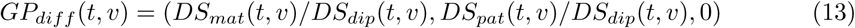

where

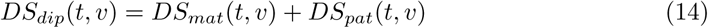

### Identification of Parent-of-origin effects

Parent-of-origin effects is a generic name that characterizes many different types of genetic effects whose effect depends on the parental origin of the assessed allele, the most known probably being strict paternal and maternal effects. However, many other type of POE exists (Supplementary Figure 15). To increase our likelihood of detecting POE effect, we use the differential scan to directly assess the differential effect between paternal and maternal alleles among heterozygous individuals. To then infer the direction of the effect, we use a paternal-specific and maternal-specific GWAS scan, which separately evaluate the effect of each parental haplotype. To perform GWAS analysis, we used Regenie v3.2.9^51^. We performed the model fitting only on the genotyped variants as recommended (i.e regenie step1). We then tested paternal, maternal and differential haplotypes (i.e *GP_pat_*, *GP_mat_* and *GP_diff_*) separately (i.e regenie step2). We denote the p-values resulting from these GWAS *P_P_*, *P_M_* and *P_D_* for paternal, maternal and differential GWAS, respectively. We used the first 20 Principal Components (PCs), age and sex as covariates. In addition, we restricted our association tests to individuals who self-identified as “White British” and have a similar genetic ancestry determined by principal components analysis (UK Biobank field 22006), and we restricted the differential scan to variants with imputed PofO dosages greater than 0.99, as done previously^3^.

Given the complexity and diverse nature of POE effects, we employed a multi-faceted approach to identify novel putative associations. Beside a traditional genome-wide scan, we leveraged biological knowledge by focusing on known imprinted regions, which are more likely to exhibit POE. Second, recognizing that POE effects can also arise outside of these known regions, we expanded our analysis to perform a GWAS scan focusing on regions exhibiting additive association with the examined trait. For these analyses, we selected a total of 59 phenotypes, mostly from blood biomarkers and morphological traits. This comprehensive approach allowed us to maximize the potential for discovering novel POE signals across a broad spectrum of traits. To account for the number of traits tested, we computed phenotypic correlation between each pair of traits. We then used the function *simpleM* from the R package *hscovar* to compute the effective number of traits tested, which resulted in *N_eff_* (*traits*) = 48. All associations were pruned using genetic distance smaller than 500kb and LD*<*0.01.

To identify POE genome-wide, we used a stringent POE significance threshold (i.e, differential P-value *P_D_*) of 5 × 10^−8^*/*48 = 1 × 10^−9^ to correct for the number of independent tests performed across our 59 selected traits.

To identify POE within additive regions, we first selected significant additive associations (p-value *<* 5 × 10^−8^*/*48 = 1.04 × 10^−9^), which we then tested for POE using the differential GWAS. We corrected our POE significance threshold for the number of identified additive association such that the differential p-value *P_D_* threshold = 0.05*/N_a_*, where *N_a_* is the number of independent additive associations.

To identify POE within imprinted regions, we limited our analysis to variants within a 500kb window around known imprinted genes, as these regions are biologically predisposed to display differential effects based on parental origin. This targeted strategy enabled us to leverage prior biological knowledge to enhance the likelihood of identifying true POE effects. In practice, we use imprinted regions coordinates published previously^5^. To ensure statistical rigor while maintaining sensitivity, we adjusted the POE significance threshold by accounting for the LD structure within these regions. After calculating the number of effective independent tests, we applied a significance threshold of *P_D_ <* 0.05*/*16, 574 = 3.01 × 10^−6^, where 16,574 is the number of independent tests performed within imprinted regions. Additionally, we verified whether the identified associations would pass the more stringent threshold of 6.27 × 10^−8^, which corresponds to 3.01 × 10^−6^*/*48, where 48 is the effective number of traits tested.

### Conditional parent-of-origin effects analyses

To explore the presence of secondary, independent POEs near primary loci, we performed conditional analyses using Regenie^51^. This involved including the lead LD-pruned variant as a covariate in the model to account for its effect, to obtain a conditional differential P-value *P_Dc_* for the remaining variants. By conditioning on the primary variant, we aimed to identify additional POEs that might otherwise remain undetected due to LD or proximity filtering. This approach ensures that any secondary signals are independent of the primary association, providing a clearer understanding of the genetic architecture and potential multiple regulatory mechanisms within a locus.

### Replication of known parent-of-origin associations

We used publicly available summary data for height studies directly reporting POEs or reporting separate paternal and maternal GWAS estimates^3,4,5,9,10,11,12,13^. When differential GWAS coefficients were not directly available, we computed a differential scan similar to the one performed in our study:

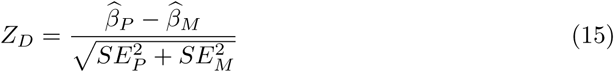

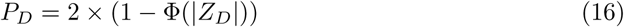

where Φ is the cumulative distribution function of the standard normal distribution and |*Z_D_*| is the absolute value of the z-score.

### Sex-specific differences in Parent-of-Origin effects

We performed a differential GWAS scan separately for males and females to obtain sex-specific coefficients. We then assessed significant differences of effects using a z-score approach:

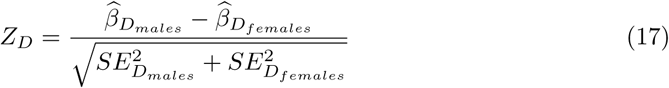

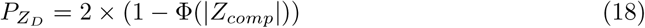

### SNPs heritability

We used the software LD Score Regression^52^ to compute SNPs heritability (*h*^2^) from our GWAS summary data. We used the publicly available LD scores from 1000 Genome Project (KGP). We munged GWAS summary data and estimated heritability as recommended by the authors of the software and with default parameters. For *h*^2^ computed on subsets of data (i.e within- and outside imprinted regions), we weighted the estimates by the proportion of variants included in the data subset. To assess difference between heritability estimates, we combined estimates into a z-score :

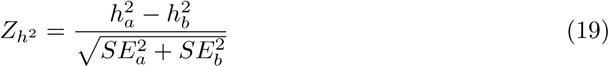

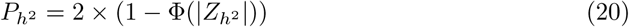

where *a* and *b* are (i) paternal and maternal estimates (Figures 6A,B), or (ii) differential estimates within imprinted regions and outside imprinted regions (Figure 6C).

### Replication analyses in the Norwegian Mother, Father and Child Cohort Study

The Norwegian Mother, Father, and Child Cohort Study (MoBa)^21,22^ is a prospective cohort study that recruited pregnant women in Norway between 1999 and 2008. The study enrolled approximately 114,500 children, 95,200 mothers, and 75,000 fathers from 50 hospitals across Norway. Anthropometric measurements of children were collected at hospitals at birth and during routine visits at 6 weeks, 3 months, 6 months, 8 months, 1 year, 1.5 years, 2 years, 3 years, 5 years, 7 and 8 years. Parents transcribed these measurements into questionnaires^22^. For the analysis of POEs, the parental origin of alleles was inferred using parental genomes. Parent-offspring trios were excluded if they met any of the following criteria: stillborn, deceased, twins, missing data in the Norwegian Medical Birth Registry, missing anthropometric measurements at birth, pregnancies where the mother did not respond to the first questionnaire, or lastly missing parental DNA samples. Genotyping was performed on multiple sets of trios randomly selected from the study biobank, for a total of 45,402 offsprings with both parents genotyped and at least one BMI measurements.

We estimated POEs on infancy and childhood BMI at eleven time points (at 6 weeks, 3, 6 and 8 months, and 1, 1.5, 2, 3, 5, 7 and 8 years of age, Supplementary Table 8) using REGENIE and including as covariates genotyping batch effects for the child, mother, and father, the first 10 principal components, and the child’s sex. Similarly to the UK Biobank and Estonian Biobank cohort, we tested parental alleles in the second step. We then estimated the differential coefficients by comparing paternal *vs* maternal coefficients using a Z-score approach.

We restricted this analysis to 6 loci found to be associated with obesity-related traits in the UK Biobank cohort. Since BMI time points are not uniformly distributed and may contain redundant information, we estimated the number of effective traits tested to apply appropriate multiple testing correction. This resulted in 5 independent measurements, setting our POE significant threshold at 0.05*/*(6 × 5) = 1.67 × 10^−3^.

### Estonian Biobank genotype processing

We used the Quality Controlled (QC) genotype data genome built GRCh38 in Variant Call Format (vcf) provided by the Estonian Biobank team^26^. Similar to when processing the UKBB data, we used SHAPEIT5^20^ to phase autosomes and chromosome X, and KING^48^ to estimate relatedness. To impute the EstBB genotype data, we use the Estonian Biobank WGS reference panel, including 2,695 individuals. We performed phasing of the reference panel using the two steps process of SHAPEIT5^20^. We used the resulting reference panel to impute the data SNP array data using IMPUTE5^50^. For all the remaining analysis, we proceed as for the UK Biobank. To infer the PofO of haplotypes for individuals without available parental genomes, we used only IBD sharing on chromosome X and sibling score, since chromosome Y WGS and mtDNA WGS were not available to compute MVS. To perform association analysis, we focused on traits for which we had at least 20,000 individuals in the Estonian Biobank. This resulted in 10 traits, namely standing height, glucose, type 2 diabetes, glycated haemoglobin, HDL cholesterol, triglycerides, hip circumference, creatinine, cystatin C and platelet count. We derived phenotypes from electronic health records (EHR), unless NMR metabolomics measurements were available^53^, which were available in greater sample size compared to blood biomarkers (for glucose, triglycerides, HDL cholesterol and creatinine).

## Data availability

The summary data will be publicly accessible for download upon publication through our database (http://poedb.dcsr.unil.ch/, currently hosting data from our previous study). Prior to publication, data can be obtained by contacting the corresponding authors. The UK Biobank genetic data are available under restricted access. Access can be obtained by application via the UK Biobank Access Management System (https://www.ukbiob ank.ac.uk/enable-your-research/apply-for-access). The Estonian Biobank data is also available under restricted access. The access to the Estonian Biobank data must be approved by the Scientific Advisory Committee of the Estonian Biobank and by the Estonian Committee on Bioethics and Human Research. More details are available at https://genomics.ut.ee/en/content/estonian-biobank#dataaccess. Data produced as part of this study (i.e inter-chromosomally phased data and PofO information) will be returned to their respective biobanks. Access will be granted upon publication to approved researchers.

## Code availability

The THORIN IBD mapping software version 1.2 is available under MIT license at https://github.com/RJHFMSTR/THORIN. Detailed documentation, tutorials and pipeline to infer the parent-of-origin of alleles are available at https://rjhfmstr.github.io/THORIN/. All other custom codes used as part of this study are provided within the parent-of-origin inference pipeline.

## Acknowledgments

We thank all biobanks participants for sharing their data. The Estonian Biobank data analysis was carried out in part in the High-Performance Computing Center of University of Tartu, Estonia. The UK Biobank data analysis was carried out in part in the High-Performance Computing Center of University of Lausanne, Switzerland. Metabolomics data of the Estonian Biobank has been generated by as part of a collaboration between Nightingale Health Plc and Institute of Genomics, University of Tartu. The development of quality-controlled dataset was supported by the Estonian Research Council grant no PRG1291. This research has been conducted using the UK Biobank Application Number 16389 and 66995, and funded by the Swiss National Science Foundation (SNSF) project grant SNSF 310030-189147. S.J was supported by grants Helse Vest’s Open Research Grant (grants #912250 and F-12144), the Novo Nordisk Foundation (grant NNF19OC0057445) and the Research Council of Norway (grant #315599). M.V was supported by the Research Council of Norway (grant #301178).

## Ethics statement

The activities of the EstBB are regulated by the Human Genes Research Act, which was adopted in 2000 specifically for the operations of the EstBB. Individual level data analysis in the EstBB was carried out under ethical approval 1.1-12/295 from the Estonian Committee on Bioethics and Human Research (Estonian Ministry of Social Affairs), using data according to release application nr T38 from the Estonian Biobank.

The Norwegian Mother, Father and Child Cohort Study (MoBa) informed consent was obtained from all study participants. The administrative board of MoBa led by the Norwegian Institute of Public Health approved the study protocol. The establishment of MoBa and initial data collection was based on a license from the Norwegian Data Protection Agency and approval from The Regional Committee for Medical Research Ethics. The MoBa cohort is currently regulated by the Norwegian Health Registry Act. The study was approved by The Regional Committee for Medical Research Ethics (#2012/67).

## Authors information

### Authors contributions

R.J.H. and Z.K. designed the study and wrote the manuscript. R.J.H. performed all experiments. O.D. contributed to statistical phasing and sex chromosomes integration. S.R. contributed to imputation experiments. T.C. contributed to the implementation of the sibling model. A.V.D.G. contributed to optimizing haplotype data encoding. L.M., R.M. and F.D.P contributed to the analyses in the Estonian Biobank cohort. The Estonian Biobank research team performed data collection, genotyping and QC of the Estonian Biobank genetic data. J.K. and N.T. developed quality control criteria for the Estonian Biobank metabolite data. R.K., M.V., and S.J. contributed to the analyses in the MoBa cohort. All authors reviewed the final manuscript. The project has been supervised by Z.K.

### Corresponding authors

Correspondence to Robin J. Hofmeister and Zoltán Kutalik.

### Additional authors affiliations

#### Estonian Biobank research team

Andres Metspalu^1^, Lili Milani^1^, Tõnu Esko^1^, Reedik Mägi^1^, Mari Nelis^1^ and Georgi Hudjashov^1^

^1^Estonian Genome Centre, Institute of Genomics, University of Tartu

## Supplementary Figures

**Supplementary Fig. 1.**
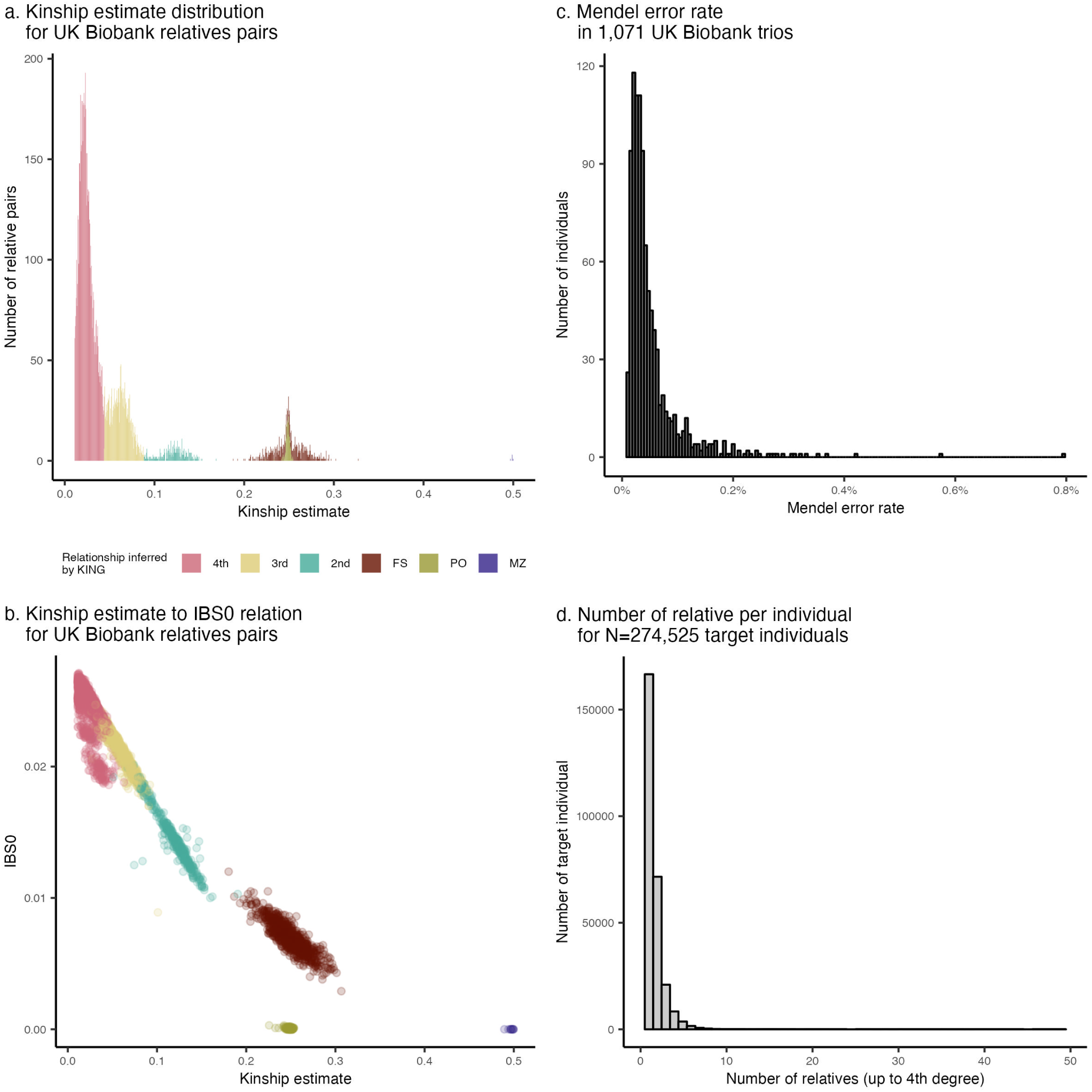
Relatedness inference and close relative clustering in the UK Biobank. **a**) Kinship estimates distribution and **b**) Kinship estimates versus IBS0 estimates across all UK Biobank monozygotic twins (MZ), first siblings (FS), parent-offspring (PO), second-, third-, and fourth-degree relative pairs. **c**) Mendel error rate distribution across 1,071 UK Biobank parent-offspring trios. None of the trios exhibited a high error rate that would necessitate exclusion. **d**) Distribution of the number of close relatives (up to the fourth degree) per individual across 274,252 UK Biobank individuals with available surrogate parent cluster(s). We found on average 1.65 relatives per individual.

**Supplementary Fig. 2.**
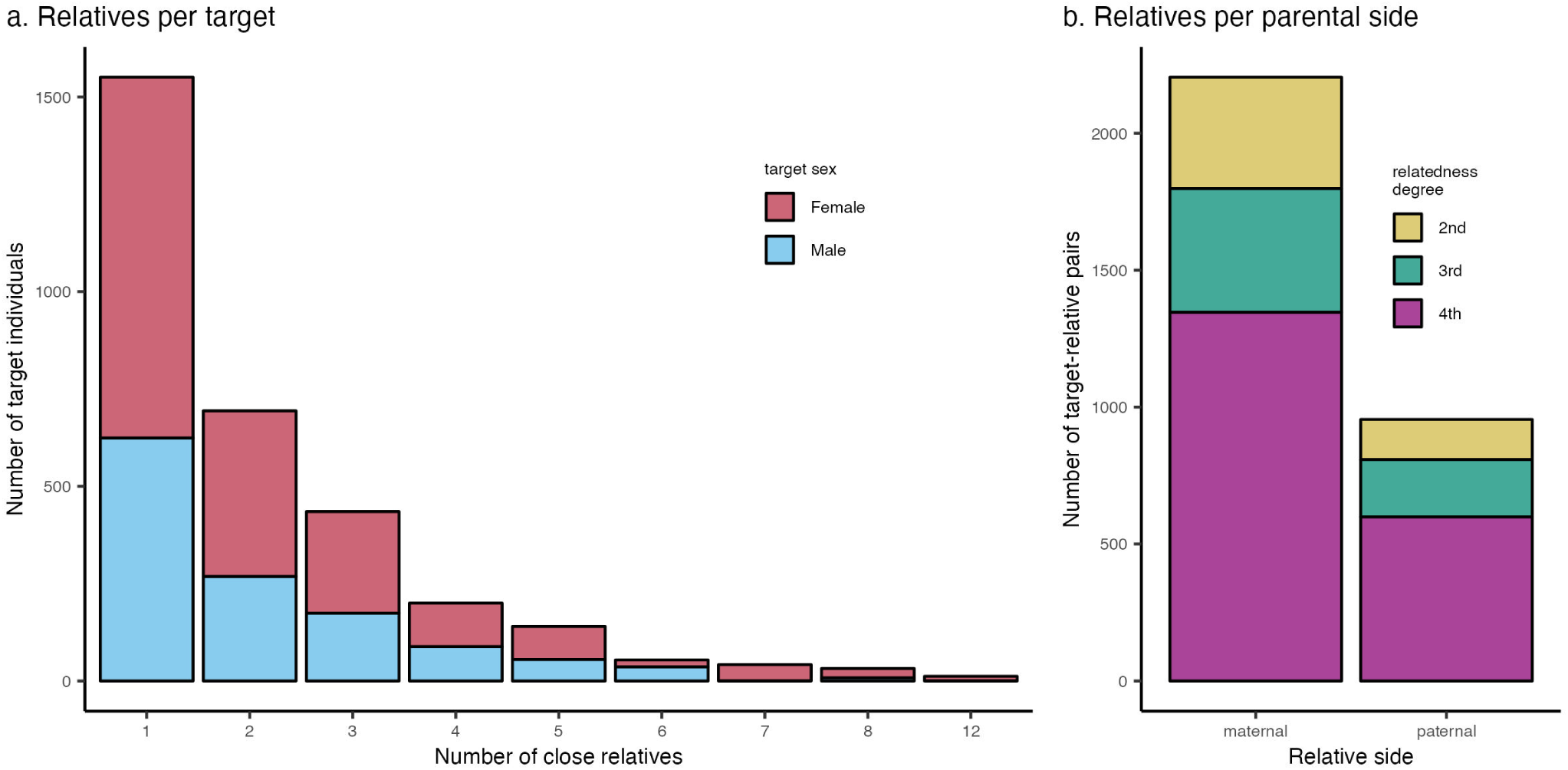
Validation cohort structure. **a**) Distribution of the number of close relatives (up to the fourth degree) per individual across 2,141 UK Biobank individuals with both available surrogate parent cluster(s) and available parental genomes. Most target individuals are females (60.3%) and have a single surrogate parent (49%). **b**) Distribution of surrogate mother and surrogate father stratified by relatedness degree for 3,160 target-relative pairs. Approximately 70% of surrogate parents in the validation cohort are on the maternal side (i.e., surrogate mothers), and the majority are fourth-degree relatives pairs (61.5%).

**Supplementary Fig. 3.**
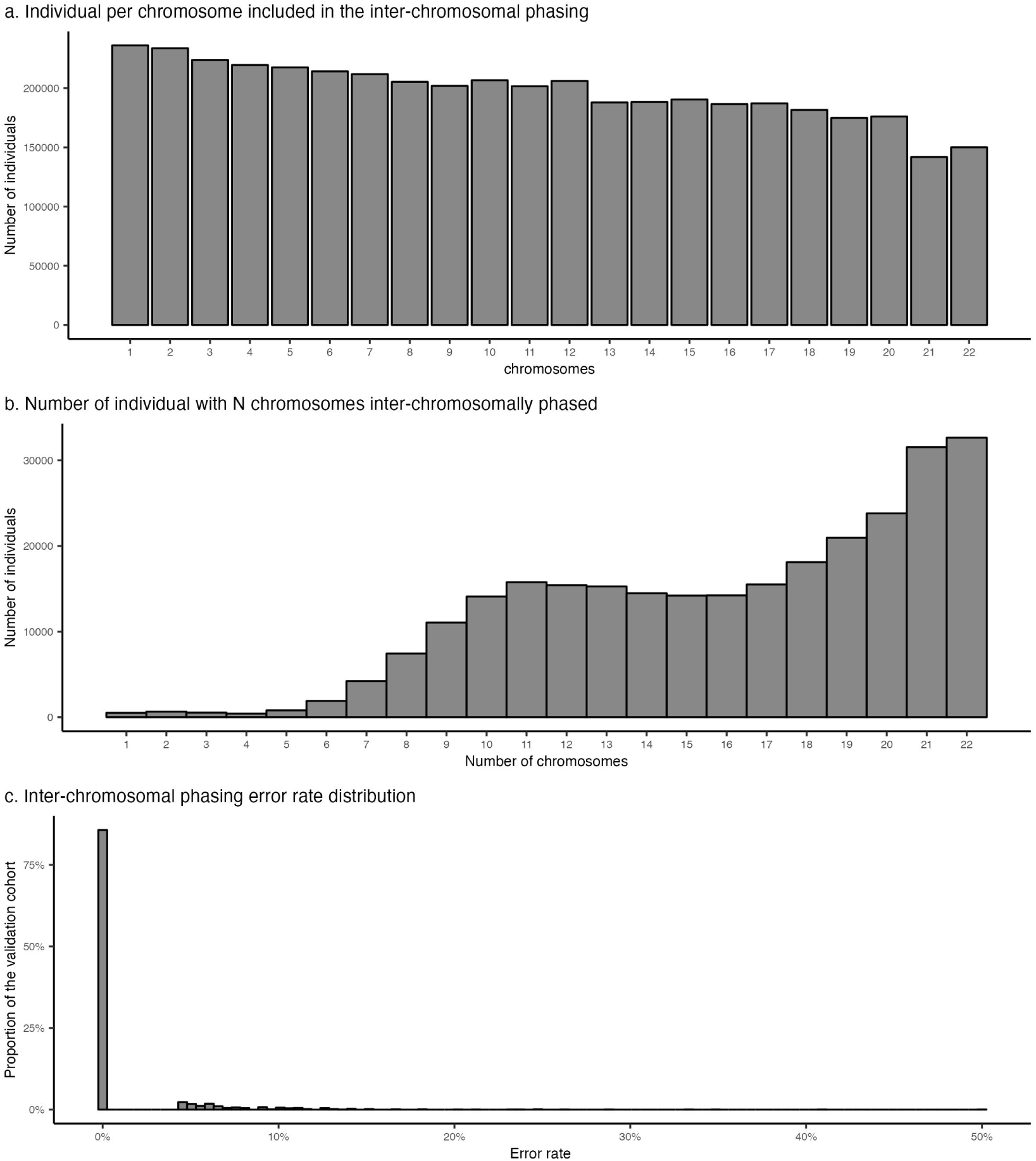
Inter-chromosomal phasing from close relative groups. **a**) Distribution of the number of individuals with a given chromosome included in the inter-chromosomal phasing. The sample sizes vary across chromosomes, as inter-chromosomal phasing requires at least one IBD segment shared with a surrogate parent on the chromosome. Larger chromosomes are more frequently included, likely due to a higher likelihood of containing at least one IBD segment. **b**) Distribution of the number of individuals with N chromosomes (x-axis) successfully included in the inter-chromosomal phasing. Only 12% of individuals (32,651) had all 22 autosomes phased interchromosomally. On average, individuals had 16 chromosomes included in inter-chromosomal phasing. **c**) Distribution of inter-chromosomal phasing error rates across individuals in the validation cohort. Error rates were calculated by comparing phasing results based on surrogate parents to the ground truth obtained from available parental genomes. Most individuals (85.6%) were perfectly phased, with an average error rate of only 1.06% for flipped haplotypes, underscoring the robustness and reliability of the inter-chromosomal phasing method in accurately resolving parental haplotype sets.

**Supplementary Fig. 4.**
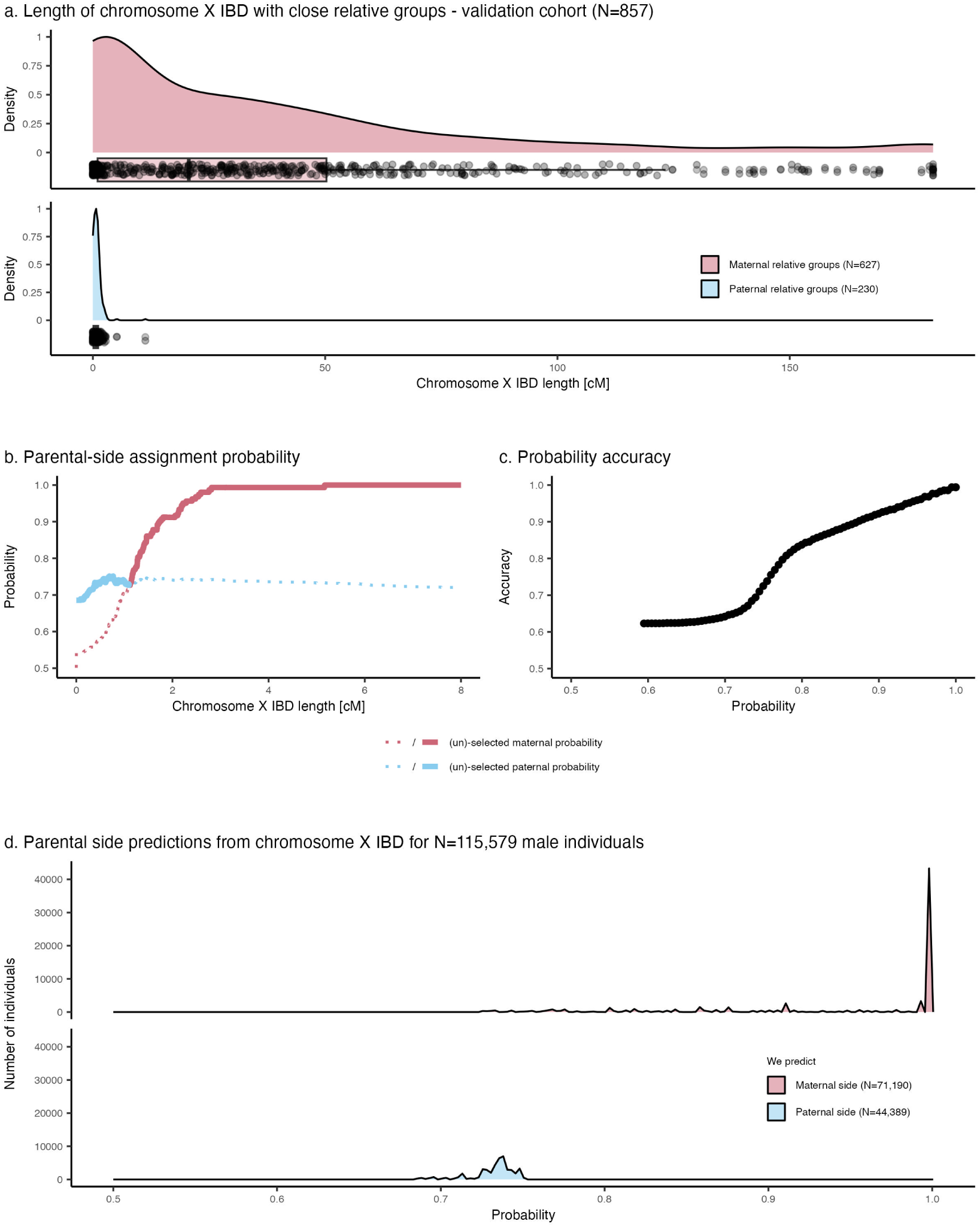
Parental side assignment from chromosome X IBD analysis in male individuals. **a**) Density distribution of chromosome X IBD segment length in centimorgan (cM) between targets and surrogate father (blue) and targets and surrogate mother (red) across the validation cohort’s male individuals. Boxes indicate the interquartile range (IQR), with the bottom and top of the box representing the 25th (Q1) and 75th (Q3) percentiles, respectively. The horizontal line within the box represents the median (50th percentile). Whiskers extend to the smallest and largest values within *Q*1*−*1.5 *× IQR* and *Q*3 + 1.5 *× IQR*. Each point represent a target-relative pair. **b**) Parental side probabilities depending on chromosome X IBD segment length derived from the validation cohort. Selected and unselected maternal (red) and paternal (blue) probabilities for a given x-axis value are indicated by solid and dotted lines, respectively. **c**) Accuracy of parental side predictions (y-axis) as a function of the parental side probability (x-axis). **d**) Distribution of maternal (red) and paternal (blue) side assignment probabilities derived chromosome X IBD across 115,579 UK Biobank male individuals.

**Supplementary Fig. 5.**
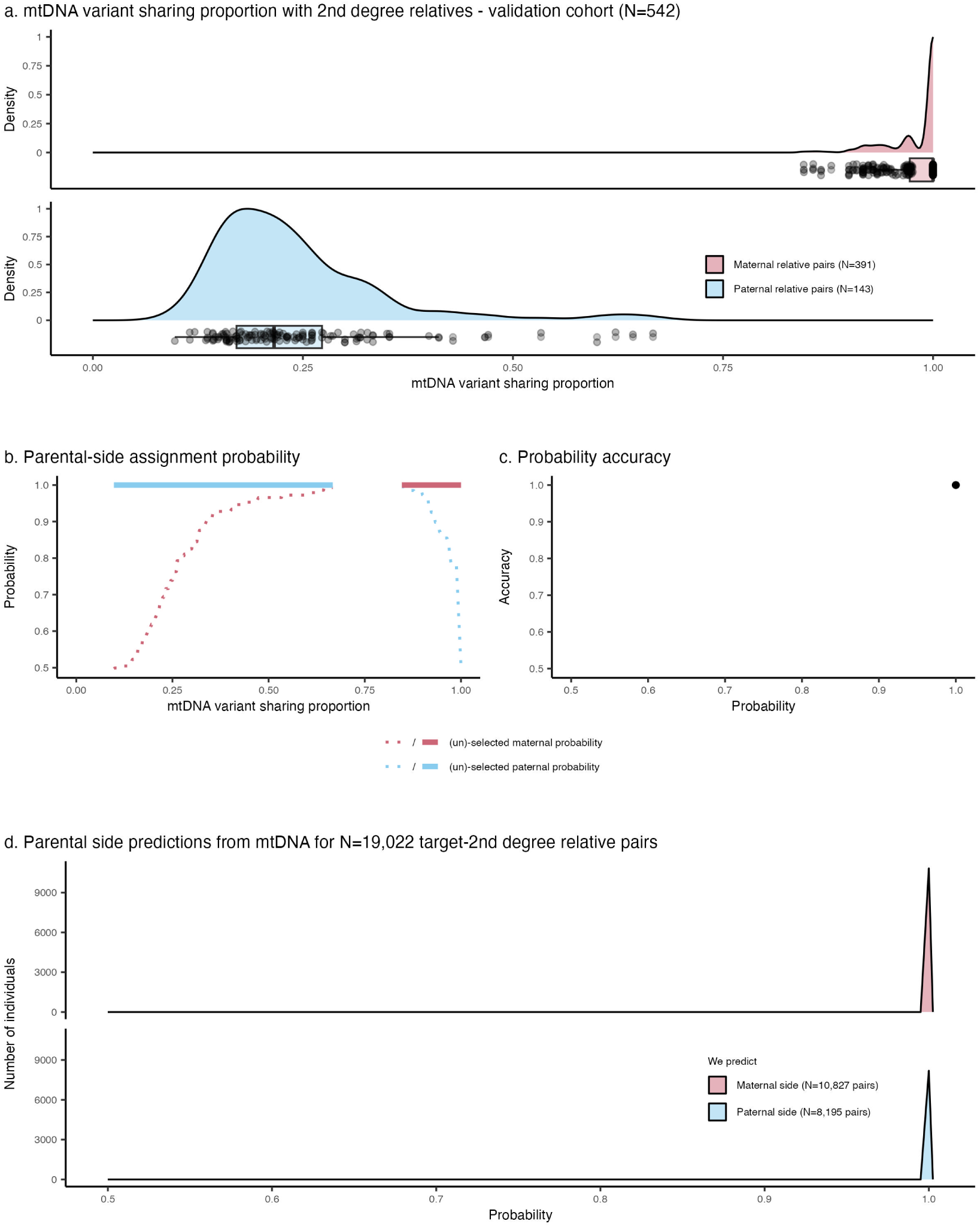
Parental side assignment from mitochondrial DNA Minor Variant Sharing (MVS) analysis in 2*^nd^* degree relative pairs. a) Density distribution of mtDNA Minor Variant Sharing (MVS) between targets and 2*^nd^*degree surrogate father (blue) and targets and 2*^nd^*degree surrogate mother (red) across the validation cohort’s individuals. Boxes indicate the interquartile range (IQR), with the bottom and top of the box representing the 25th (Q1) and 75th (Q3) percentiles, respectively. The horizontal line within the box represents the median (50th percentile). Whiskers extend to the smallest and largest values within *Q*1*−*1.5 *× IQR* and *Q*3 +1.5 *×IQR*. Each point represent a target-relative pair. **b**) Parental side probabilities depending on mtDNA MVS derived from the validation cohort. Selected and unselected maternal (red) and paternal (blue) probabilities for a given x-axis value are indicated by solid and dotted lines, respectively. **c**) Accuracy of parental side predictions (y-axis) as a function of the parental side probability (x-axis). **d**) Distribution of maternal (red) and paternal (blue) side assignment probabilities derived from mtDNA MVS across 19,022 UK Biobank 2*^nd^* degree relative pairs.

**Supplementary Fig. 6.**
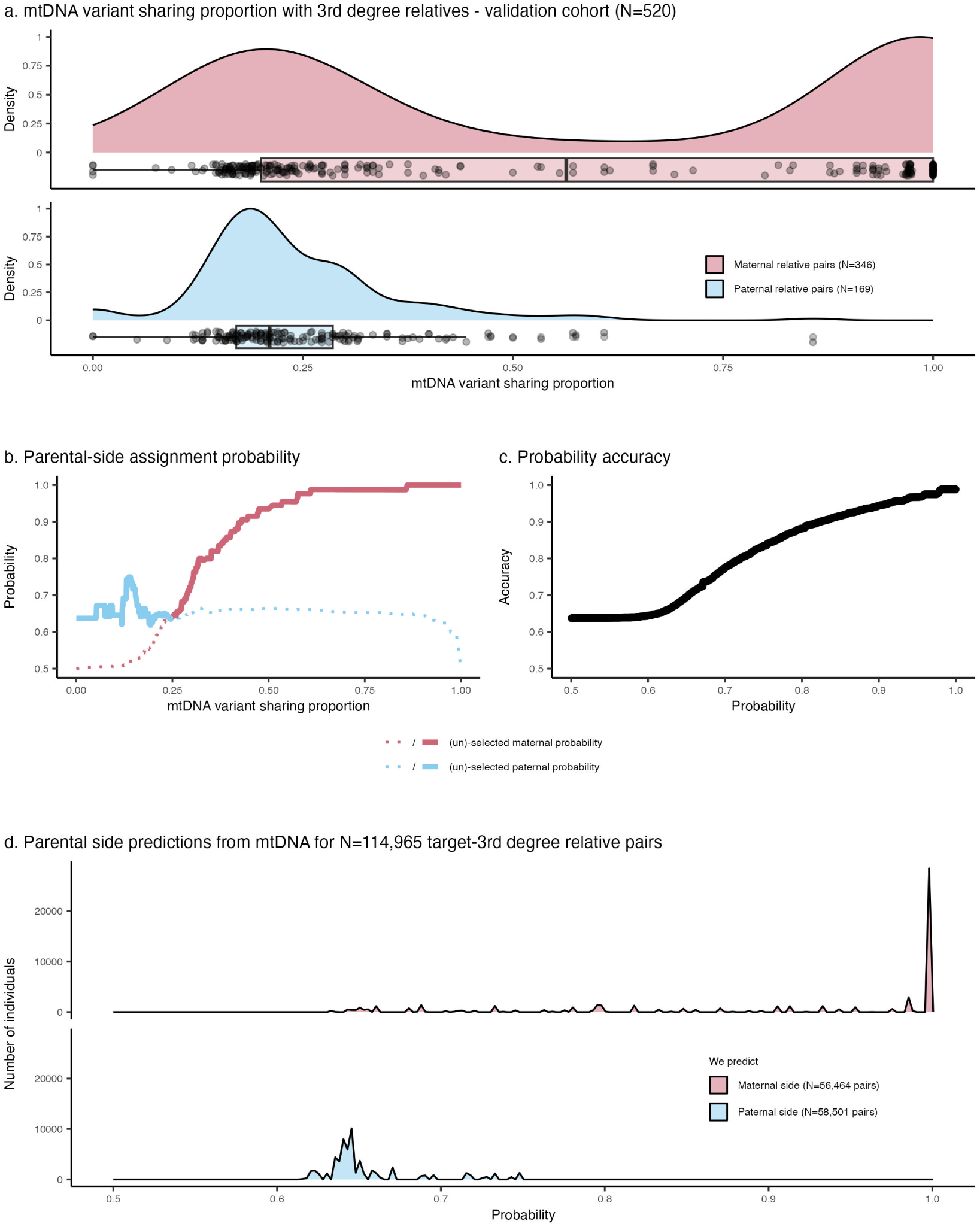
Parental side assignment from mitochondrial DNA Minor Variant Sharing (MVS) analysis in 3*^rd^* degree relative pairs. a) Density distribution of mtDNA Minor Variant Sharing (MVS) between targets and 3*^rd^* degree surrogate father (blue) and targets and 3*^rd^* degree surrogate mother (red) across the validation cohort’s individuals. Boxes indicate the interquartile range (IQR), with the bottom and top of the box representing the 25th (Q1) and 75th (Q3) percentiles, respectively. The horizontal line within the box represents the median (50th percentile). Whiskers extend to the smallest and largest values within *Q*1*−*1.5 *× IQR* and *Q*3 +1.5 *×IQR*. Each point represent a target-relative pair. **b**) Parental side probabilities depending on mtDNA MVS derived from the validation cohort. Selected and unselected maternal (red) and paternal (blue) probabilities for a given x-axis value are indicated by solid and dotted lines, respectively. **c**) Accuracy of parental side predictions (y-axis) as a function of the parental side probability (x-axis). **d**) Distribution of maternal (red) and paternal (blue) side assignment probabilities derived from mtDNA MVS across 114,965 UK Biobank 3*^rd^* degree relative pairs.

**Supplementary Fig. 7.**
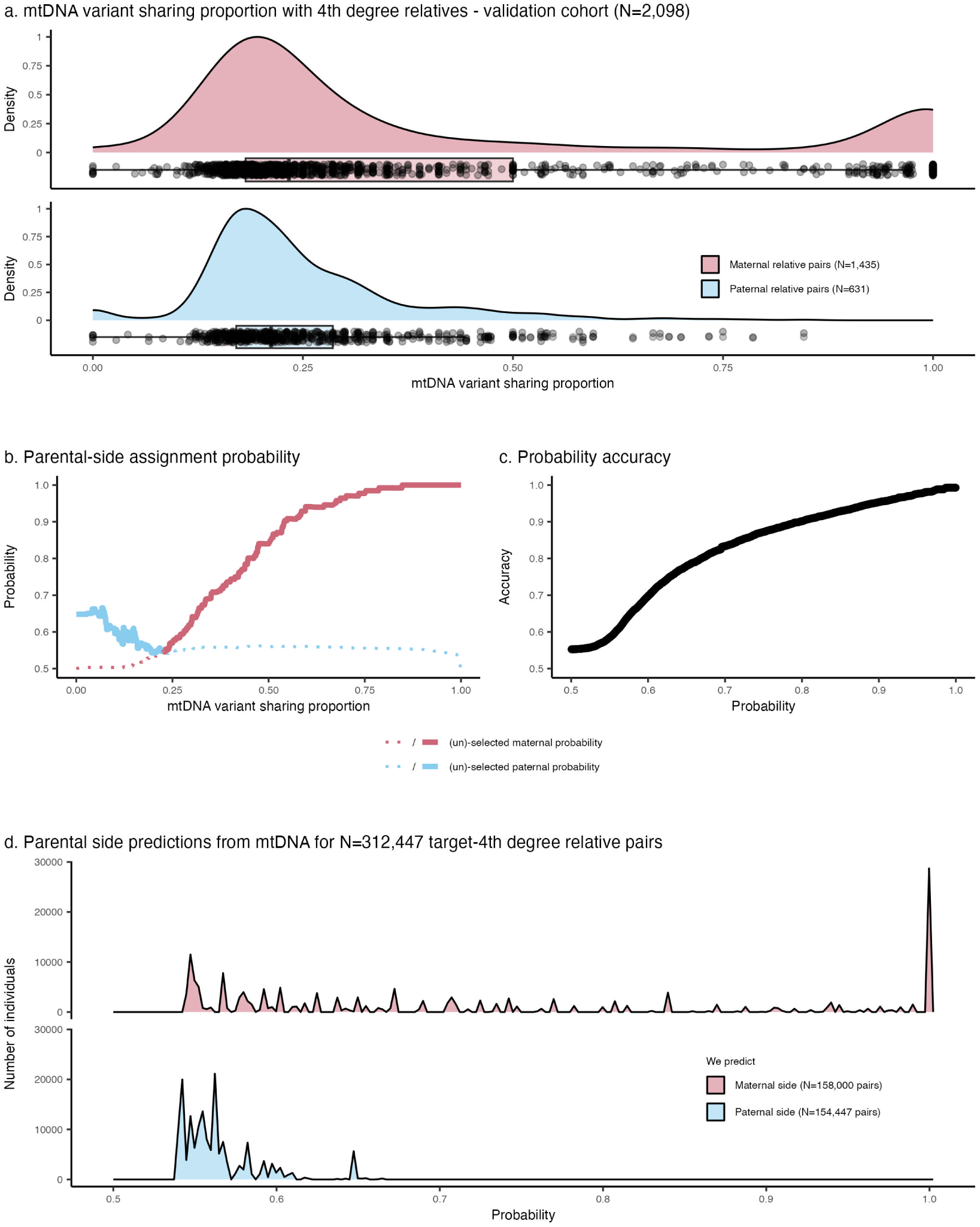
Parental side assignment from mitochondrial DNA Minor Variant Sharing (MVS) analysis in 4*^th^* degree relative pairs. a) Density distribution of mtDNA Minor Variant Sharing (MVS) between targets and 4*^th^* degree surrogate father (blue) and targets and 4*^th^* degree surrogate mother (red) across the validation cohort’s individuals. Boxes indicate the interquartile range (IQR), with the bottom and top of the box representing the 25th (Q1) and 75th (Q3) percentiles, respectively. The horizontal line within the box represents the median (50th percentile). Whiskers extend to the smallest and largest values within *Q*1*−*1.5 *× IQR* and *Q*3 +1.5 *×IQR*. Each point represent a target-relative pair. **b**) Parental side probabilities depending on mtDNA MVS derived from the validation cohort. Selected and unselected maternal (red) and paternal (blue) probabilities for a given x-axis value are indicated by solid and dotted lines, respectively. **c**) Accuracy of parental side predictions (y-axis) as a function of the parental side probability (x-axis). **d**) Distribution of maternal (red) and paternal (blue) side assignment probabilities derived from mtDNA MVS across 312,447 UK Biobank 4*^th^* degree relative pairs.

**Supplementary Fig. 8.**
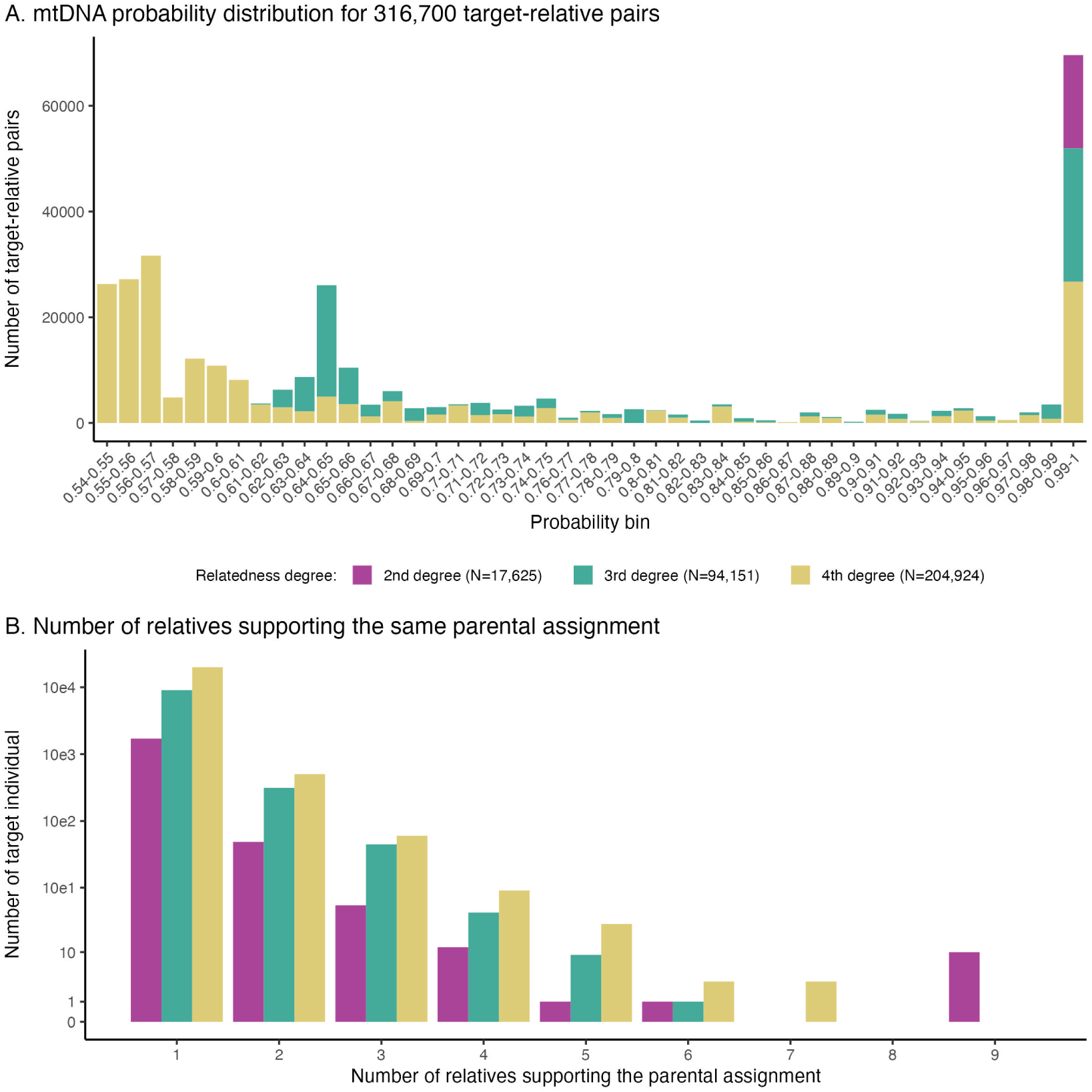
Selection of mitochondrial DNA (mtDNA) Minor Variant Sharing (MVS) predictors for parental side assignments. a) Distribution of parental side assignment probabilities for 316,700 target-relative pairs, stratified by relatedness degree (2*^nd^*, 3*^rd^*, and 4*^th^*degree). For individuals with multiple relatives of the same degree, only the relative with the highest predicted accuracy was retained. Probabilities are binned at intervals of 0.01 for visualization. **b**) Log-scale distribution of the number of target individuals (y-axis) based on the number of relatives (x-axis) supporting the same parental side assignment, further stratified by degree of relatedness.

**Supplementary Fig. 9.**
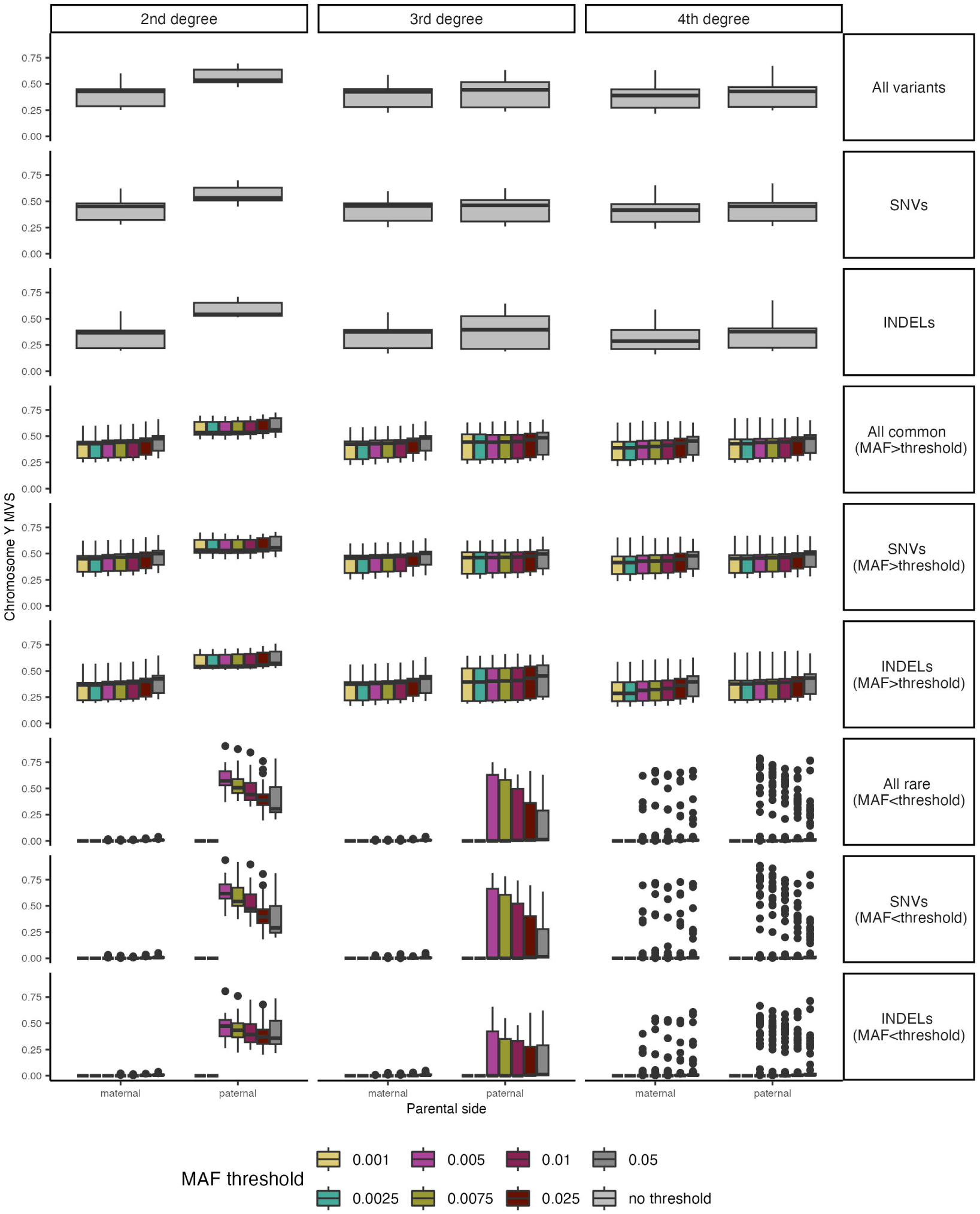
Chromosome Y Minor Variant Sharing (MVS) analysis. The y-axis represents the proportion of chromosome Y minor variants shared (MVS), stratified by parental side assignments (maternal or paternal, x-axis), relatedness degrees (columns: 2*^nd^*, 3*^rd^*, and 4*^th^* degree), variant categories (rows), and minor allele frequency (MAF) thresholds (color-coded). Categories include all variants, single nucleotide variants (SNVs), insertions/deletions (INDELs), common variants (MAF *>* threshold), and rare variants (MAF *<* threshold). The MAF thresholds are denoted in the legend for clearer distinction of variant frequencies.

**Supplementary Fig. 10.**
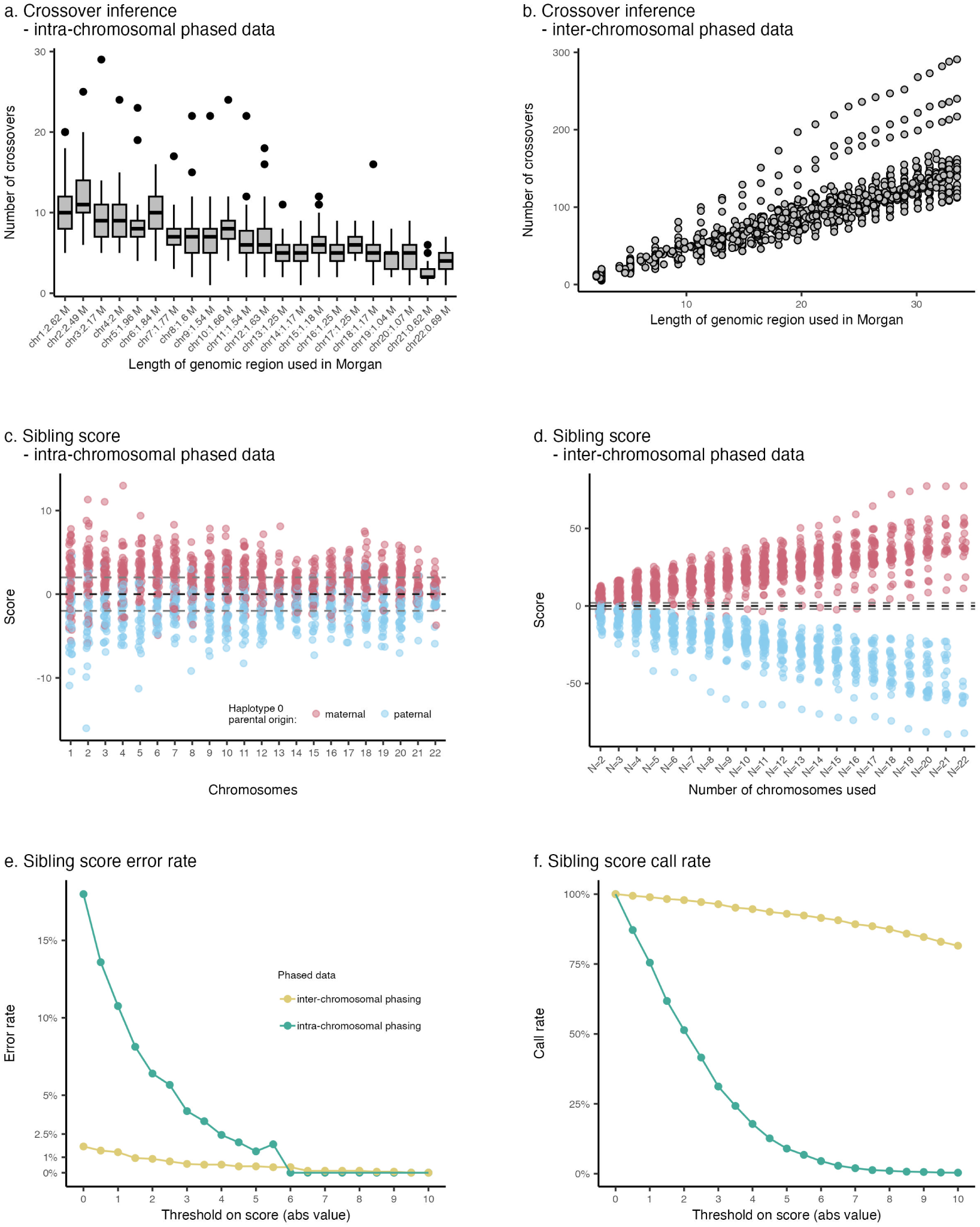
Evaluation of sibling scores from intra- and inter-chromosomal phasing in N=88 individuals of the validation cohort. a) Distribution of the number of inferred crossovers (y-axis) per chromosome (x-axis) derived from intra-chromosomally phased data. **b**) Distribution of the number of inferred crossovers (y-axis) relative to the genomic length used (in Morgans, x-axis) derived from inter-chromosomally phased data. **c**) Distribution of sibling scores (y-axis) per chromosome (x-axis) using intra-chromosomally phased data. Maternal and paternal haplotypes are color-coded in red and blue, respectively. **d**) Distribution of sibling scores (y-axis) relative to the number of chromosomes included in inter-chromosomally phased data (x-axis). **e**) Error rates (y-axis) of sibling scores for varying score thresholds (absolute values, x-axis). For example, excluding ambiguous scores between −2 and 2 reduced error rates to 1.4% with inter-chromosomal phasing, compared to 6.4% with intra-chromosomal phasing. **f**) Call rates (y-axis) of sibling scores for varying score thresholds (absolute values, x-axis), showing the proportion of individuals meeting the threshold criteria.

**Supplementary Fig. 11.**
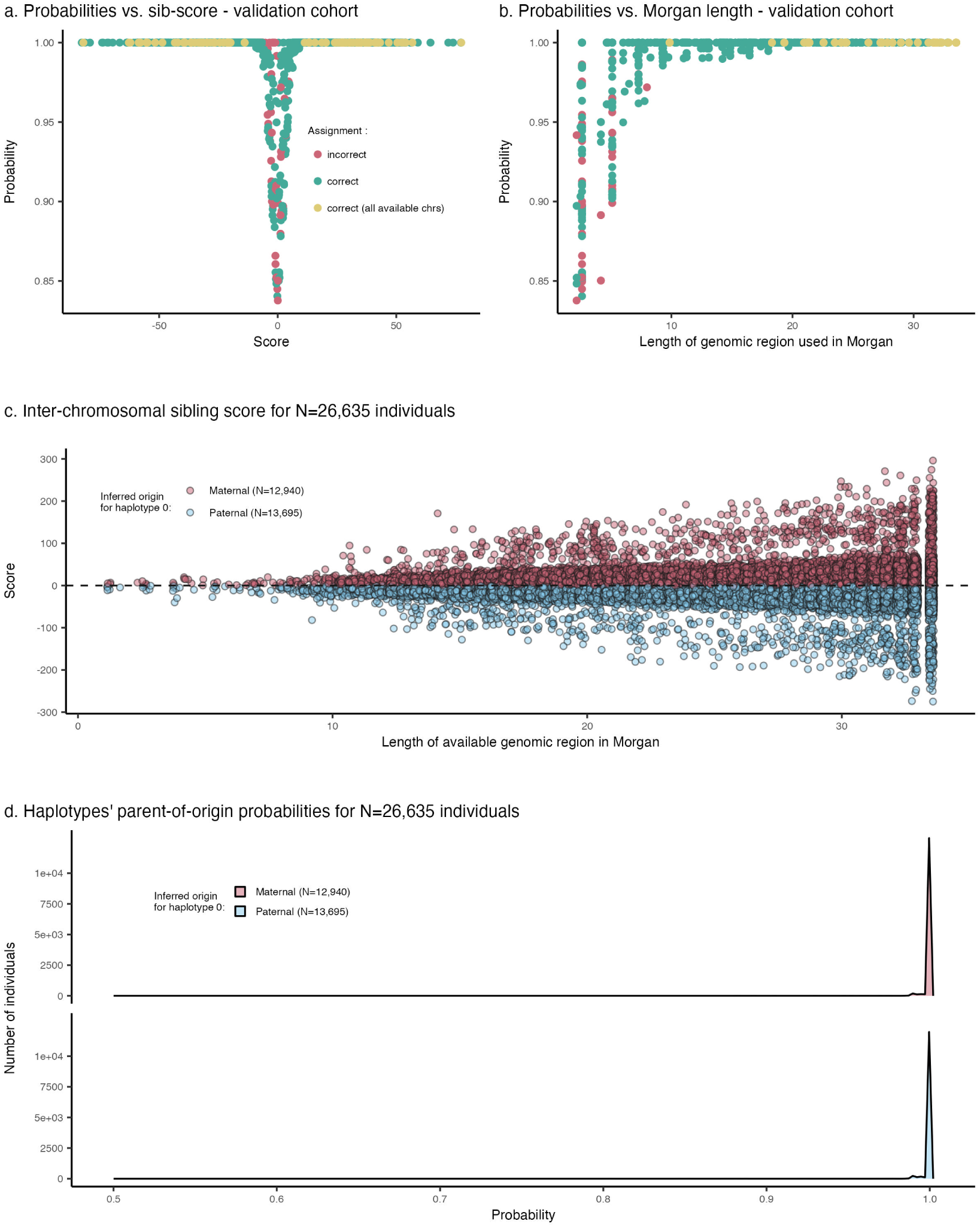
Validation and derivation of parent-of-origin probabilities using sibling scores from inter-chromosomal phased data. a) PofO probabilities (y-axis) as a function of sibling scores (x-axis) in the validation cohort. Each dot represents a specific configuration (an individual with a given number of chromosomes used). We varied the number of chromosome used per individual to assess the accuracy of different configurations (see Supplementary Figure 10D). Red dots indicate incorrect parental assignments, green dots indicate correct assignments, and yellow dots denote correct assignments using the maximum available chromosomes (*N_max_*, corresponds to the full set of inter-chromosomally phased chromosome for a given individual). No errors were observed when *N_max_* chromosomes were used. **b**) PofO probabilities (y-axis) plotted against the total available genomic length in Morgans (x-axis) for sibling score calculation in the validation cohort. Each configuration is represented as a dot (color-coded as in panel a). Increasing genomic length results in higher assignment accuracy, with perfect accuracy achieved at *N_max_* (yellow dots). **c**) Sibling scores (y-axis) derived for N=26,635 individuals across available genomic lengths (x-axis). Most configurations (99.5%) cover more than 10 Morgans and include over 30 inferred crossover events, ensuring high assignment accuracy (as indicated in the validation panels a and b). **d**) Distribution of PofO probabilities for 26,635 individuals. A majority of individuals (*>*95%) exhibit probabilities of 1, resulting in perfect accuracy within the validation cohort (see yellow dots in panel a).

**Supplementary Fig. 12.**
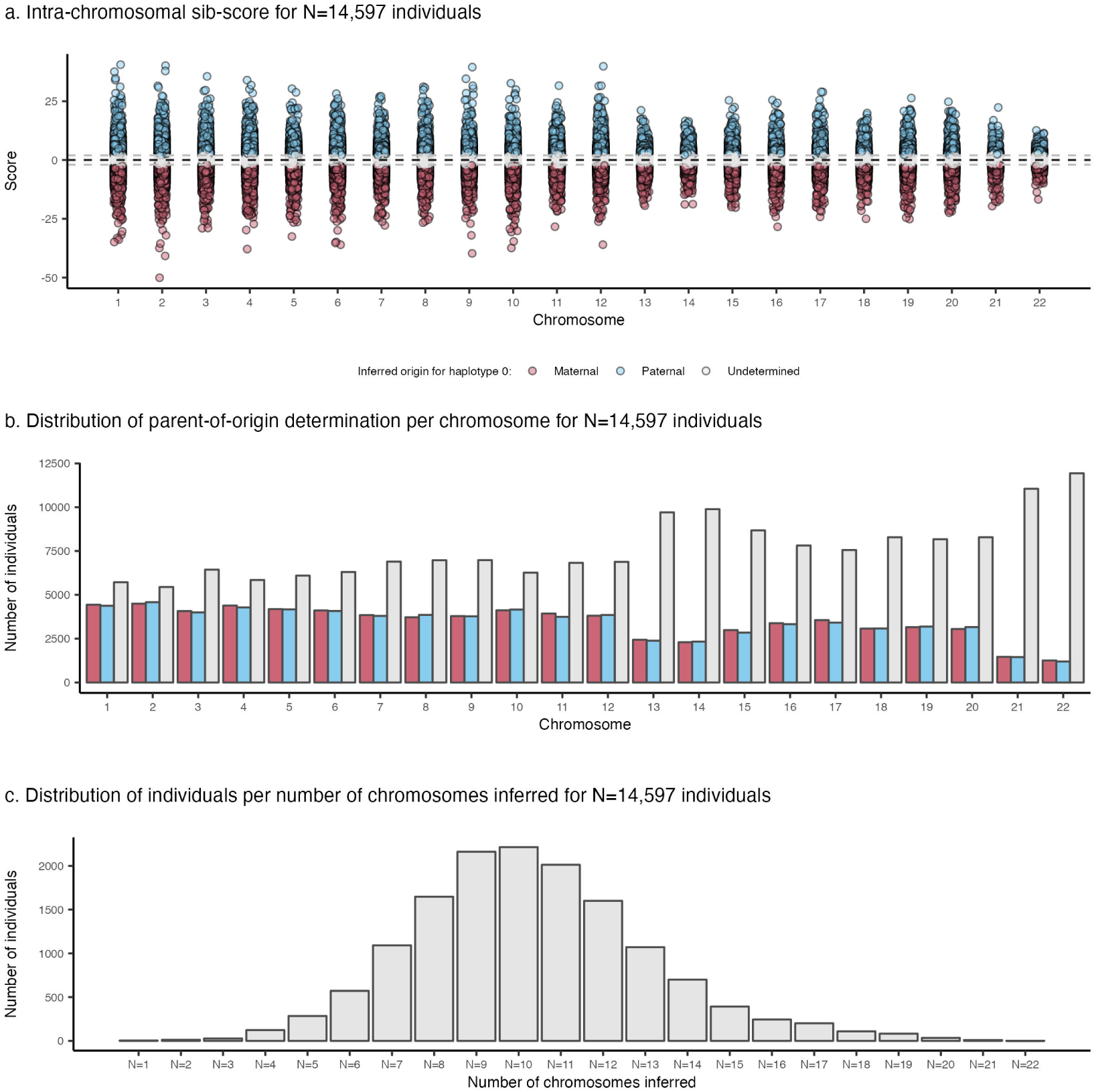
Derivation of parent-of-origin probabilities using sibling scores from intra-chromosomal phased data. **a**) Sibling scores calculated for each chromosome independently (x-axis). Each dot corresponds to a single chromosome, with blue and red dots indicating paternal and maternal assignment, respectively, for the first haplotype. Undetermined assignments are shown in gray. **b**) Distribution of the number of individuals (y-axis) with inferred PofO for each chromosome (x-axis). Larger chromosomes exhibit higher numbers of PofO inference (e.g., 8,811 individuals for chromosome 1), while smaller chromosomes show fewer inferences (e.g., 2,450 individuals for chromosome 22), reflecting the increased likelihood of crossovers on larger chromosomes. **c**) Distribution of the number of chromosomes with inferred PofO per individual (x-axis), with counts shown on the y-axis. On average, individuals have 10.3 chromosomes with PofO inferred. This is lower compared to the average of 16 chromosomes inferred using inter-chromosomal phasing (see Supplementary Figure 3B).

**Supplementary Fig. 13.**
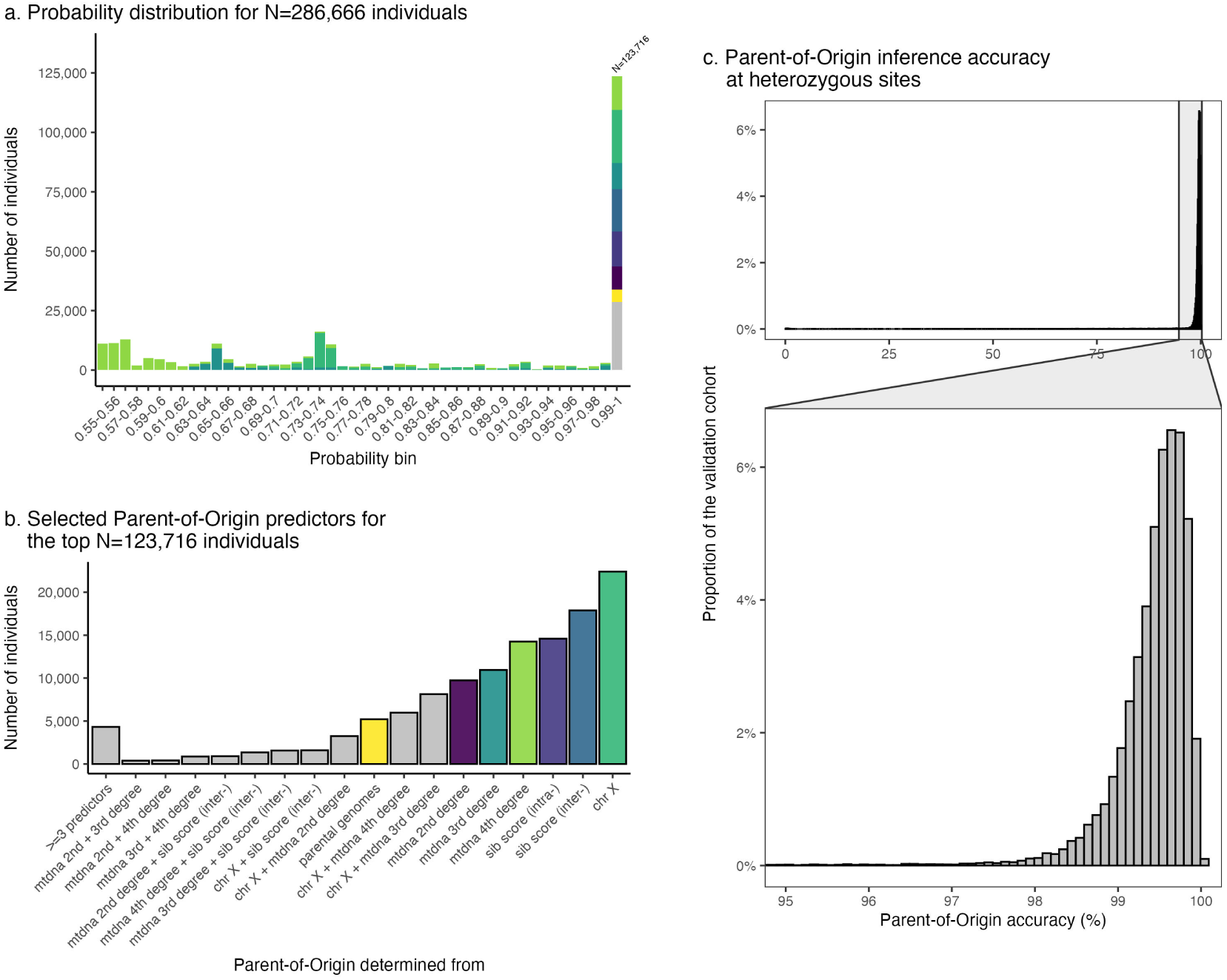
Selected parent-of-origin predictors and error rate. **a**) Distribution of PofO probabilities for N=286,666 individuals. The x-axis represents probability bins, and the y-axis shows the number of individuals. Colors correspond to the predictors used for PofO determination, as indicated in panel (b). **b**) Distribution of selected PofO predictors for individuals having a PofO probability greater than 0.99. **c**) Distribution of the PofO inference accuracy. Errors were computed at heterozygous site only by comparing the PofO determined using our approach to the one obtained from parental genomes. We found an average accuracy of 97.94%. This rate is impacted by individuals for which entire haplotypes are incorrect, resulting from inter-chromosomal phasing errors, totaling 64 haplotypes (in the validation cohort). These are likely due to the presence of relatives sharing both paternal and maternal IBD segments, which may be due to consanguinity, and were not filtered out during the construction of the validation cohort. As a result, only a few individuals’ haplotypes decrease the global accuracy, and most individuals have a PofO correctly assigned. Indeed, the majority of haplotypes (83.9%) exhibit accuracy above 99%, and half of the haplotypes have more than 0.52% accuracy.

**Supplementary Fig. 14.**
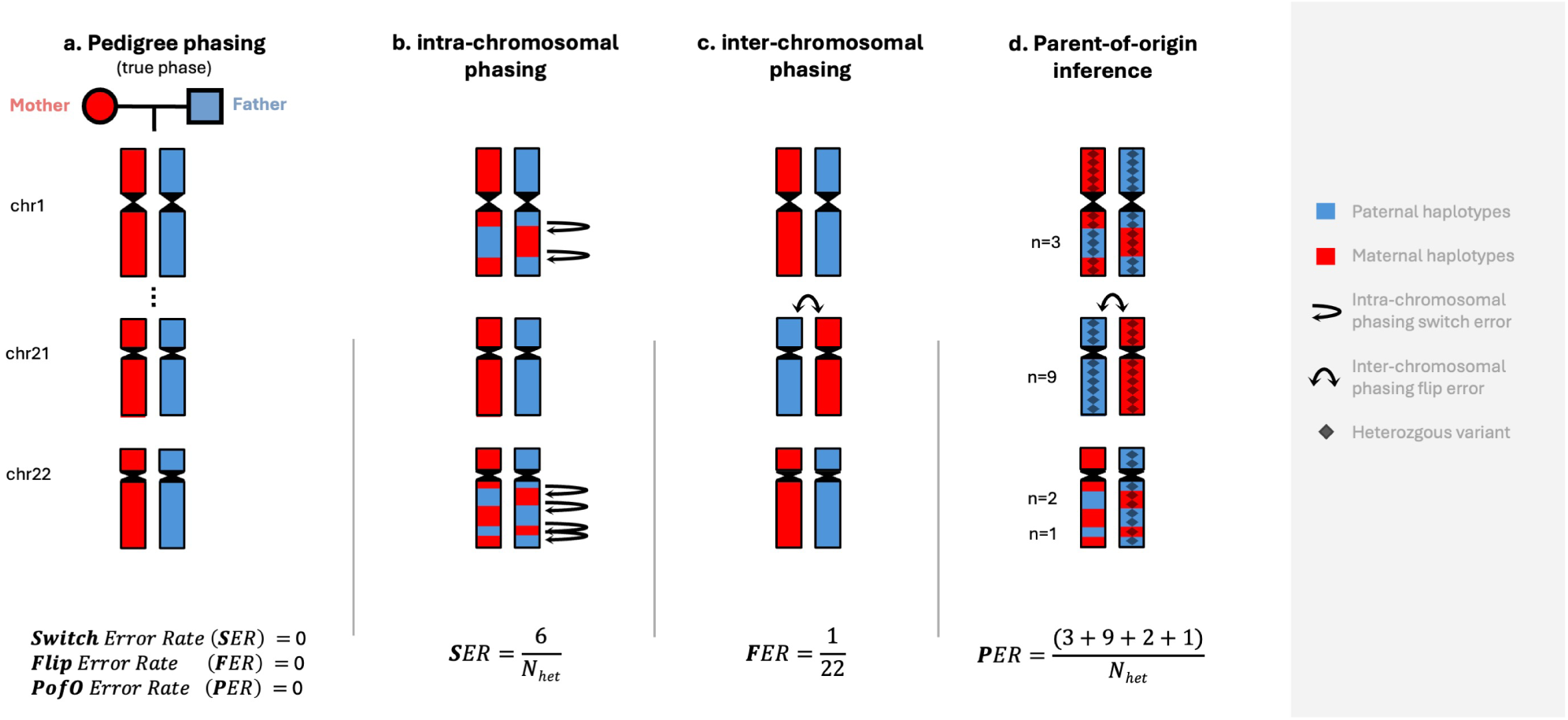
Illustration of errors in statistical methods. **a**) A perfectly phased individual (both intra- and inter-chromosomally). Phasing is performed using the parental genomes: maternal (red) and paternal (blue). In this scenario, the PofO is directly inferred alongside the phasing process without errors. **b**) An individual with intra-chromosomal phasing errors introduced by traditional phasing software (e.g., SHAPEIT5^20^). These errors are represented as switches between parental haplotypes within the same chromosome (horizontal arrows), with 6 switches shown here. Since switches occur only at heterozygous sites, the switch error rate (SER) is calculated as *SER* = 6*/N_het_*, where *N_het_* is the total number of heterozygous sites. **c**) An individual with inter-chromosomal phasing errors but assumed perfect intra-chromosomal phasing (no switches within the same chromosome). Errors in this case are represented as flips of entire parental haplotypes across chromosomes (vertical arrow), with one flip shown here. The flip error rate (FER) is calculated as the proportion of chromosomes flipped relative to the majority, ranging from 0% to 50%. **d**) An individual with combined intra- and inter-chromosomal phasing errors. PofO inference errors aggregate contributions from both switch and flip errors. The PofO error rate (PER) is computed as the proportion of heterozygous sites (grey diamonds) incorrectly assigned. For example, in this scenario, assigning the first haplotype as maternal and the second as paternal results in a total of 15 errors.

**Supplementary Fig. 15.**
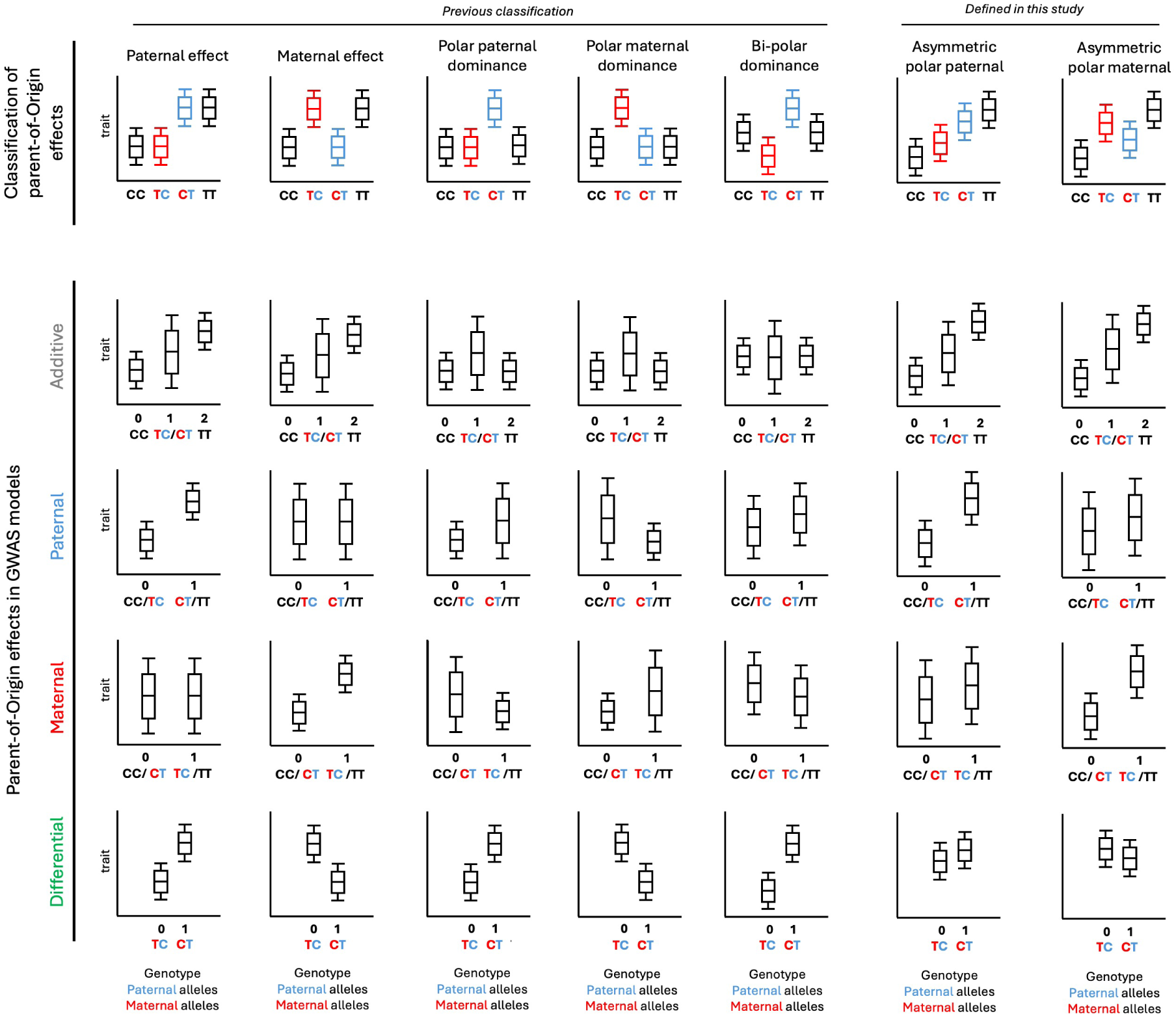
Classification and detection of parent-of-origin effects in GWAS. Top panel. illustrates the classification of POEs^14^ and shows the variation in trait values (y-axis) across genotypes and parental origin of alleles (x-axis). Maternal alleles are shown in red, and paternal alleles are shown in blue. Boxplots represent the distribution of trait values for different genotypes: black boxes for homozygotes, red boxes for maternal heterozygotes, and blue boxes for paternal heterozygotes. Each boxplot includes the 25th, 50th (median), and 75th percentiles, with whiskers extending to the minimum and maximum values. **Bottom panels** show the interpretation of the different POEs (x-axis) using various GWAS models (y-axis), including additive, paternal, maternal, and differential models. In this study, we identified POEs using the differential GWAS model.

**Supplementary Fig. 16.**
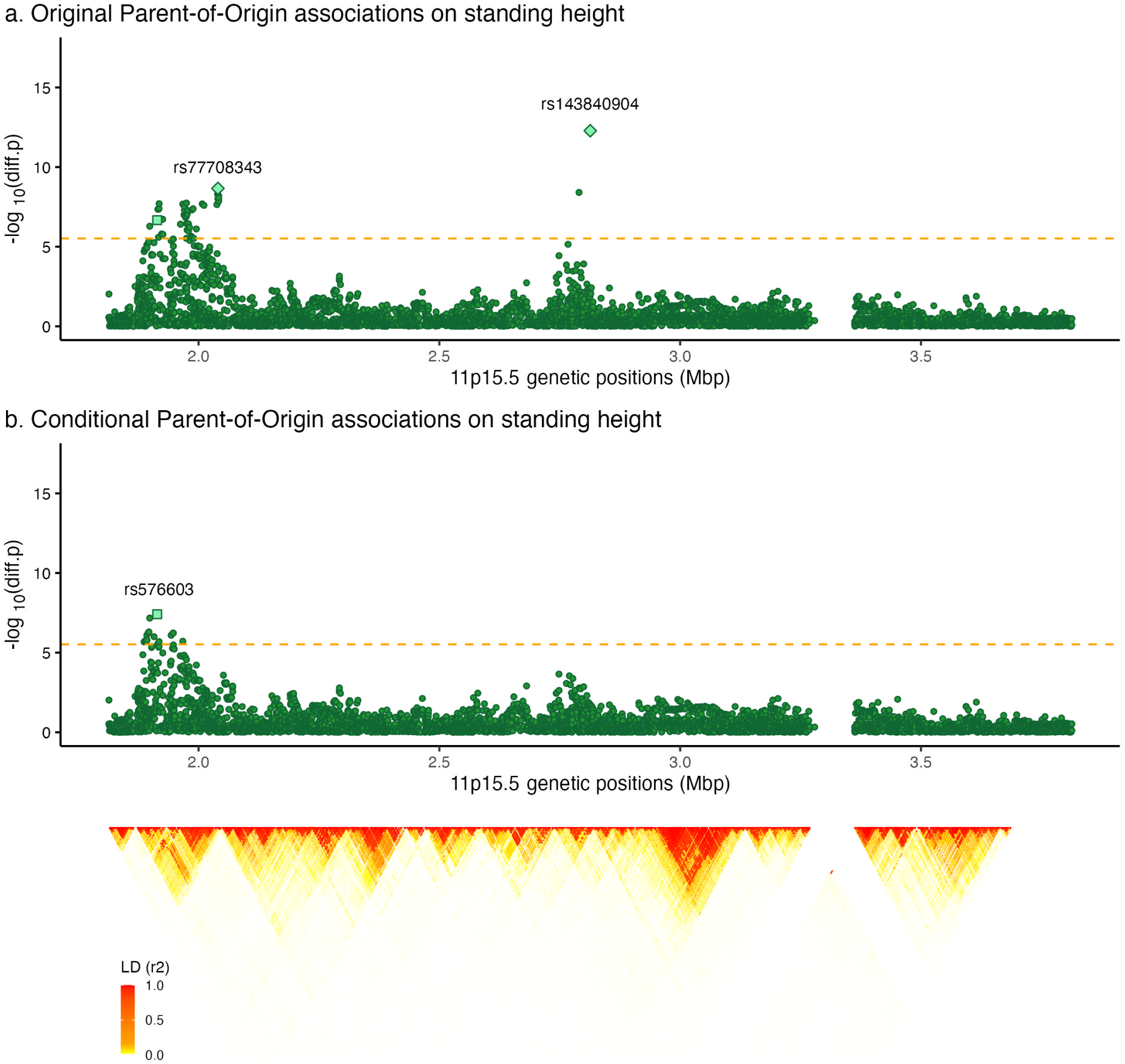
Conditional analysis on standing height at 11p15.5. **a**) Original, and **b**) Conditional differential GWAS show the association strength (*−log*_10_(*p − value*), y-axis) against the genomic position (x-axis) at the 11p15.5 region. Each point represents a genetic variant. Diamonds indicate the variants with independent, significant POE on standing height found in this study in the primary GWAS scan. The square represents the variant with significant and independent POE identified in the conditional analysis. Orange dashed lines represent the significance threshold used in this study when focusing on imprinted regions (3.1 *×* 10^−06^). Bottom panel shows the Linkage Disequilibrium (LD) pattern, ranging from LD=0 (white) to LD=1 (red).

**Supplementary Fig. 17.**
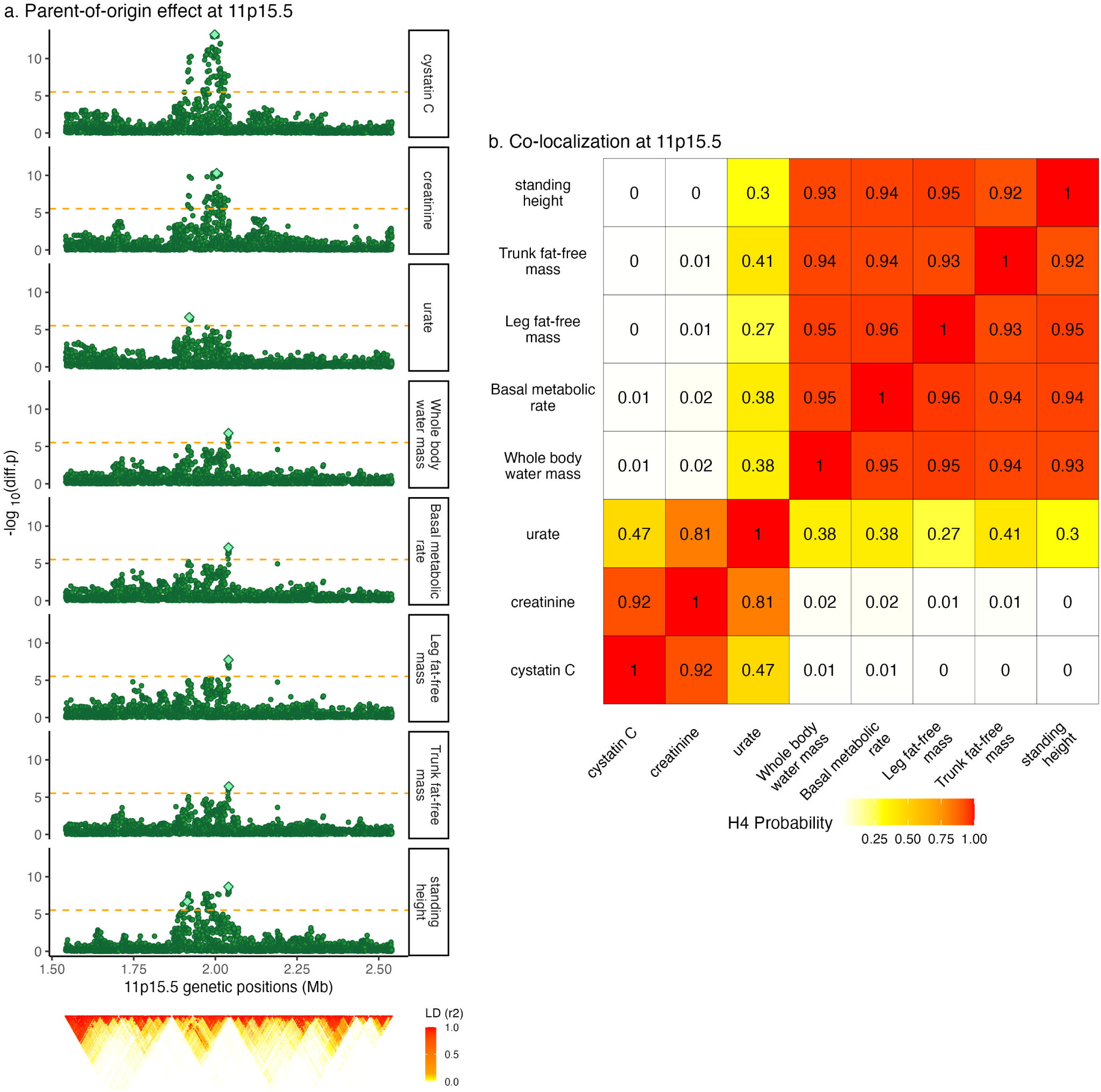
Co-localization analyses at 11p15.5. **a**) Parent-of-origin associations across traits at 11p15.5 show the differential GWAS association strength (*−log*_10_(*p − value*), y-axis) against the genomic position (x-axis). Orange dashed lines represent the significance threshold used in this study when focusing on imprinted regions (3.1 *×* 10*^−^*^6^). Each point represents a genetic variant. Light green diamonds shows lead POE reported in Table 1. **b**) Co-localization probability heatmap (i.e, shared causal variant probability *H*_4_).

**Supplementary Fig. 18.**
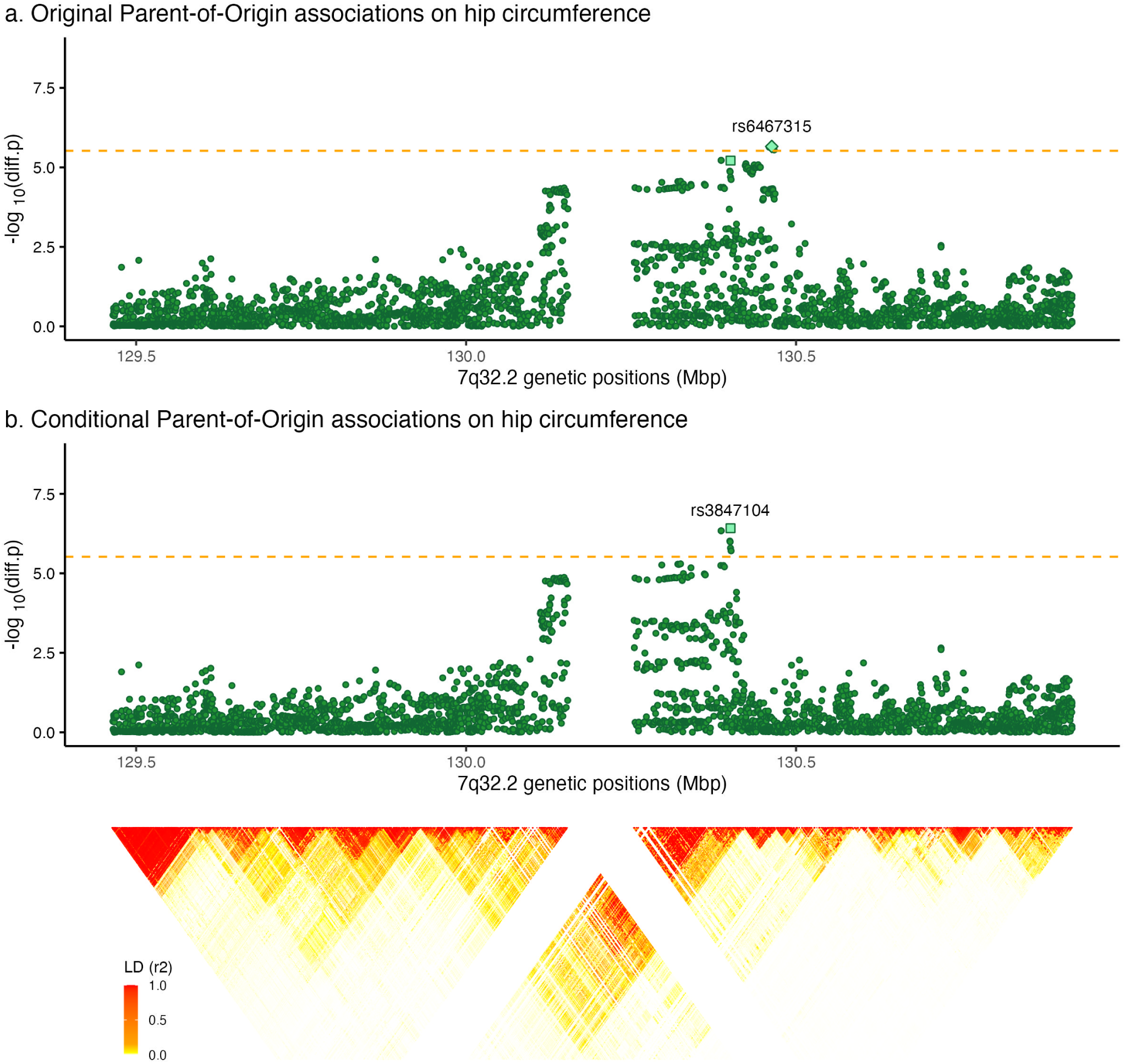
Conditional analysis on hip circumference at 7q32.2. **a**) Original, and **b**) Conditional differential GWAS show the association strength (*−log*_10_(*p−value*), y-axis) against the genomic position (x-axis) at the 7q32.2 region. Each point represents a genetic variant. The diamond indicates the variant with significant POE on hip circumference found in this study in the primary GWAS scan. The square indicates the variant with significant and independent POE identified in the conditional analysis. Orange dashed lines represent the significance threshold used in this study when focusing on imprinted regions (3.1 *×* 10*^−^*^6^). Bottom panel shows the Linkage Disequilibrium (LD) pattern, ranging from LD=0 (white) to LD=1 (red).

**Supplementary Fig. 19.**
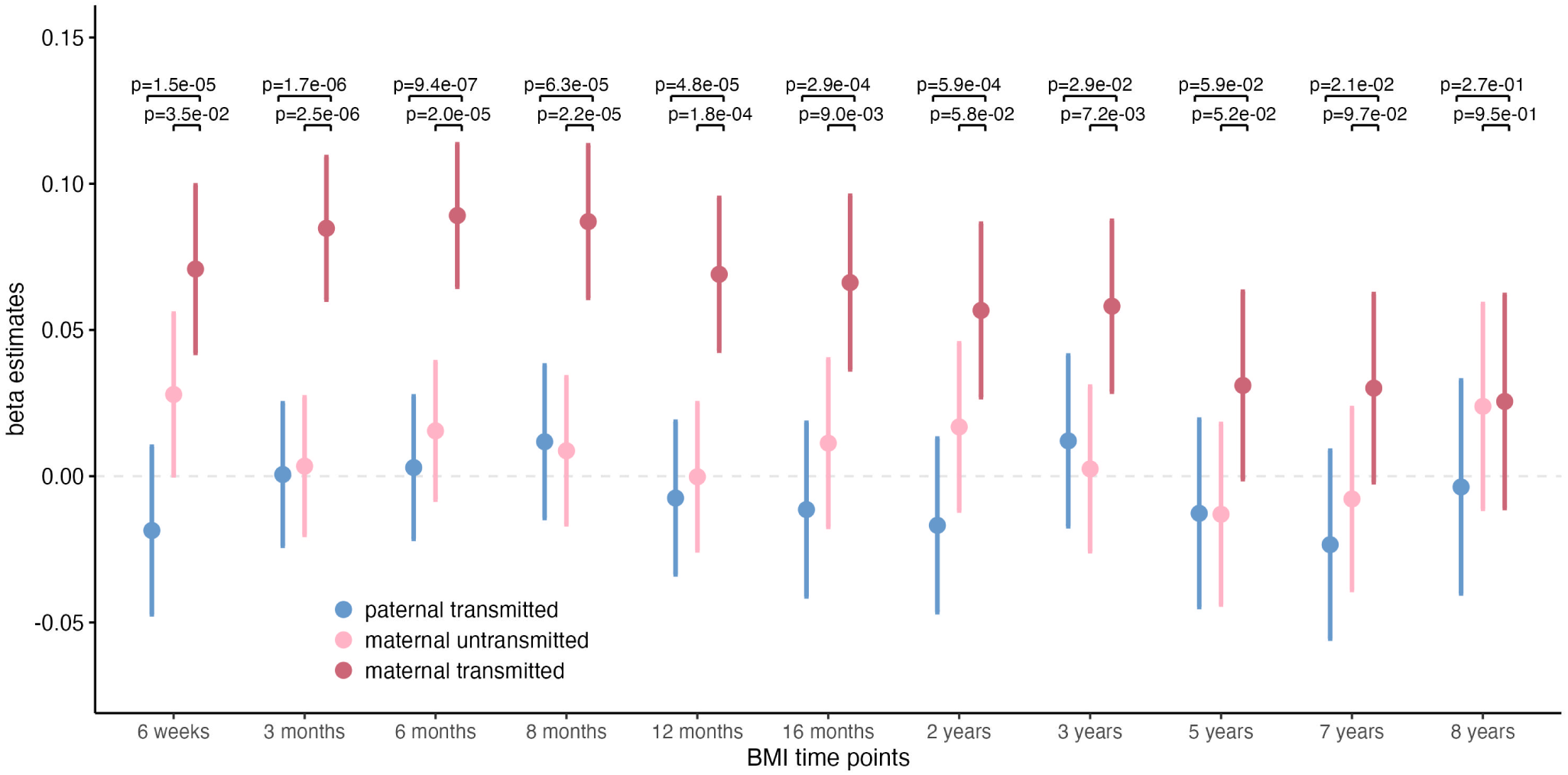
Parent-of-origin effects of rs6467315 on different BMI time points. Beta estimates and 95% confidence intervals (y-axis) of paternal transmitted (blue), maternal transmitted (red) and maternal untransmitted (pink) alleles effects on different BMI time points (x-axis). We assessed the differences in effects between (i) maternal and paternal transmitted alleles and (ii) maternal transmitted and untransmitted alleles using a Z-score approach, analogous to a differential GWAS test.

**Supplementary Fig. 20.**
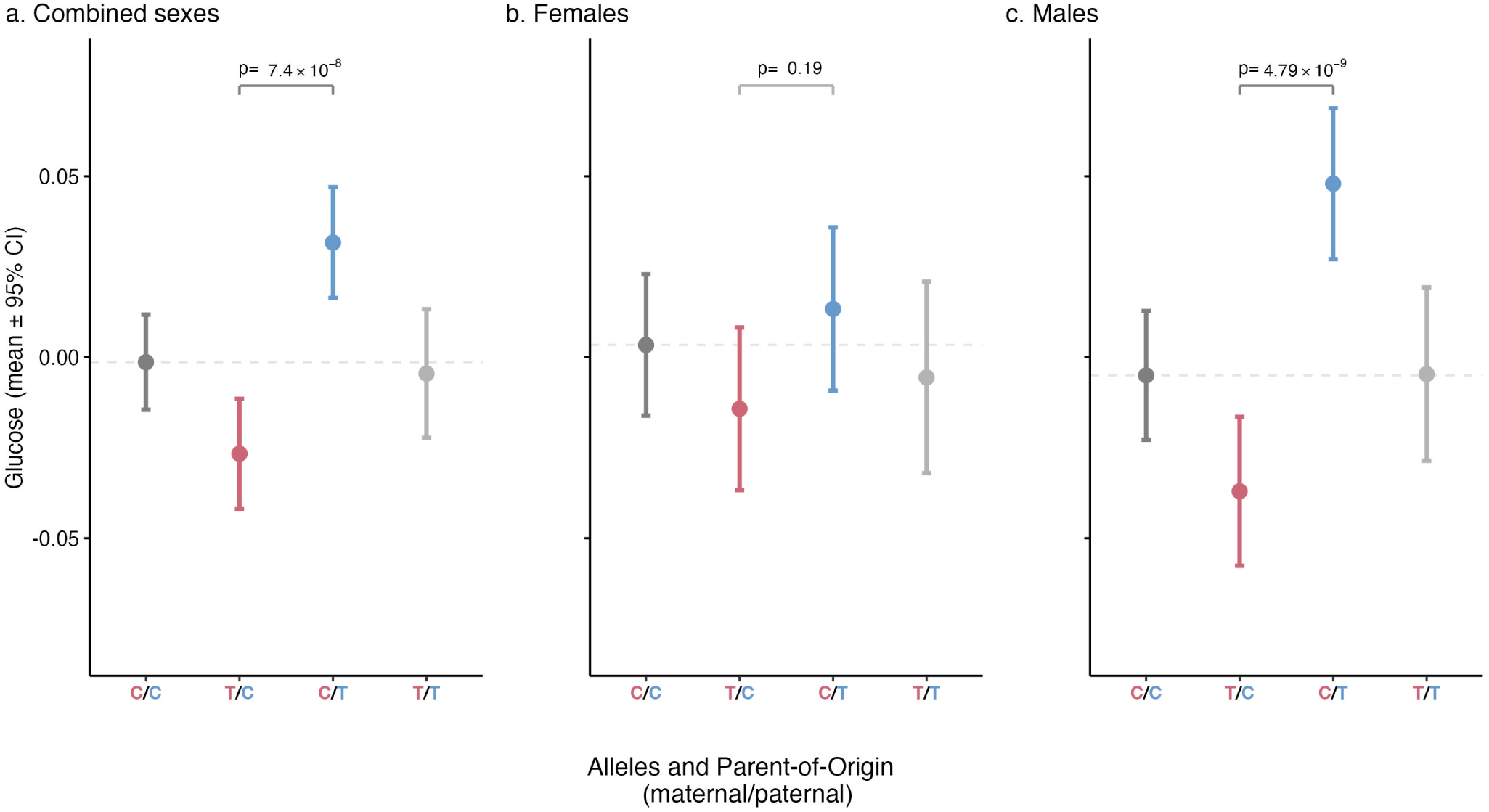
Sex-specific POE of rs4417225 on glucose levels. Effects of genotypes and PofO of alleles (x-axis) on glucose level (y-axis) for (**a**) both sexes combined, (**b**) females only, and (**c**) males only. Red markers represent maternal heterozygotes, blue markers represent paternal heterozygotes, and dark and light grey represent homozygotes. The data points show the mean glucose levels, with error bars indicating the 95% confidence intervals. A grey dashed line represents the mean glucose level for individuals with the reference genotype (C/C). Significance values from the differential GWAS tests are annotated within each panel.

**Supplementary Fig. 21.**
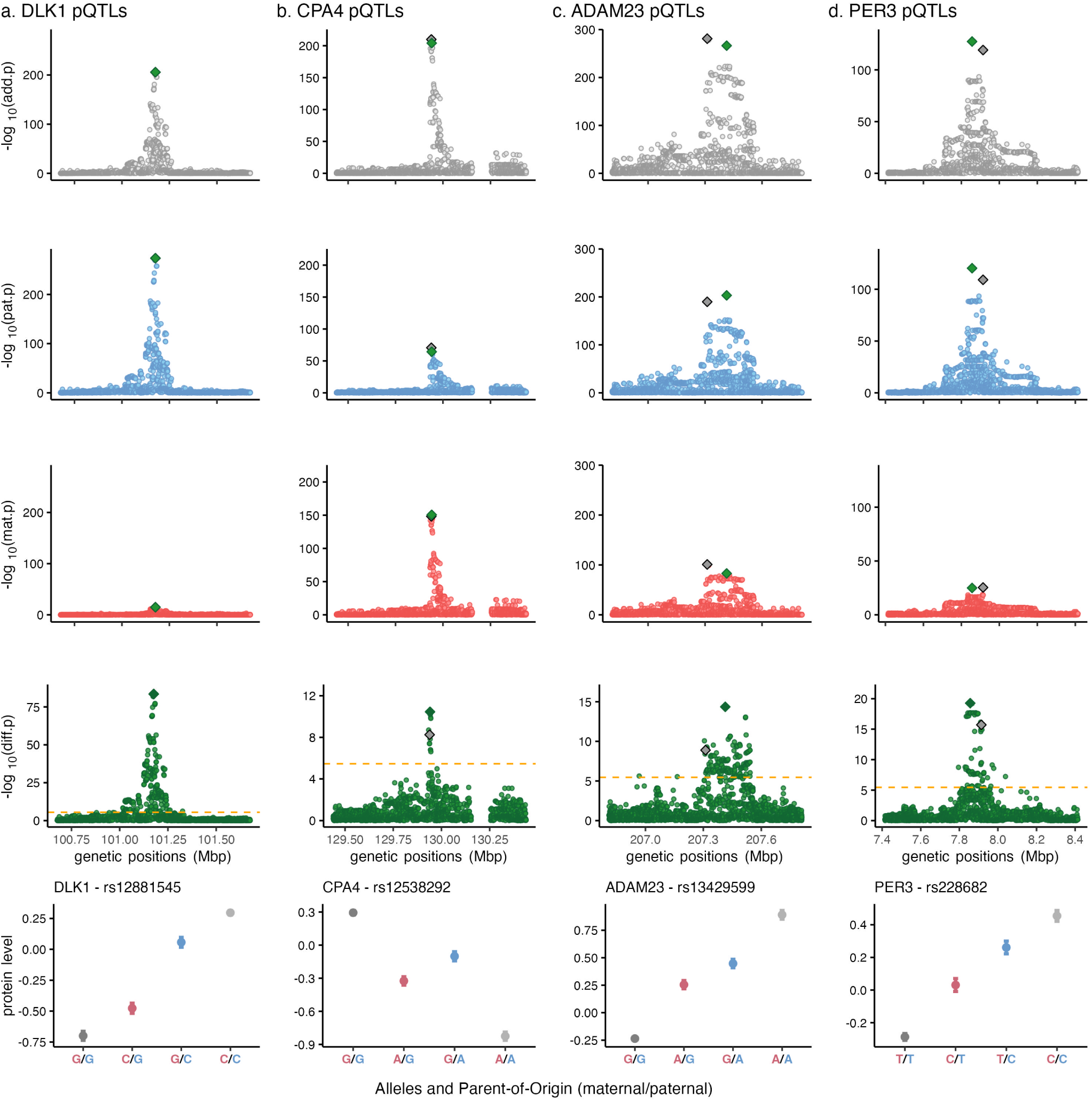
Significant Parent-of-Origin pQTLs. Association strength (*−log*_10_(*p − value*), y-axis) against the genomic position (x-axis) for additive (grey), paternal (blue), maternal (red) and differential (green) GWAS on (**a**) DLK1, (**b**) CPA4, (**c**) ADAM23 and (**d**) PER3. Diamonds show the primary additive association previously reported^25^ (grey) and the primary POE-pQTL detected in this study (green). Orange dashed line indicate the significance threshold (*P_D_ <* 0.05*/*14285 = 3.5 *×* 10*^−^*^6^). **Bottom panel** show the effects of alleles and PofO on normalized protein levels.

**Supplementary Fig. 22.**
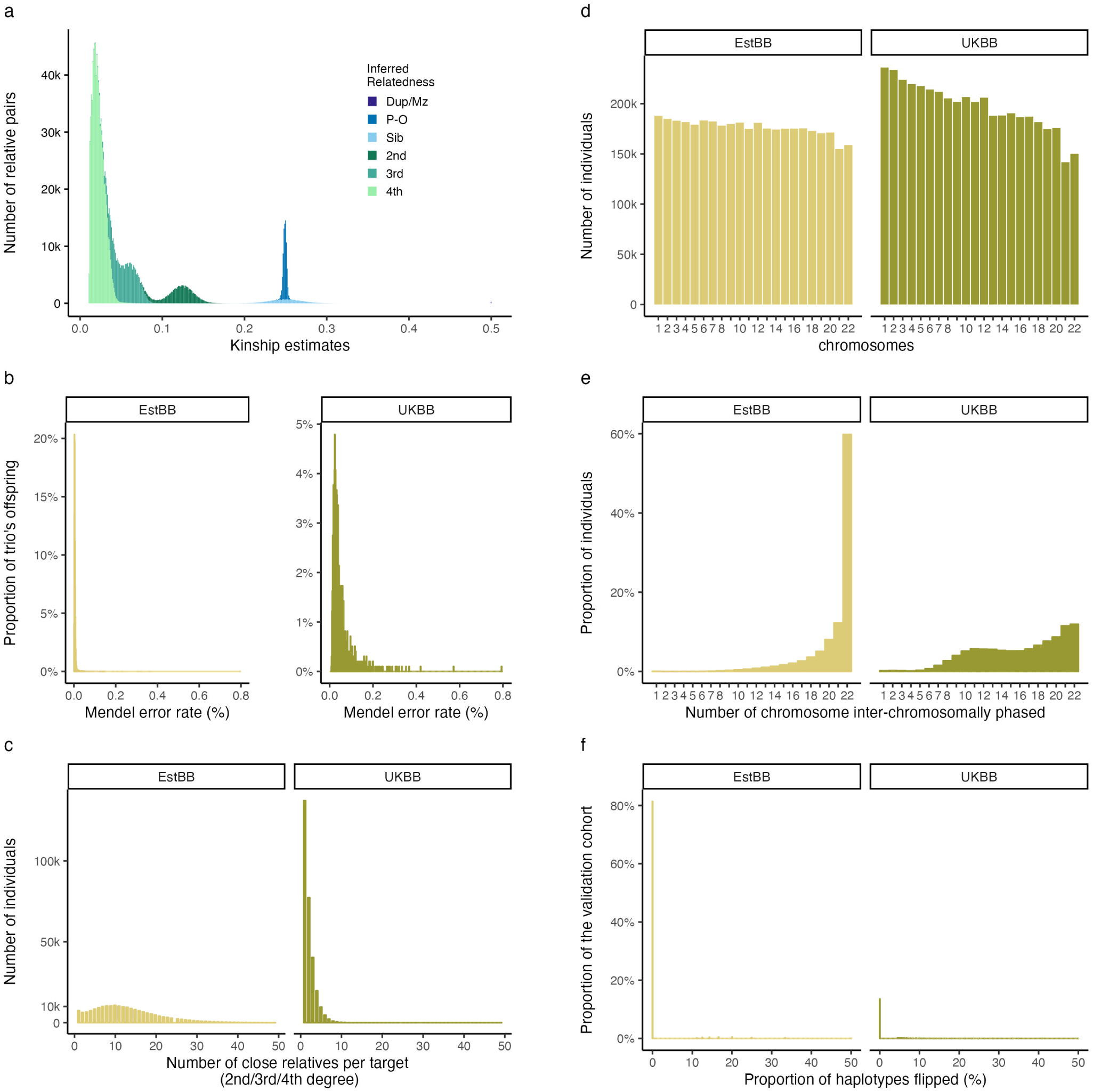
Overview of relatedness metrics and inter-chromosomal phasing in the Estonian Biobank (EstBB), with comparisons to the UK Biobank (UKBB). a) Distribution of inferred relatedness across the EstBB cohort. Dup/MZ=monozygotic twins; P-O=parent-offspring; Sib=sibling; 2nd=2nd degree relative pairs; 3rd=3rd degree relative pairs; 4th=4th degree relative pairs. Relatedness was inferred using the KING software v2.2.7^48^. **b**) Distribution of Mendel error in trio’s offspring for both the EstBB and UKBB cohorts. **c**) Distribution of the number of close relative per individual. **d**) Distribution of the number of individuals (y-axis) with a given chromosome (x-axis) successfully included in the inter-chromosomal phasing. **e**) Distribution of the number of individuals with N chromosomes (x-axis) successfully included in the inter-chromosomal phasing. **f**) Distribution of inter-chromosomal phasing error rate in the validation cohorts, calculated by comparing phasing results from surrogate parents to those obtained using parental genomes as the ground truth.

**Supplementary Fig. 23.**
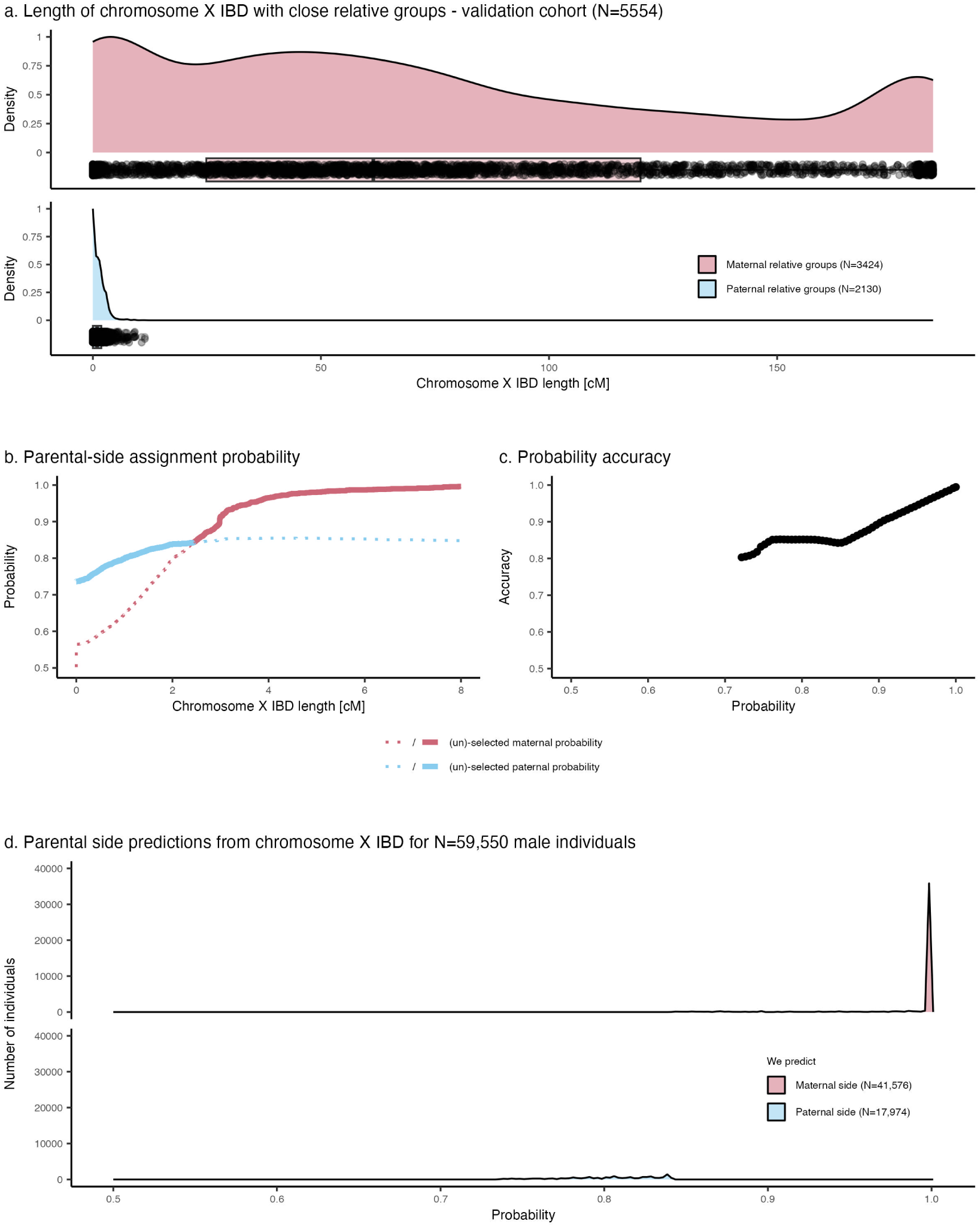
Parental side assignment from chromosome X IBD analysis in male individuals in the Estonian Biobank. **a**) Density distribution of chromosome X IBD segment length in centimorgan (cM) between targets and surrogate father (blue) and targets and surrogate mother (red) across the validation cohort’s male individuals. Boxes indicate the interquartile range (IQR), with the bottom and top of the box representing the 25th (Q1) and 75th (Q3) percentiles, respectively. The horizontal line within the box represents the median (50th percentile). Whiskers extend to the smallest and largest values within *Q*1*−*1.5 *× IQR* and *Q*3 + 1.5 *× IQR*. Each point represent a target-relative pair. **b**) Parental side probabilities depending on chromosome X IBD segment length derived from the validation cohort. Selected and unselected maternal (red) and paternal (blue) probabilities for a given x-axis value are indicated by solid and dotted lines, respectively. **c**) Accuracy of parental side predictions (y-axis) as a function of the parental side probability (x-axis). **d**) Distribution of maternal (red) and paternal (blue) side assignment probabilities derived chromosome X IBD across 59,550 Estonian Biobank male individuals.

**Supplementary Fig. 24.**
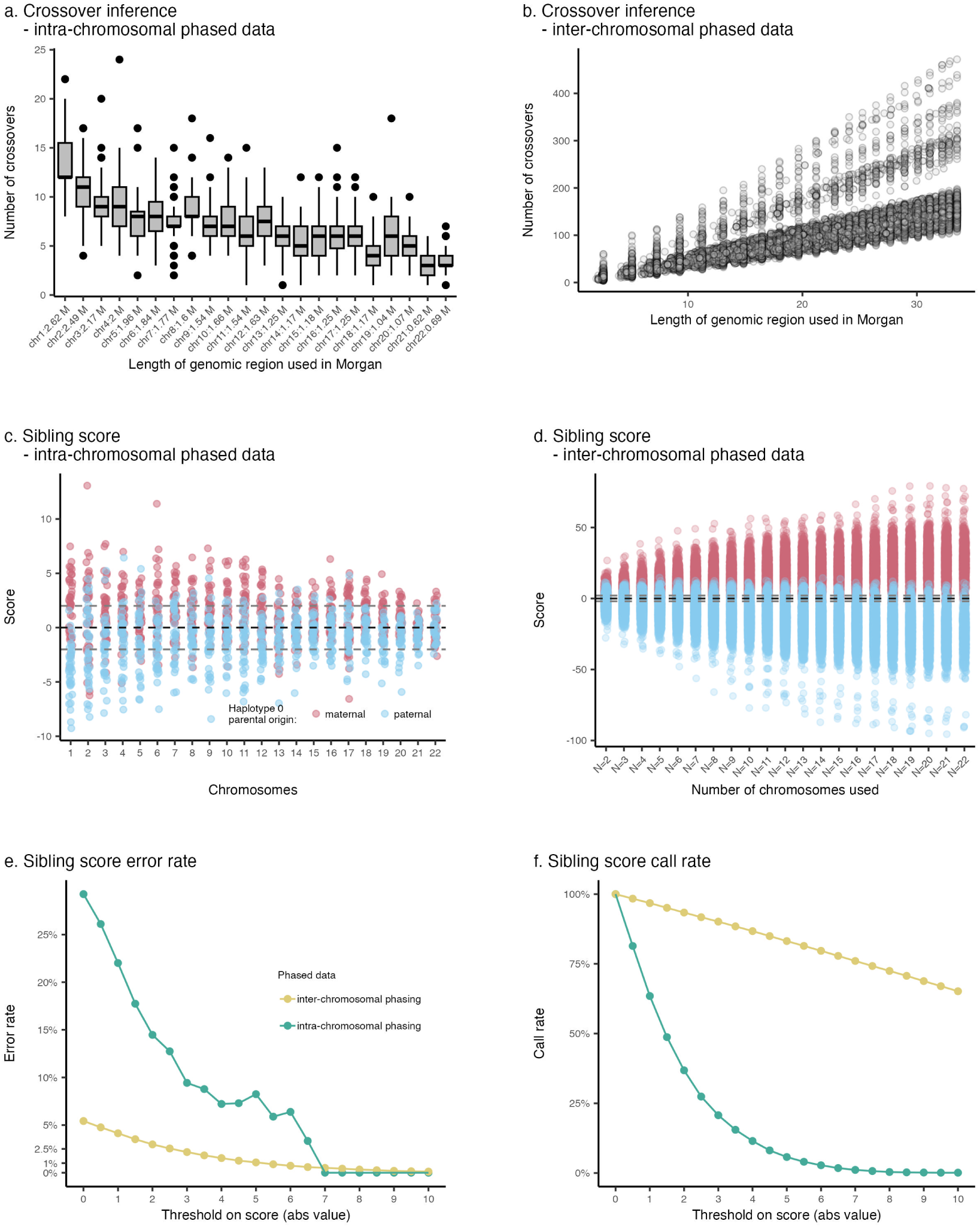
Evaluation of sibling scores from intra- and inter-chromosomal phasing in the Estonian Biobank validation cohort. a) Distribution of the number of inferred crossovers (y-axis) per chromosome (x-axis) derived from intra-chromosomally phased data. **b**) Distribution of the number of inferred crossovers (y-axis) relative to the genomic length used (in Morgans, x-axis) derived from inter-chromosomally phased data. **c**) Distribution of sibling scores (y-axis) per chromosome (x-axis) using intra-chromosomally phased data. Maternal and paternal haplotypes are color-coded in red and blue, respectively. **d**) Distribution of sibling scores (y-axis) relative to the number of chromosomes included in inter-chromosomally phased data (x-axis). **e**) Error rates (y-axis) of sibling scores for varying score thresholds (absolute values, x-axis). **f**) Call rates (y-axis) of sibling scores for varying score thresholds (absolute values, x-axis), showing the proportion of individuals meeting the threshold criteria.

**Supplementary Fig. 25.**
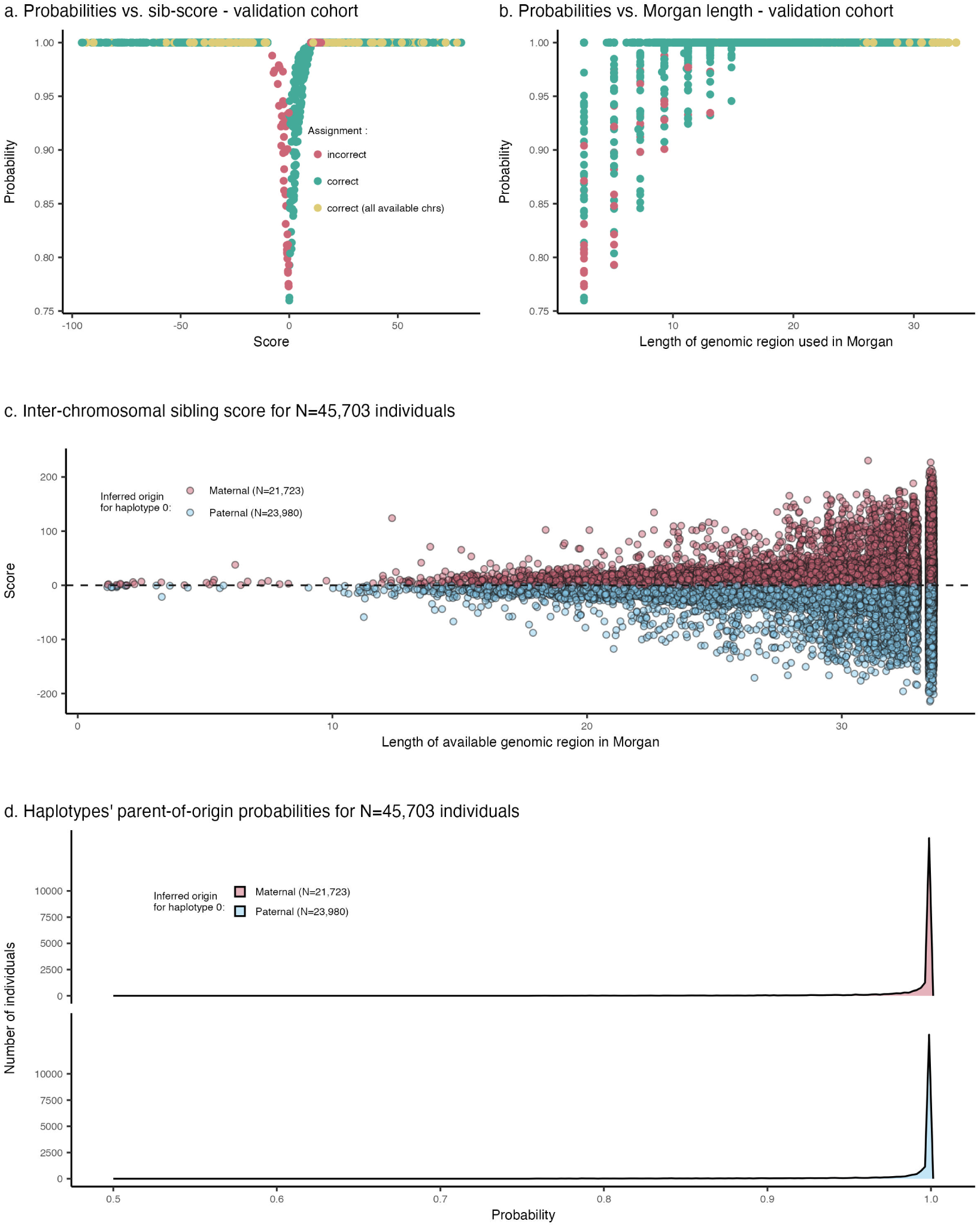
Validation and derivation of parent-of-origin probabilities using sibling scores from inter-chromosomal phased data in the Estonian Biobank. a) PofO probabilities (y-axis) as a function of sibling scores (x-axis) in the validation cohort. Each dot represents a specific configuration (an individual with a given number of chromosomes used). We varied the number of chromosome used per individual to assess the accuracy of different configurations. Red dots indicate incorrect parental assignments, green dots indicate correct assignments, and yellow dots denote correct assignments using the maximum available chromosomes (*N_max_*, corresponds to the full set of inter-chromosomally phased chromosome for a given individual). No errors were observed when *N_max_* chromosomes were used. **b**) PofO probabilities (y-axis) plotted against the total available genomic length in Morgans (x-axis) for sibling score calculation in the validation cohort. Each configuration is represented as a dot (color-coded as in panel a). Increasing genomic length results in higher assignment accuracy, with perfect accuracy achieved at *N_max_* (yellow dots). **c**) Sibling scores (y-axis) derived for N=45,703 individuals across available genomic lengths (x-axis). **d**) Distribution of PofO probabilities for 45,703 Estonian Biobank individuals.

**Supplementary Fig. 26.**
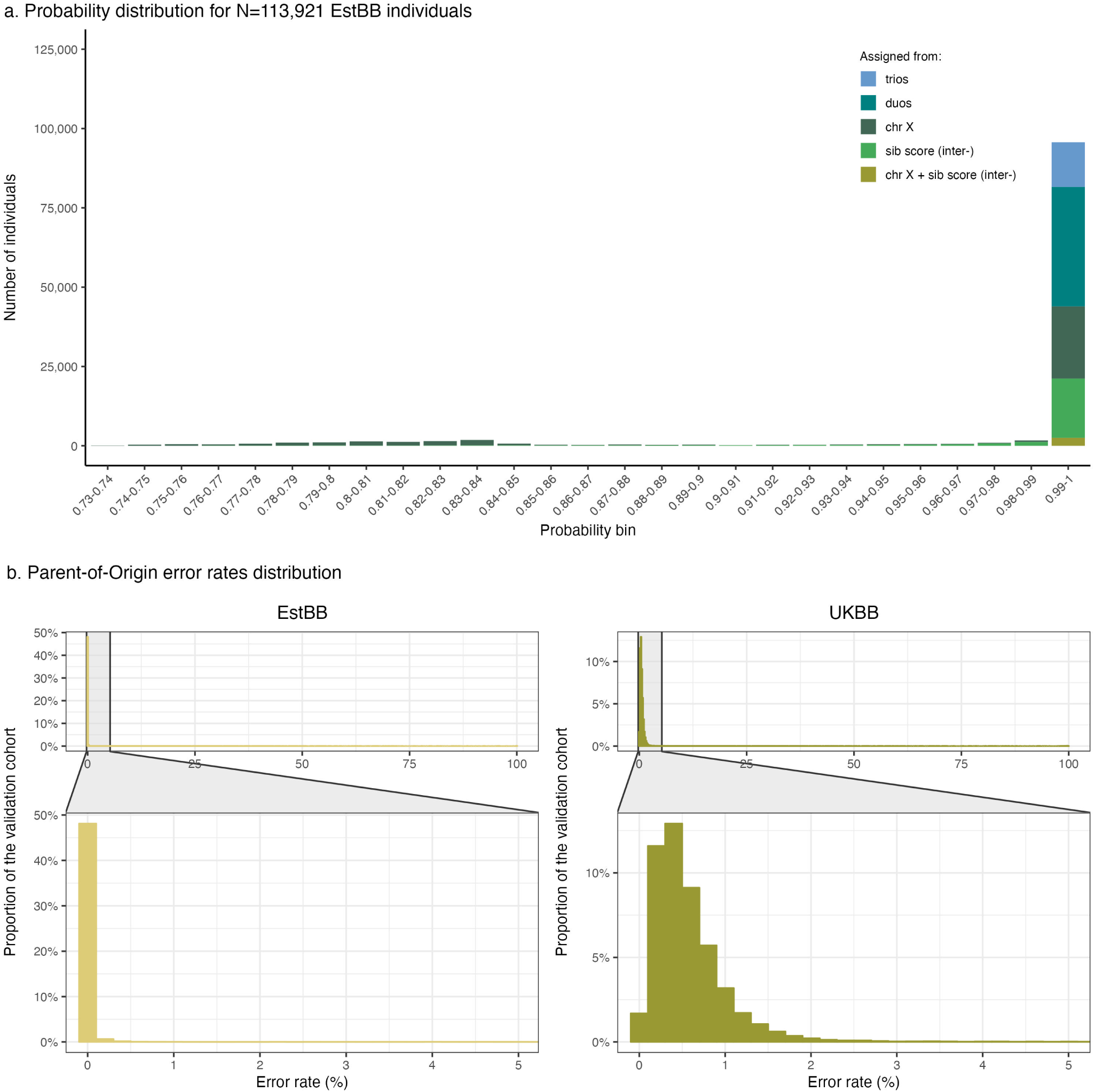
Parent-of-origin probability distribution and error rates in the Estonian Biobank. **a**) Distribution of PofO probabilities for N=113,921 individuals in the Estonian Biobank, color-coded by the source of inference (e.g., chromosome X data, sibling score from inter-chromosomal phased data, duos, and trios). Most individuals exhibit high PofO probabilities, reflecting the accuracy of the approach. **b**) Comparison of PofO error rate distributions between the Estonian Biobank (EstBB) and UK Biobank (UKBB) validation cohorts. The EstBB shows consistently lower error rates, likely attributed to the higher number of close relatives per individual, which enhances IBD detection and improves inter-chromosomal phasing accuracy. Zoomed panels show the error rates below 5%, highlighting the robustness of PofO inference in both biobanks.

## Supplementary Tables

Supplementary Tables can also be visualized online and downloaded via this link.

**Supplementary Table 1.** UK Biobank traits selected for this study. TRAIT: phenotype name; N: number of individuals (selected as (i) self-reported as white British, confirmed by PCA, according to UK Biobank field 22006; (ii) having a PofO probability *>* 0.99, as defined for the differential GWAS^3^). For type 2 diabetes, we indicated cases and controls. TS ratio: Relative leucocyte telomere length; SHBG: Sex Hormone-Binding Globulin.

| TRAIT | N | N males | N females |
| --- | --- | --- | --- |
| Telomere length | 109355 | 58552 | 50803 |
| Sodium in urine | 109372 | 58565 | 50807 |
| Creatinine | 109379 | 58568 | 50811 |
| Microalbumin in urine | 109379 | 58568 | 50811 |
| Vitamin D | 109385 | 58572 | 50813 |
| Urate | 109385 | 58572 | 50813 |
| Triglycerides | 109385 | 58572 | 50813 |
| Total protein | 109385 | 58572 | 50813 |
| Testosterone | 109385 | 58572 | 50813 |
| Total bilirubin | 109385 | 58572 | 50813 |
| SHBG | 109385 | 58572 | 50813 |
| Rheumatoid factor | 109385 | 58572 | 50813 |
| Phosphate | 109385 | 58572 | 50813 |
| Oestradiol | 109385 | 58572 | 50813 |
| Lipoprotein A | 109385 | 58572 | 50813 |
| LDL direct | 109385 | 58572 | 50813 |
| IGF-1 | 109385 | 58572 | 50813 |
| HDL cholesterol | 109385 | 58572 | 50813 |
| Glycated haemoglobin | 109385 | 58572 | 50813 |
| Glucose | 109385 | 58572 | 50813 |
| Gamma glutamyltransferase | 109385 | 58572 | 50813 |
| Cystatin C | 109385 | 58572 | 50813 |
| C-reactive protein | 109385 | 58572 | 50813 |
| Creatinine | 109385 | 58572 | 50813 |
| Cholesterol | 109385 | 58572 | 50813 |
| Calcium | 109385 | 58572 | 50813 |
| Urea | 109385 | 58572 | 50813 |
| Direct bilirubin | 109385 | 58572 | 50813 |
| Aspartate aminotransferase | 109385 | 58572 | 50813 |
| Apolipoprotein B | 109385 | 58572 | 50813 |
| Apolipoprotein A | 109385 | 58572 | 50813 |
| Alanine aminotransferase | 109385 | 58572 | 50813 |
| Alkaline phosphatase | 109385 | 58572 | 50813 |
| Albumin | 109385 | 58572 | 50813 |
| Reticulocyte count | 109385 | 58572 | 50813 |
| Nucleated red blood cell count | 109385 | 58572 | 50813 |
| Basophil count | 109385 | 58572 | 50813 |
| Eosinophil count | 109385 | 58572 | 50813 |
| Neutrophil count | 109385 | 58572 | 50813 |
| Monocyte count | 109385 | 58572 | 50813 |
| Lymphocyte count | 109385 | 58572 | 50813 |
| Platelet count | 109385 | 58572 | 50813 |
| Red blood cell | 109385 | 58572 | 50813 |
| White blood cell | 109385 | 58572 | 50813 |
| Standing height | 109385 | 58572 | 50813 |
| Hip circumference | 109385 | 58572 | 50813 |
| Waist circumference | 109385 | 58572 | 50813 |
| Trunk fat-free mass | 109385 | 58572 | 50813 |
| Trunk fat percentage | 109385 | 58572 | 50813 |
| Arm fat-free mass | 109385 | 58572 | 50813 |
| Arm fat percentage | 109385 | 58572 | 50813 |
| Leg fat-free mass | 109385 | 58572 | 50813 |
| Leg fat percentage | 109385 | 58572 | 50813 |
| Basal metabolic rate | 109385 | 58572 | 50813 |
| Whole body water mass | 109385 | 58572 | 50813 |
| Body fat percentage | 109385 | 58572 | 50813 |
| Body mass index | 109385 | 58572 | 50813 |
| Birth weight | 109385 | 58572 | 50813 |
| Type 2 diabetes | 9189 (99007) | 6265 (51513) | 2925 (47495) |

**Supplementary Table 2.**
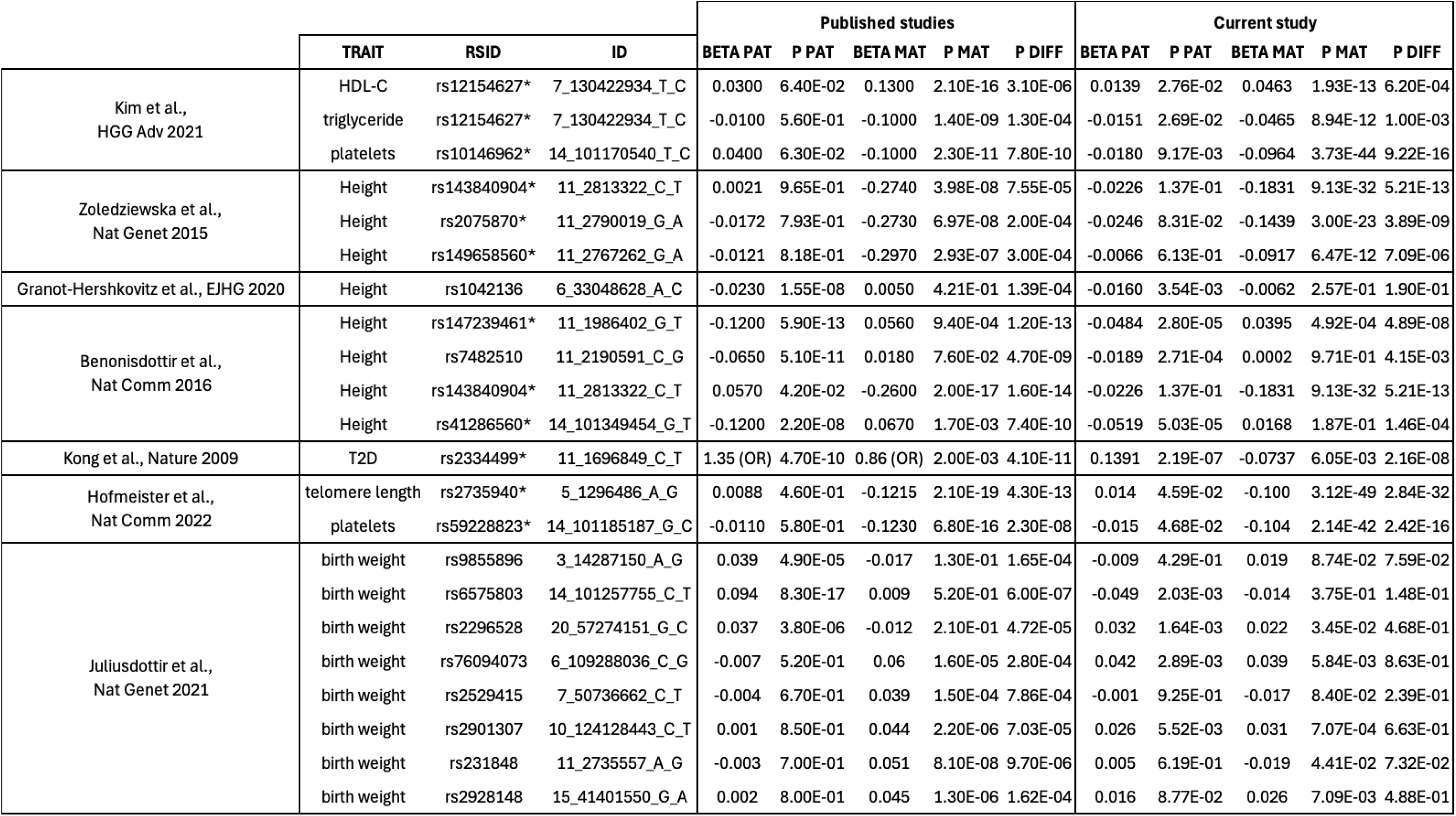
Replication of previously published Parent-of-origin effects. TRAIT: phenotype name; RSID: variant rs id; ID: chromosome position(hg19) reference allele alternative allele; BETA and P denote effect sizes and P-values; PAT, MAT, and DIFF denote paternal, maternal and differential tests; *: associations replicated in our study.

**Supplementary Table 3.** Imprinted regions. Imprinted regions extracted from Kong *et al.*^5^ and lifted over to hg19.

| Chromosome | Region start | Region end | Genes overlapping region |
| --- | --- | --- | --- |
| chr1 | 68011644 | 69016459 | ARHI |
| chr2 | 80015483 | 81031487 | LRRTM1 |
| chr4 | 89117065 | 90118980 | NAPIL5 |
| chr6 | 56682421 | 57691161 | PRIM2 |
| chr6 | 143761436 | 144885734 | PLAGL1,HYMAI |
| chr6 | 159890130 | 161376014 | IGF2R,SLC22A2,SLC22A3 |
| chr7 | 50154712 | 51361157 | GRB10 |
| chr7 | 93714535 | 95425726 | SGCE,PEG10,PPP1R9A |
| chr7 | 96149701 | 97154142 | DLX5 |
| chr7 | 129432993 | 130918860 | CPA4,MEST,KLF14 |
| chr8 | 949568 | 2156642 | DLGAP2 |
| chr8 | 140113081 | 141215299 | KCNK9 |
| chr11 | 1516405 | 3686539 | H19,IGF2,IGF2AS,INS,KCNQ1,KCNQ1DN,CDKN1C,SLC22A1LS,SLC22A1L,PHLDA2,OSBPL5 |
| chr11 | 5911654 | 7479277 | SMPD1,ZNF215 |
| chr11 | 31909324 | 32957086 | WT1 |
| chr14 | 100693252 | 101827367 | DLK1,MEG3 |
| chr15 | 23310105 | 24432449 | MKRN3,MAGEL2,NDN |
| chr15 | 24700069 | 26608348 | SNURF,IPW,UBE3A,ATP10A |
| chr16 | 2986109 | 3993490 | ZNF597 |
| chr18 | 44054572 | 45056448 | TCEB3C |
| chr19 | 56785919 | 57852075 | ZIM2,PEG3 |
| chr20 | 35649606 | 36652091 | NNAT |
| chr20 | 41643052 | 42670534 | L3MBTL |
| chr20 | 56914783 | 57986249 | NESP55,GNAS1 |

**Supplementary Table 4.** Replication of standing height association with sitting height. CHR: chromosome; POS: genetic position (hg19); SNP ID: variant rs id; A0: reference allele; A1: assessed allele; A1FREQ: A1 allele frequency. BETA, SE and P denote effect sizes, standard errors and P-values; PAT, MAT, DIFF and ADD denote paternal, maternal, differential and additive tests. TRAIT: phenotype name.

| CHR | POS | SNP ID | A0 | A1 | A1FREQ | BETA PAT | SE PAT | P PAT | BETA MAT | SE MAT | P MAT | BETA DIFF | SE DIFF | P DIFF | BETA ADD | SE ADD | P ADD | TRAIT |
| --- | --- | --- | --- | --- | --- | --- | --- | --- | --- | --- | --- | --- | --- | --- | --- | --- | --- | --- |
| 11 | 1918083 | rs7105710 | T | C | 0.546 | 0.001 | 0.005 | 8.18E-01 | -0.013 | 0.005 | 8.00E-03 | 0.014 | 0.007 | 4.10E-02 | -0.006 | 0.003 | 8.70E-02 | Sitting height |
| 11 | 2040272 | rs77708343 | A | G | 0.039 | -0.055 | 0.012 | 8.51E-06 | 0.046 | 0.012 | 1.21E-04 | -0.101 | 0.017 | 4.17E-09 | -0.003 | 0.009 | 6.95E-01 | Sitting height |
| 11 | 2813322 | rs143840904 | C | T | 0.019 | -0.044 | 0.018 | 1.30E-02 | -0.131 | 0.018 | 6.61E-13 | 0.087 | 0.025 | 3.80E-04 | -0.086 | 0.013 | 1.06E-11 | Sitting height |

**Supplementary Table 5.**
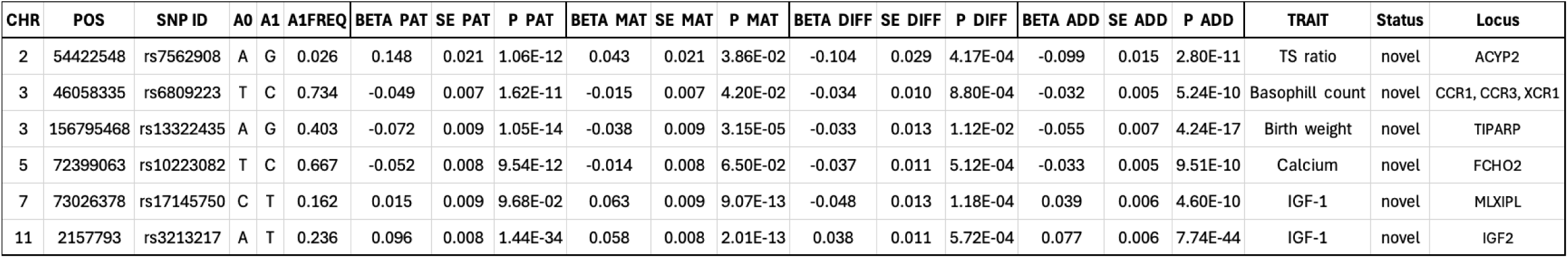
Parent-of-origin effect within additively associated regions, per phenotype. CHR: chromosome; POS: genetic position (hg19); SNP ID: variant rs id; A0: reference allele; A1: assessed allele; A1FREQ: A1 allele frequency. BETA, SE and P denote effect sizes, standard errors and P-values; PAT, MAT, DIFF and ADD denote paternal, maternal, differential and additive tests. TRAIT: phenotype name.

| CHR | POS | SNP ID | A0 | A1 | A1FREQ | BETA PAT | SE PAT | P PAT | BETA MAT | SE MAT | P MAT | BETA DIFF | SE DIFF | P DIFF | BETA ADD | SE ADD | P ADD | TRAIT | Status | Locus |
| --- | --- | --- | --- | --- | --- | --- | --- | --- | --- | --- | --- | --- | --- | --- | --- | --- | --- | --- | --- | --- |
| 2 | 54422548 | rs7562908 | A | G | 0.026 | 0.148 | 0.021 | 1.06E-12 | 0.043 | 0.021 | 3.86E-02 | -0.104 | 0.029 | 4.17E-04 | -0.099 | 0.015 | 2.80E-11 | TS ratio | novel | ACYP2 |
| 3 | 46058335 | rs6809223 | T | C | 0.734 | -0.049 | 0.007 | 1.62E-11 | -0.015 | 0.007 | 4.20E-02 | -0.034 | 0.010 | 8.80E-04 | -0.032 | 0.005 | 5.24E-10 | Basophil count | novel | CCR1, CCR3, XCR1 |
| 3 | 156795468 | rs13322435 | A | G | 0.403 | -0.072 | 0.009 | 1.05E-14 | -0.038 | 0.009 | 3.15E-05 | -0.033 | 0.013 | 1.12E-02 | -0.055 | 0.007 | 4.24E-17 | Birth weight | novel | TIPARP |
| 5 | 72399063 | rs10223082 | T | C | 0.667 | -0.052 | 0.008 | 9.54E-12 | -0.014 | 0.008 | 6.50E-02 | -0.037 | 0.011 | 5.12E-04 | -0.033 | 0.005 | 9.51E-10 | Calcium | novel | FCHO2 |
| 7 | 73026378 | rs17145750 | C | T | 0.162 | 0.015 | 0.009 | 9.68E-02 | 0.063 | 0.009 | 9.07E-13 | -0.048 | 0.013 | 1.18E-04 | 0.039 | 0.006 | 4.60E-10 | IGF-1 | novel | MLXIPL |
| 11 | 2157793 | rs3213217 | A | T | 0.236 | 0.096 | 0.008 | 1.44E-34 | 0.058 | 0.008 | 2.01E-13 | 0.038 | 0.011 | 5.72E-04 | 0.077 | 0.006 | 7.74E-44 | IGF-1 | novel | IGF2 |

**Supplementary Table 6.**
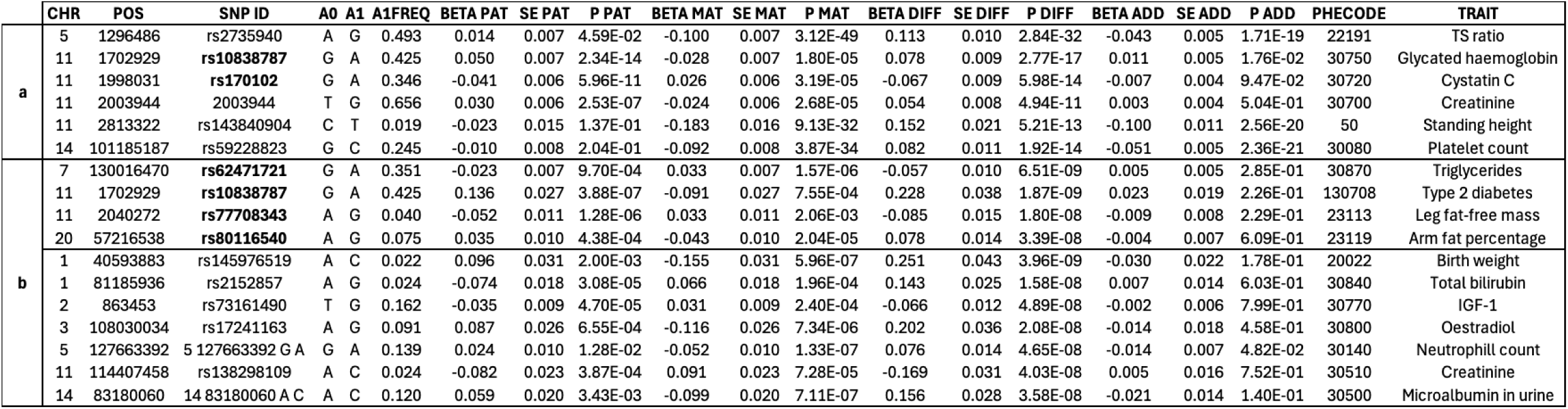
Parent-of-origin effect genome-wide. CHR: chromosome; POS: genetic position (hg19); SNP ID: variant rs id; A0: reference allele; A1: assessed allele; A1FREQ: A1 allele frequency. BETA, SE and P denote effect sizes, standard errors and P-values; PAT, MAT, DIFF and ADD denote paternal, maternal, differential and additive tests. TRAIT: phenotype name. **a**) POEs robust when correcting for the number of traits tested (*P_D_ <* 1 *×* 10*^−^*^9^). **b**) POEs genome-wide significant at the single trait level (1 *×* 10*^−^*^9^ *< P_D_ <* 5 *×* 10*^−^*^8^).

**Supplementary Table 7.**
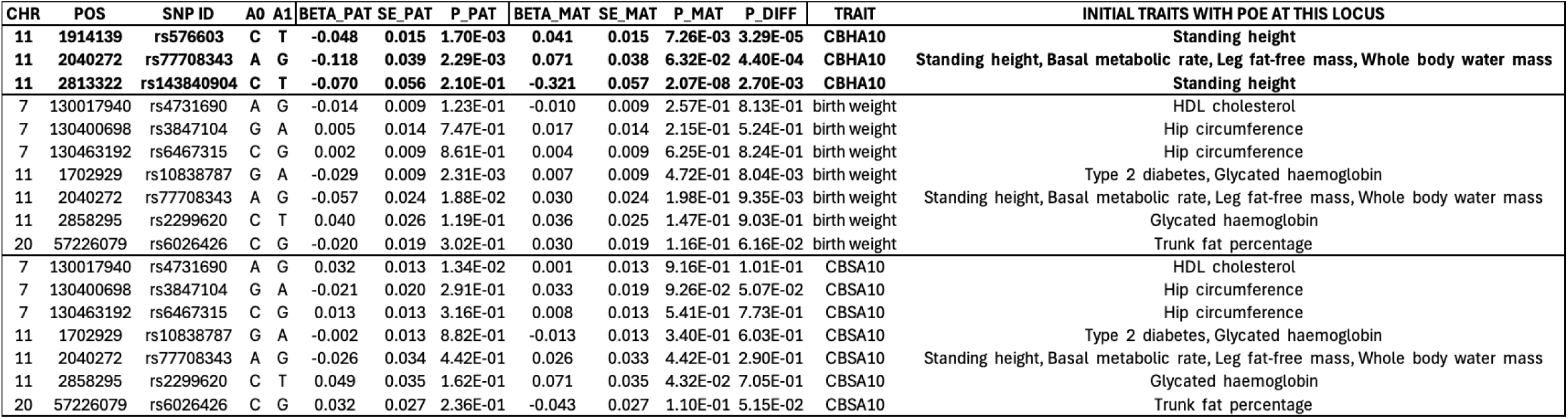
Replication of associations with early childhood traits in the UK Biobank cohort. CHR: chromosome; POS: genetic position (hg19); SNP ID: variant rs id; A0: reference allele; A1: assessed allele; A1FREQ: A1 allele frequency. BETA, SE and P denote effect sizes, standard errors and P-values; PAT, MAT, DIFF and ADD denote paternal, maternal, differential and additive tests. TRAIT: phenotype name. Bold indicate significant POEs; CBHA10: comparative body height at age 10; CBSA10: comparative body size at age 10.

| CHR | POS | SNP ID | A0 | A1 | BETA_PAT | SE_PAT | P_PAT | BETA_MAT | SE_MAT | P_MAT | P_DIFF | TRAIT | INITIAL TRAITS WITH POE AT THIS LOCUS |
| --- | --- | --- | --- | --- | --- | --- | --- | --- | --- | --- | --- | --- | --- |
| **11** | **1914139** | **rs576603** | **C** | **T** | **-0.048** | **0.015** | **1.70E-03** | **0.041** | **0.015** | **7.26E-03** | **3.29E-05** | **CBHA10** | **Standing height** |
| **11** | **2040272** | **rs77708343** | **A** | **G** | **-0.118** | **0.039** | **2.29E-03** | **0.071** | **0.038** | **6.32E-02** | **4.40E-04** | **CBHA10** | **Standing height, Basal metabolic rate, Leg fat-free mass, Whole body water mass** |
| **11** | **2813322** | **rs143840904** | **C** | **T** | **-0.070** | **0.056** | **2.10E-01** | **-0.321** | **0.057** | **2.07E-08** | **2.70E-03** | **CBHA10** | **Standing height** |
| 7 | 130017940 | rs4731690 | A | G | -0.014 | 0.009 | 1.23E-01 | -0.010 | 0.009 | 2.57E-01 | 8.13E-01 | birth weight | HDL cholesterol |
| 7 | 130400698 | rs3847104 | G | A | 0.005 | 0.014 | 7.47E-01 | 0.017 | 0.014 | 2.15E-01 | 5.24E-01 | birth weight | Hip circumference |
| 7 | 130463192 | rs6467315 | C | G | 0.002 | 0.009 | 8.61E-01 | 0.004 | 0.009 | 6.25E-01 | 8.24E-01 | birth weight | Hip circumference |
| 11 | 1702929 | rs10838787 | G | A | -0.029 | 0.009 | 2.31E-03 | 0.007 | 0.009 | 4.72E-01 | 8.04E-03 | birth weight | Type 2 diabetes, Glycated haemoglobin |
| 11 | 2040272 | rs77708343 | A | G | -0.057 | 0.024 | 1.88E-02 | 0.030 | 0.024 | 1.98E-01 | 9.35E-03 | birth weight | Standing height, Basal metabolic rate, Leg fat-free mass, Whole body water mass |
| 11 | 2858295 | rs2299620 | C | T | 0.040 | 0.026 | 1.19E-01 | 0.036 | 0.025 | 1.47E-01 | 9.03E-01 | birth weight | Glycated haemoglobin |
| 20 | 57226079 | rs6026426 | C | G | -0.020 | 0.019 | 3.02E-01 | 0.030 | 0.019 | 1.16E-01 | 6.16E-02 | birth weight | Trunk fat percentage |
| 7 | 130017940 | rs4731690 | A | G | 0.032 | 0.013 | 1.34E-02 | 0.001 | 0.013 | 9.16E-01 | 1.01E-01 | CBSA10 | HDL cholesterol |
| 7 | 130400698 | rs3847104 | G | A | -0.021 | 0.020 | 2.91E-01 | 0.033 | 0.019 | 9.26E-02 | 5.07E-02 | CBSA10 | Hip circumference |
| 7 | 130463192 | rs6467315 | C | G | 0.013 | 0.013 | 3.16E-01 | 0.008 | 0.013 | 5.41E-01 | 7.73E-01 | CBSA10 | Hip circumference |
| 11 | 1702929 | rs10838787 | G | A | -0.002 | 0.013 | 8.82E-01 | -0.013 | 0.013 | 3.40E-01 | 6.03E-01 | CBSA10 | Type 2 diabetes, Glycated haemoglobin |
| 11 | 2040272 | rs77708343 | A | G | -0.026 | 0.034 | 4.42E-01 | 0.026 | 0.033 | 4.42E-01 | 2.90E-01 | CBSA10 | Standing height, Basal metabolic rate, Leg fat-free mass, Whole body water mass |
| 11 | 2858295 | rs2299620 | C | T | 0.049 | 0.035 | 1.62E-01 | 0.071 | 0.035 | 4.32E-02 | 7.05E-01 | CBSA10 | Glycated haemoglobin |
| 20 | 57226079 | rs6026426 | C | G | 0.032 | 0.027 | 2.36E-01 | -0.043 | 0.027 | 1.10E-01 | 5.15E-02 | CBSA10 | Trunk fat percentage |

**Supplementary Table 8.** Number of individuals per BMI time points in the MoBa cohort.

| BMI time point | N |
| --- | --- |
| BMI at 6 Weeks | 29791 |
| BMI at 3 Months | 41865 |
| BMI at 6 Months | 42346 |
| BMI at 8 Months | 36790 |
| BMI at 1 Year | 36721 |
| BMI at 16 Months | 27595 |
| BMI at 2 Years | 27435 |
| BMI at 3 Years | 27985 |
| BMI at 5 Years | 22667 |
| BMI at 7 Years | 23280 |
| BMI at 8 Years | 17888 |

**Supplementary Table 9.** Replication of associations with infancy and childhood BMI in the MoBa cohort. SNP ID: variant rs id; BETA, SE and P denote effect sizes, standard errors and P-values; PAT, MAT and MAT NT denote paternal transmitted, maternal transmitted, and maternal untransmitted alleles association coefficients; P DIFF: Z-score P-value for the differential effect between paternal and maternal transmitted alleles; P DIFF MAT: Z-score P-value for the differential effect between maternal transmitted and maternal untransmitted alleles. Bold P DIFF denote significant POEs.

| ID | BMI timepoints | BETA PAT | SE PAT | P PAT | BETA MAT | SE MAT | P MAT | BETA MAT NT | SE MAT NT | P MAT NT | P DIFF | P DIFF MAT | INITIAL TRAITS WITH POE AT THIS LOCUS |
| --- | --- | --- | --- | --- | --- | --- | --- | --- | --- | --- | --- | --- | --- |
| rs6467315 | 6 weeks | -0.019 | 0.015 | 2.04E-01 | 0.071 | 0.015 | 1.26E-06 | 0.028 | 0.014 | 4.74E-02 | **1.51E-05** | 3.47E-02 | Hip circumference |
|  | 3 months | 0.001 | 0.012 | 9.62E-01 | 0.085 | 0.012 | 9.46E-12 | 0.003 | 0.012 | 7.74E-01 | **1.68E-06** | 2.50E-06 |  |
|  | 6 months | 0.003 | 0.012 | 8.12E-01 | 0.089 | 0.012 | 7.72E-13 | 0.016 | 0.012 | 1.96E-01 | **9.43E-07** | 2.02E-05 |  |
|  | 8 months | 0.012 | 0.013 | 3.75E-01 | 0.087 | 0.013 | 6.15E-11 | 0.009 | 0.013 | 4.97E-01 | **6.30E-05** | 2.23E-05 |  |
|  | 12 months | -0.007 | 0.013 | 5.75E-01 | 0.069 | 0.013 | 2.20E-07 | 0.000 | 0.013 | 9.88E-01 | **4.85E-05** | 1.82E-04 |  |
|  | 16 months | -0.011 | 0.015 | 4.51E-01 | 0.066 | 0.015 | 1.24E-05 | 0.011 | 0.015 | 4.39E-01 | **2.90E-04** | 9.05E-03 |  |
|  | 2 years | -0.017 | 0.015 | 2.67E-01 | 0.057 | 0.015 | 1.81E-04 | 0.017 | 0.015 | 2.47E-01 | **5.95E-04** | 5.81E-02 |  |
|  | 3 years | 0.012 | 0.015 | 4.17E-01 | 0.058 | 0.015 | 9.60E-05 | 0.003 | 0.014 | 8.61E-01 | 2.89E-02 | 7.20E-03 |  |
|  | 5 years | -0.013 | 0.016 | 4.38E-01 | 0.031 | 0.016 | 5.77E-02 | -0.013 | 0.016 | 4.09E-01 | 5.86E-02 | 5.24E-02 |  |
|  | 7 years | -0.023 | 0.016 | 1.54E-01 | 0.030 | 0.016 | 6.63E-02 | -0.008 | 0.016 | 6.24E-01 | 2.10E-02 | 9.68E-02 |  |
|  | 8 years | -0.004 | 0.019 | 8.43E-01 | 0.026 | 0.019 | 1.69E-01 | 0.024 | 0.018 | 1.81E-01 | 2.66E-01 | 9.48E-01 |  |
| rs10838787 | 6 weeks | -0.006 | 0.015 | 6.76E-01 | 0.028 | 0.015 | 5.61E-02 | 0.016 | 0.014 | 2.57E-01 | 9.96E-02 | 5.51E-01 | Type 2 diabetes, Glycated haemoglobin, Glucose |
|  | 3 months | -0.020 | 0.013 | 1.16E-01 | 0.019 | 0.013 | 1.28E-01 | 0.025 | 0.012 | 3.68E-02 | 2.88E-02 | 7.28E-01 |  |
|  | 6 months | -0.028 | 0.013 | 2.75E-02 | 0.017 | 0.013 | 1.82E-01 | 0.024 | 0.012 | 4.81E-02 | 1.23E-02 | 6.86E-01 |  |
|  | 8 months | -0.046 | 0.013 | 6.09E-04 | 0.011 | 0.013 | 4.20E-01 | 0.030 | 0.013 | 2.12E-02 | 2.74E-03 | 3.12E-01 |  |
|  | 12 months | -0.042 | 0.013 | 1.96E-03 | 0.009 | 0.013 | 5.22E-01 | 0.030 | 0.013 | 1.83E-02 | 8.24E-03 | 2.42E-01 |  |
|  | 16 months | -0.044 | 0.015 | 3.58E-03 | 0.008 | 0.015 | 5.91E-01 | 0.007 | 0.015 | 6.52E-01 | 1.47E-02 | 9.40E-01 |  |
|  | 2 years | -0.029 | 0.015 | 5.51E-02 | 0.003 | 0.015 | 8.63E-01 | -0.001 | 0.015 | 9.56E-01 | 1.39E-01 | 8.70E-01 |  |
|  | 3 years | -0.038 | 0.015 | 1.15E-02 | -0.002 | 0.015 | 8.84E-01 | 0.015 | 0.014 | 2.92E-01 | 9.21E-02 | 4.04E-01 |  |
|  | 5 years | -0.021 | 0.016 | 2.11E-01 | -0.010 | 0.016 | 5.37E-01 | 0.013 | 0.016 | 4.12E-01 | 6.54E-01 | 3.11E-01 |  |
|  | 7 years | -0.018 | 0.017 | 2.71E-01 | -0.004 | 0.017 | 8.24E-01 | 0.024 | 0.016 | 1.32E-01 | 5.35E-01 | 2.29E-01 |  |
|  | 8 years | -0.001 | 0.019 | 9.76E-01 | -0.007 | 0.019 | 6.91E-01 | 0.036 | 0.018 | 4.67E-02 | 7.95E-01 | 9.61E-02 |  |
| rs2299620 | 6 weeks | -0.040 | 0.040 | 3.17E-01 | -0.025 | -0.040 | 5.36E-01 | 0.013 | 0.037 | 7.30E-01 | 2.53E-01 | 4.91E-01 | Glycated haemoglobin |
|  | 3 months | 0.017 | 0.034 | 6.17E-01 | -0.045 | -0.034 | 1.79E-01 | 0.034 | 0.032 | 2.92E-01 | 5.60E-01 | 8.92E-02 |  |
|  | 6 months | 0.043 | 0.034 | 2.13E-01 | -0.015 | -0.034 | 6.67E-01 | -0.013 | 0.032 | 6.96E-01 | 5.59E-01 | 6.86E-01 |  |
|  | 8 months | 0.044 | 0.037 | 2.35E-01 | 0.016 | -0.036 | 6.62E-01 | 0.007 | 0.034 | 8.36E-01 | 2.47E-01 | 8.62E-01 |  |
|  | 12 months | 0.021 | 0.037 | 5.71E-01 | 0.034 | -0.036 | 3.41E-01 | 0.010 | 0.034 | 7.81E-01 | 2.85E-01 | 6.20E-01 |  |
|  | 16 months | 0.030 | 0.042 | 4.71E-01 | 0.050 | -0.041 | 2.22E-01 | -0.015 | 0.039 | 7.05E-01 | 1.71E-01 | 2.51E-01 |  |
|  | 2 years | 0.036 | 0.042 | 3.83E-01 | 0.035 | -0.041 | 3.92E-01 | 0.046 | 0.039 | 2.35E-01 | 2.21E-01 | 8.55E-01 |  |
|  | 3 years | 0.008 | 0.041 | 8.49E-01 | 0.055 | -0.041 | 1.78E-01 | 0.019 | 0.038 | 6.22E-01 | 2.80E-01 | 5.18E-01 |  |
|  | 5 years | -0.004 | 0.045 | 9.32E-01 | 0.020 | -0.044 | 6.47E-01 | 0.012 | 0.041 | 7.79E-01 | 7.94E-01 | 8.87E-01 |  |
|  | 7 years | 0.066 | 0.045 | 1.45E-01 | 0.082 | -0.045 | 6.65E-02 | 0.005 | 0.042 | 8.96E-01 | 1.99E-02 | 2.10E-01 |  |
|  | 8 years | -0.007 | 0.051 | 8.88E-01 | 0.074 | -0.050 | 1.40E-01 | 0.011 | 0.047 | 8.13E-01 | 3.53E-01 | 3.61E-01 |  |
| rs4731690 | 6 weeks | 0.008 | 0.015 | 5.82E-01 | 0.025 | 0.015 | 8.35E-02 | 0.016 | 0.014 | 2.52E-01 | 4.04E-01 | 6.49E-01 | HDL cholesterol, Triglycerides, SHBG |
|  | 3 months | -0.001 | 0.012 | 9.14E-01 | 0.034 | 0.012 | 6.90E-03 | 0.019 | 0.012 | 1.19E-01 | 4.70E-02 | 3.88E-01 |  |
|  | 6 months | 0.006 | 0.012 | 6.12E-01 | 0.039 | 0.012 | 1.72E-03 | 0.007 | 0.012 | 5.86E-01 | 6.33E-02 | 6.04E-02 |  |
|  | 8 months | 0.008 | 0.013 | 5.41E-01 | 0.031 | 0.013 | 2.10E-02 | 0.004 | 0.013 | 7.39E-01 | 2.30E-01 | 1.53E-01 |  |
|  | 12 months | 0.004 | 0.013 | 7.40E-01 | 0.037 | 0.013 | 6.00E-03 | -0.002 | 0.013 | 8.59E-01 | 8.76E-02 | 3.56E-02 |  |
|  | 16 months | 0.017 | 0.015 | 2.58E-01 | 0.010 | 0.015 | 4.99E-01 | -0.002 | 0.015 | 9.18E-01 | 7.48E-01 | 5.76E-01 |  |
|  | 2 years | 0.036 | 0.015 | 1.87E-02 | 0.005 | 0.015 | 7.39E-01 | -0.007 | 0.015 | 6.41E-01 | 1.53E-01 | 5.73E-01 |  |
|  | 3 years | 0.021 | 0.015 | 1.57E-01 | -0.007 | 0.015 | 6.18E-01 | -0.019 | 0.014 | 1.85E-01 | 1.76E-01 | 5.75E-01 |  |
|  | 5 years | 0.041 | 0.016 | 1.23E-02 | -0.012 | 0.016 | 4.69E-01 | -0.002 | 0.016 | 8.89E-01 | 2.25E-02 | 6.71E-01 |  |
|  | 7 years | -0.003 | 0.016 | 8.60E-01 | 0.015 | 0.016 | 3.54E-01 | -0.020 | 0.016 | 2.07E-01 | 4.35E-01 | 1.23E-01 |  |
|  | 8 years | 0.019 | 0.019 | 3.17E-01 | 0.007 | 0.019 | 7.13E-01 | -0.042 | 0.018 | 1.86E-02 | 6.55E-01 | 5.77E-02 |  |
| rs6026426 | 6 weeks | -0.069 | -0.028 | 1.20E-02 | 0.021 | 0.027 | 4.39E-01 | 0.006 | 0.026 | 8.33E-01 | 2.06E-01 | 4.84E-01 | Trunk fat percentage, Arm, Leg and Body fat percentages |
|  | 3 months | -0.059 | -0.023 | 1.06E-02 | -0.006 | 0.023 | 7.95E-01 | -0.012 | 0.022 | 5.90E-01 | 4.49E-02 | 5.74E-01 |  |
|  | 6 months | -0.063 | -0.023 | 6.68E-03 | -0.005 | 0.023 | 8.37E-01 | -0.026 | 0.022 | 2.40E-01 | 3.76E-02 | 3.33E-01 |  |
|  | 8 months | -0.055 | -0.025 | 2.57E-02 | -0.008 | 0.024 | 7.39E-01 | -0.034 | 0.024 | 1.54E-01 | 6.79E-02 | 2.18E-01 |  |
|  | 12 months | -0.058 | -0.025 | 1.90E-02 | -0.024 | 0.024 | 3.32E-01 | -0.026 | 0.023 | 2.76E-01 | 1.85E-02 | 1.46E-01 |  |
|  | 16 months | -0.035 | -0.028 | 2.21E-01 | -0.034 | 0.027 | 2.08E-01 | -0.028 | 0.027 | 3.03E-01 | 7.92E-02 | 1.05E-01 |  |
|  | 2 years | -0.043 | -0.028 | 1.29E-01 | -0.020 | 0.027 | 4.73E-01 | -0.002 | 0.027 | 9.50E-01 | 1.12E-01 | 5.77E-01 |  |
|  | 3 years | -0.005 | -0.028 | 8.60E-01 | -0.036 | 0.027 | 1.76E-01 | -0.002 | 0.026 | 9.37E-01 | 2.86E-01 | 3.05E-01 |  |
|  | 5 years | -0.065 | -0.031 | 3.29E-02 | -0.030 | 0.029 | 3.05E-01 | -0.041 | 0.029 | 1.57E-01 | 2.44E-02 | 8.46E-02 |  |
|  | 7 years | -0.063 | -0.031 | 3.87E-02 | -0.013 | 0.030 | 6.61E-01 | -0.013 | 0.029 | 6.47E-01 | 7.40E-02 | 5.26E-01 |  |
|  | 8 years | -0.033 | -0.035 | 3.39E-01 | -0.006 | 0.033 | 8.50E-01 | -0.038 | 0.033 | 2.43E-01 | 4.11E-01 | 3.42E-01 |  |
| rs77708343 | 6 weeks | -0.004 | 0.031 | 8.97E-01 | 0.057 | 0.031 | 6.87E-02 | 0.017 | 0.030 | 5.61E-01 | 1.68E-01 | 3.62E-01 | Standing height, Basal metabolic rate, Leg fat-free mass |
|  | 3 months | -0.055 | 0.027 | 3.88E-02 | 0.021 | 0.027 | 4.24E-01 | 0.023 | 0.026 | 3.58E-01 | 4.29E-02 | 9.54E-01 |  |
|  | 6 months | -0.057 | 0.027 | 3.18E-02 | 0.033 | 0.027 | 2.13E-01 | 0.025 | 0.026 | 3.21E-01 | 1.66E-02 | 8.29E-01 |  |
|  | 8 months | -0.077 | 0.029 | 7.56E-03 | 0.040 | 0.029 | 1.62E-01 | 0.043 | 0.027 | 1.11E-01 | 4.03E-03 | 9.40E-01 |  |
|  | 12 months | -0.039 | 0.029 | 1.70E-01 | 0.050 | 0.029 | 8.59E-02 | 0.043 | 0.027 | 1.13E-01 | 2.88E-02 | 8.71E-01 |  |
|  | 16 months | -0.043 | 0.032 | 1.88E-01 | 0.015 | 0.033 | 6.52E-01 | 0.037 | 0.031 | 2.24E-01 | 2.14E-01 | 6.23E-01 |  |
|  | 2 years | -0.028 | 0.033 | 3.88E-01 | 0.044 | 0.033 | 1.82E-01 | 0.003 | 0.030 | 9.18E-01 | 1.20E-01 | 3.62E-01 |  |
|  | 3 years | -0.014 | 0.032 | 6.67E-01 | 0.044 | 0.033 | 1.82E-01 | 0.011 | 0.030 | 7.04E-01 | 2.09E-01 | 4.65E-01 |  |
|  | 5 years | -0.049 | 0.035 | 1.57E-01 | 0.061 | 0.036 | 8.70E-02 | 0.038 | 0.033 | 2.48E-01 | 2.69E-02 | 6.41E-01 |  |
|  | 7 years | -0.038 | 0.035 | 2.82E-01 | 0.080 | 0.036 | 2.50E-02 | 0.089 | 0.034 | 8.60E-03 | 1.87E-02 | 8.63E-01 |  |
|  | 8 years | -0.023 | 0.041 | 5.68E-01 | 0.047 | 0.040 | 2.45E-01 | 0.060 | 0.038 | 1.11E-01 | 2.22E-01 | 8.06E-01 |  |
| rs3847104 | 6 weeks | 0.012 | 0.023 | 5.94E-01 | -0.031 | 0.023 | 1.78E-01 | 0.009 | 0.022 | 6.97E-01 | 1.85E-01 | 2.16E-01 | Hip circumference |
|  | 3 months | -0.013 | 0.019 | 4.88E-01 | -0.036 | 0.019 | 6.25E-02 | -0.017 | 0.019 | 3.70E-01 | 4.06E-01 | 4.74E-01 |  |
|  | 6 months | 0.003 | 0.019 | 8.71E-01 | -0.031 | 0.019 | 1.09E-01 | -0.005 | 0.019 | 7.74E-01 | 2.10E-01 | 3.40E-01 |  |
|  | 8 months | -0.018 | 0.021 | 3.75E-01 | -0.020 | 0.020 | 3.34E-01 | 0.015 | 0.020 | 4.43E-01 | 9.57E-01 | 2.20E-01 |  |
|  | 12 months | 0.004 | 0.021 | 8.42E-01 | -0.010 | 0.021 | 6.33E-01 | 0.004 | 0.020 | 8.34E-01 | 6.32E-01 | 6.25E-01 |  |
|  | 16 months | 0.004 | 0.023 | 8.78E-01 | 0.006 | 0.023 | 8.09E-01 | 0.024 | 0.023 | 2.93E-01 | 9.50E-01 | 5.76E-01 |  |
|  | 2 years | -0.001 | 0.023 | 9.63E-01 | -0.037 | 0.023 | 1.16E-01 | 0.024 | 0.023 | 2.89E-01 | 2.81E-01 | 6.21E-02 |  |
|  | 3 years | 0.016 | 0.023 | 4.91E-01 | -0.016 | 0.023 | 4.92E-01 | 0.017 | 0.022 | 4.58E-01 | 3.31E-01 | 3.12E-01 |  |
|  | 5 years | 0.016 | 0.025 | 5.32E-01 | 0.005 | 0.025 | 8.49E-01 | 0.032 | 0.025 | 1.88E-01 | 7.57E-01 | 4.33E-01 |  |
|  | 7 years | -0.024 | 0.025 | 3.34E-01 | -0.018 | 0.025 | 4.70E-01 | 0.034 | 0.025 | 1.68E-01 | 8.61E-01 | 1.39E-01 |  |
|  | 8 years | 0.029 | 0.029 | 3.03E-01 | -0.025 | 0.028 | 3.89E-01 | 0.008 | 0.028 | 7.69E-01 | 1.81E-01 | 4.10E-01 |  |

**Supplementary Table 10.**
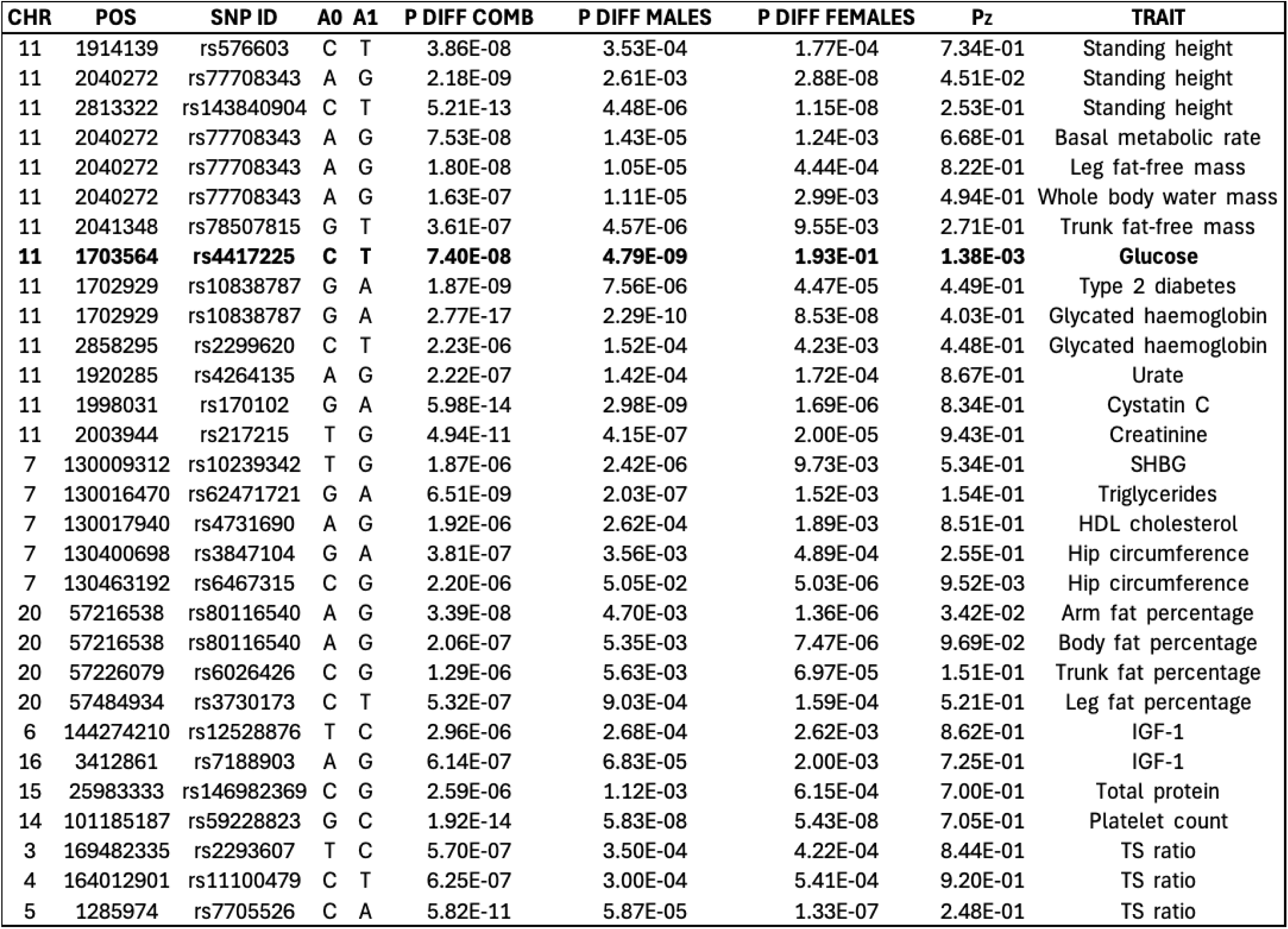
Sex-specificity of Parent-of-Origin effects. CHR: chromosome; POS: genetic position (hg19); SNP ID: variant rs id; A0: reference allele; A1: assessed allele; P

| CHR | POS | SNP ID | A0 | A1 | P DIFF COMB | P DIFF MALES | P DIFF FEMALES | P<sub>Z</sub> | TRAIT |
| --- | --- | --- | --- | --- | --- | --- | --- | --- | --- |
| 11 | 1914139 | rs576603 | C | T | 3.86E-08 | 3.53E-04 | 1.77E-04 | 7.34E-01 | Standing height |
| 11 | 2040272 | rs77708343 | A | G | 2.18E-09 | 2.61E-03 | 2.88E-08 | 4.51E-02 | Standing height |
| 11 | 2813322 | rs143840904 | C | T | 5.21E-13 | 4.48E-06 | 1.15E-08 | 2.53E-01 | Standing height |
| 11 | 2040272 | rs77708343 | A | G | 7.53E-08 | 1.43E-05 | 1.24E-03 | 6.68E-01 | Basal metabolic rate |
| 11 | 2040272 | rs77708343 | A | G | 1.80E-08 | 1.05E-05 | 4.44E-04 | 8.22E-01 | Leg fat-free mass |
| 11 | 2040272 | rs77708343 | A | G | 1.63E-07 | 1.11E-05 | 2.99E-03 | 4.94E-01 | Whole body water mass |
| 11 | 2041348 | rs78507815 | G | T | 3.61E-07 | 4.57E-06 | 9.55E-03 | 2.71E-01 | Trunk fat-free mass |
| **11** | **1703564** | **rs4417225** | **C** | **T** | **7.40E-08** | **4.79E-09** | **1.93E-01** | **1.38E-03** | **Glucose** |
| 11 | 1702929 | rs10838787 | G | A | 1.87E-09 | 7.56E-06 | 4.47E-05 | 4.49E-01 | Type 2 diabetes |
| 11 | 1702929 | rs10838787 | G | A | 2.77E-17 | 2.29E-10 | 8.53E-08 | 4.03E-01 | Glycated haemoglobin |
| 11 | 2858295 | rs2299620 | C | T | 2.23E-06 | 1.52E-04 | 4.23E-03 | 4.48E-01 | Glycated haemoglobin |
| 11 | 1920285 | rs4264135 | A | G | 2.22E-07 | 1.42E-04 | 1.72E-04 | 8.67E-01 | Urate |
| 11 | 1998031 | rs170102 | G | A | 5.98E-14 | 2.98E-09 | 1.69E-06 | 8.34E-01 | Cystatin C |
| 11 | 2003944 | rs217215 | T | G | 4.94E-11 | 4.15E-07 | 2.00E-05 | 9.43E-01 | Creatinine |
| 7 | 130009312 | rs10239342 | T | G | 1.87E-06 | 2.42E-06 | 9.73E-03 | 5.34E-01 | SHBG |
| 7 | 130016470 | rs62471721 | G | A | 6.51E-09 | 2.03E-07 | 1.52E-03 | 1.54E-01 | Triglycerides |
| 7 | 130017940 | rs4731690 | A | G | 1.92E-06 | 2.62E-04 | 1.89E-03 | 8.51E-01 | HDL cholesterol |
| 7 | 130400698 | rs3847104 | G | A | 3.81E-07 | 3.56E-03 | 4.89E-04 | 2.55E-01 | Hip circumference |
| 7 | 130463192 | rs6467315 | C | G | 2.20E-06 | 5.05E-02 | 5.03E-06 | 9.52E-03 | Hip circumference |
| 20 | 57216538 | rs80116540 | A | G | 3.39E-08 | 4.70E-03 | 1.36E-06 | 3.42E-02 | Arm fat percentage |
| 20 | 57216538 | rs80116540 | A | G | 2.06E-07 | 5.35E-03 | 7.47E-06 | 9.69E-02 | Body fat percentage |
| 20 | 57226079 | rs6026426 | C | G | 1.29E-06 | 5.63E-03 | 6.97E-05 | 1.51E-01 | Trunk fat percentage |
| 20 | 57484934 | rs3730173 | C | T | 5.32E-07 | 9.03E-04 | 1.59E-04 | 5.21E-01 | Leg fat percentage |
| 6 | 144274210 | rs12528876 | T | C | 2.96E-06 | 2.68E-04 | 2.62E-03 | 8.62E-01 | IGF-1 |
| 16 | 3412861 | rs7188903 | A | G | 6.14E-07 | 6.83E-05 | 2.00E-03 | 7.25E-01 | IGF-1 |
| 15 | 25983333 | rs146982369 | C | G | 2.59E-06 | 1.12E-03 | 6.15E-04 | 7.00E-01 | Total protein |
| 14 | 101185187 | rs59228823 | G | C | 1.92E-14 | 5.83E-08 | 5.43E-08 | 7.05E-01 | Platelet count |
| 3 | 169482335 | rs2293607 | T | C | 5.70E-07 | 3.50E-04 | 4.22E-04 | 8.44E-01 | TS ratio |
| 4 | 164012901 | rs11100479 | C | T | 6.25E-07 | 3.00E-04 | 5.41E-04 | 9.20E-01 | TS ratio |
| 5 | 1285974 | rs7705526 | C | A | 5.82E-11 | 5.87E-05 | 1.33E-07 | 2.48E-01 | TS ratio |

**Supplementary Table 11.** Replication of parent-of-origin effects in the Estonian Biobank cohort. CHR: chromosome; POS: genetic position (hg38); UKBB SNP: variant exhibiting POE on the trait in the UK Biobank (UKBB); ASSESSED SNP: variant tested for POE in the Estonian Biobank (EstBB); LD: Linkage Disequilibrium (LD) between the UKBB variant and the assessed variant, if not the same variant. LD was computed in the UK Biobank cohort; A0: reference allele of the assessed variant in the EstBB; A1: assessed allele of the assessed variant in the EstBB; N: number of individuals with available trait levels (includes individuals both homozygous and heterozygous for the assessed variant); BETA, SE and P denote effect sizes, standard errors and P-values; PAT, MAT and DIFF denote paternal, maternal and differential tests. TRAIT: phenotype name. Bold indicate replicated associations.

| CHR | POS (hg38) | UKB SNP | ASSESSED SNP | LD | A0 | A1 | N | BETA PAT | SE PAT | P PAT | BETA MAT | SE MAT | P MAT | BETA DIFF | SE DIFF | P DIFF | TRAIT |
| --- | --- | --- | --- | --- | --- | --- | --- | --- | --- | --- | --- | --- | --- | --- | --- | --- | --- |
| 14 | 100718850 | rs59228823 | rs59228823 |  | G | C | 67266 | -0.007 | 0.008 | 3.55E-01 | -0.087 | 0.008 | 4.49E-30 | 0.078 | 0.011 | 2.97E-13 | Platelet count |
| 11 | 1899055 | rs217215 | rs4264135 | 0.613 | A | G | 77270 | -0.028 | 0.006 | 2.21E-06 | 0.030 | 0.006 | 4.27E-07 | -0.058 | 0.008 | 4.40E-12 | Creatinine |
| 11 | 2792092 | rs143840904 | rs143840904 |  | C | T | 61630 | -0.005 | 0.020 | 7.98E-01 | -0.175 | 0.021 | 1.70E-17 | 0.162 | 0.028 | 7.94E-09 | Standing height |
| 7 | 130378099 | rs4731690 | rs4731690 |  | A | G | 81884 | 0.009 | 0.002 | 1.18E-05 | -0.007 | 0.002 | 1.01E-03 | 0.016 | 0.003 | 6.86E-08 | HDL cholesterol |
| 11 | 1921890 | rs576603 | rs735955 | 0.881 | G | A | 61630 | -0.027 | 0.005 | 1.39E-07 | 0.005 | 0.005 | 3.11E-01 | -0.033 | 0.007 | 6.40E-06 | Standing height |
| 11 | 1694102 | rs10838787 | rs7130416 | 0.921 | T | C | 20203 | 0.054 | 0.013 | 2.17E-05 | -0.027 | 0.013 | 3.46E-02 | 0.079 | 0.018 | 9.51E-06 | Glycated haemoglobin |
| 7 | 130376629 | rs62471721 | rs62471721 |  | G | A | 81884 | -0.012 | 0.006 | 5.21E-02 | 0.024 | 0.006 | 4.74E-05 | -0.036 | 0.008 | 1.91E-05 | Triglycerides |
| 11 | 1682334 | rs10838787 | rs4417225 | 0.998 | C | T | 85050 | 0.092 | 0.035 | 9.60E-03 | -0.101 | 0.035 | 4.17E-03 | 0.189 | 0.050 | 1.47E-04 | Type 2 diabetes |
| 11 | 1682334 | rs4417225 | rs4417225 |  | C | T | 80403 | 0.030 | 0.009 | 5.87E-04 | -0.017 | 0.009 | 5.81E-02 | 0.046 | 0.012 | 1.92E-04 | Glucose |
| 11 | 1904723 | rs170102 | rs61868802 | 0.608 | A | G | 21851 | 0.029 | 0.011 | 6.33E-03 | -0.015 | 0.011 | 1.66E-01 | 0.045 | 0.015 | 3.15E-03 | Cystatin C |
| 11 | 2837065 | rs2299620 | rs2299620 |  | C | T | 20203 | 0.060 | 0.034 | 8.07E-02 | -0.070 | 0.034 | 3.83E-02 | 0.130 | 0.048 | 6.79E-03 | Glycated haemoglobin |
| 11 | 1902116 | rs77708343 | rs145861779 | 0.479 | C | T | 61630 | -0.044 | 0.012 | 2.12E-04 | -0.006 | 0.012 | 6.36E-01 | -0.040 | 0.016 | 1.56E-02 | Standing height |
| 7 | 130715870 | rs3847104 | rs3847104 |  | G | A | 55874 | -0.010 | 0.010 | 3.29E-01 | 0.010 | 0.010 | 3.55E-01 | -0.020 | 0.015 | 1.70E-01 | Hip circumference |
| 7 | 130778999 | rs6467315 | rs10954284 | 0.999 | T | A | 55874 | 0.001 | 0.007 | 8.78E-01 | 0.009 | 0.007 | 1.81E-01 | -0.008 | 0.010 | 4.07E-01 | Hip circumference |

## References

[1] T. Moore and D. Haig. “Genomic imprinting in mammalian development: a parental tug-of-war”. In: Trends in genetics: TIG 7.2 (Feb. 1991), pp. 45–49. issn: 0168-9525. doi: 10.1016/0168-9525(91)90230-N.

[2] Juan F Macias-Velasco et al. “Parent-of-origin effects propagate through networks to shape metabolic traits”. In: eLife 11 (), e72989. issn: 2050-084X. doi: 10.7554/eLife.72989. url: https://www.ncbi.nlm.nih.gov/pmc/articles/PMC9075957/ (visited on 08/08/2024).

[3] Robin J. Hofmeister et al. “Parent-of-Origin inference for biobanks”. In: Nature Communications 13.1 (Nov. 5, 2022). Publisher: Nature Publishing Group, p. 6668. issn: 2041-1723. doi: 10.1038/s41467-022-34383-6. url: https://www.nature.com/articles/s41467-022-34383-6 (visited on 08/07/2024).

[4] Hye In Kim et al. “Genome-wide survey of parent-of-origin-specific associations across clinical traits derived from electronic health records”. In: Human Genetics and Genomics Advances 2.3 (June 11, 2021), p. 100039. issn: 2666-2477. doi: 10.1016/j.xhgg.2021.100039. url: https://www.ncbi.nlm.nih.gov/pmc/articles/PMC8756508/ (visited on 08/08/2024).

[5] Augustine Kong et al. “Parental origin of sequence variants associated with complex diseases”. In: Nature 462.7275 (Dec. 2009). Publisher: Nature Publishing Group, pp. 868–874. issn: 1476-4687. doi: 10.1038/nature08625. url: https://www.nature.com/articles/nature08625 (visited on 08/07/2024).

[6] Neil M. Davies et al. “The importance of family-based sampling for biobanks”. In: Nature 634.8035 (Oct. 2024). Publisher: Nature Publishing Group, pp. 795–803. issn: 1476-4687. doi: 10.1038/s41586-024-07721-5. url: https://www.nature.com/articles/s41586-024-07721-5 (visited on 10/25/2024).

[7] Claude Bhérer, Christopher L. Campbell, and Adam Auton. “Refined genetic maps reveal sexual dimorphism in human meiotic recombination at multiple scales”. In: Nature Communications 8.1 (Apr. 25, 2017). Publisher: Nature Publishing Group, p. 14994. issn: 2041-1723. doi: 10.1038/ncomms14994. url: https://www.nature.com/articles/ncomms14994 (visited on 11/22/2024).

[8] Ying Qiao et al. Reconstructing parent genomes using siblings and other relatives. Pages: 2024.05.10.593578 Section: New Results. May 14, 2024. doi: 10.1101/2024.05.10.593578. url: https://www.biorxiv.org/content/10.1101/2024.05.10.593578v1 (visited on 08/08/2024).

[9] Einat Granot-Hershkovitz et al. “Searching for parent-of-origin effects on cardiometabolic traits in imprinted genomic regions”. In: European Journal of Human Genetics 28.5 (May 2020). Publisher: Nature Publishing Group, pp. 646–655. issn: 1476-5438. doi: 10.1038/s41431-019-0568-1. url: https://www.nature.com/articles/s41431-019-0568-1 (visited on 10/25/2024).

[10] Thorhildur Juliusdottir et al. “Distinction between the effects of parental and fetal genomes on fetal growth”. In: Nature Genetics 53.8 (Aug. 2021), pp. 1135–1142. issn: 1546-1718. doi: 10.1038/s41588-021-00896-x.

[11] Robin N. Beaumont et al. “Genome-wide association study of placental weight identifies distinct and shared genetic influences between placental and fetal growth”. In: Nature Genetics 55.11 (Nov. 2023). Publisher: Nature Publishing Group, pp. 1807– 1819. issn: 1546-1718. doi: 10.1038/s41588-023-01520-w. url: https://www.nature.com/articles/s41588-023-01520-w (visited on 10/25/2024).

[12] Magdalena Zoledziewska et al. “Height-reducing variants and selection for short stature in Sardinia”. In: Nature Genetics 47.11 (Nov. 2015). Publisher: Nature Publishing Group, pp. 1352–1356. issn: 1546-1718. doi: 10.1038/ng.3403. url: https://www.nature.com/articles/ng.3403 (visited on 10/25/2024).

[13] Stefania Benonisdottir et al. “Epigenetic and genetic components of height regulation”. In: Nature Communications 7.1 (Nov. 16, 2016). Publisher: Nature Publishing Group, p. 13490. issn: 2041-1723. doi: 10.1038/ncomms13490. url: https://www.nature.com/articles/ncomms13490 (visited on 10/25/2024).

[14] Heather A. Lawson, James M. Cheverud, and Jason B. Wolf. “Genomic imprinting and parent-of-origin effects on complex traits”. In: *Nature reviews*. Genetics 14.9 (Sept. 2013), pp. 609–617. issn: 1471-0056. doi: 10.1038/nrg3543. url: https://www.ncbi.nlm.nih.gov/pmc/articles/PMC3926806/ (visited on 08/08/2024).

[15] Jason B. Wolf et al. “Genome-wide analysis reveals a complex pattern of genomic imprinting in mice”. In: PLoS genetics 4.6 (June 6, 2008), e1000091. issn: 1553-7404. doi: 10.1371/journal.pgen.1000091.

[16] THE GTEX CONSORTIUM. “The GTEx Consortium atlas of genetic regulatory effects across human tissues”. In: Science 369.6509 (Sept. 11, 2020). Publisher: American Association for the Advancement of Science, pp. 1318–1330. doi: 10.1126/science.aaz1776. url: https://www.science.org/doi/10.1126/science.aaz1776 (visited on 10/25/2024).

[17] Xiaoying Wang et al. “Diabetes knowledge predicts HbA1c levels of people with type 2 diabetes mellitus in rural China: a ten-month follow-up study”. In: Scientific Reports 13.1 (Oct. 25, 2023). Publisher: Nature Publishing Group, p. 18248. issn: 2045-2322. doi: 10.1038/s41598-023-45312-y. url: https://www.nature.com/articles/s41598-023-45312-y (visited on 11/18/2024).

[18] Urmo Võsa, et al. “Large-scale cis- and trans-eQTL analyses identify thousands of genetic loci and polygenic scores that regulate blood gene expression”. In: Nature Genetics 53.9 (Sept. 2021). Publisher: Nature Publishing Group, pp. 1300–1310. issn: 1546-1718. doi: 10.1038/s41588-021-00913-z. url: https://www.nature.com/articles/s41588-021-00913-z (visited on 10/25/2024).

[19] Yongtao Guan and Daniel Levy. Abundant Parent-of-origin Effect eQTL in Humans: The Framingham Heart Study. Pages: 2024.06.05.597677 Section: New Results. June 10, 2024. doi: 10.1101/2024.06.05.597677. url: https://www.biorxiv.org/content/10.1101/2024.06.05.597677v1 (visited on 11/15/2024).

[20] Robin J. Hofmeister et al. “Accurate rare variant phasing of whole-genome and whole-exome sequencing data in the UK Biobank”. In: Nature Genetics 55.7 (July 2023). Publisher: Nature Publishing Group, pp. 1243–1249. issn: 1546-1718. doi: 10.1038/s41588-023-01415-w. url: https://www.nature.com/articles/s41588-023-01415-w (visited on 08/08/2024).

[21] Per Magnus et al. “Cohort Profile Update: The Norwegian Mother and Child Cohort Study (MoBa)”. In: International Journal of Epidemiology 45.2 (Apr. 2016), pp. 382–388. issn: 1464-3685. doi: 10.1093/ije/dyw029.

[22] Øyvind Helgeland et al. “Characterization of the genetic architecture of infant and early childhood body mass index”. In: Nature Metabolism 4.3 (Mar. 2022). Publisher: Nature Publishing Group, pp. 344–358. issn: 2522-5812. doi: 10.1038/s42255-022-00549-1. url: https://www.nature.com/articles/s42255-022-00549-1 (visited on 12/03/2024).

[23] Eleonora Porcu et al. “Limited evidence for blood eQTLs in human sexual dimorphism”. In: Genome Medicine 14.1 (Aug. 11, 2022), p. 89. issn: 1756-994X. doi: 10.1186/s13073-022-01088-w. url: https://doi.org/10.1186/s13073-022-01088-w (visited on 11/01/2024).

[24] Elena Bernabeu et al. “Sex differences in genetic architecture in the UK Biobank”. In: Nature Genetics 53.9 (Sept. 2021). Publisher: Nature Publishing Group, pp. 1283– 1289. issn: 1546-1718. doi: 10.1038/s41588-021-00912-0. url: https://www.nature.com/articles/s41588-021-00912-0 (visited on 10/31/2024).

[25] Benjamin B. Sun et al. “Plasma proteomic associations with genetics and health in the UK Biobank”. In: Nature 622.7982 (Oct. 2023). Publisher: Nature Publishing Group, pp. 329–338. issn: 1476-4687. doi: 10.1038/s41586-023-06592-6. url:https://www.nature.com/articles/s41586-023-06592-6 (visited on 10/25/2024).

[26] Lili Milani et al. From Biobanking to Personalized Medicine: the journey of the Estonian Biobank. Pages: 2024.09.22.24313964. Sept. 24, 2024. doi: 10.1101/2024.09.22.24313964. url: https://www.medrxiv.org/content/10.1101/2024.09.22. 24313964v1 (visited on 10/25/2024).

[27] Veryan Codd et al. “Polygenic basis and biomedical consequences of telomere length variation”. In: Nature Genetics 53.10 (Oct. 5, 2021), p. 1425. doi: 10.1038/s41588-021-00944-6. url: https://pmc.ncbi.nlm.nih.gov/articles/PMC8492471/ (visited on 10/31/2024).

[28] Petar N. Grozdanov et al. “SHQ1 is required prior to NAF1 for assembly of H/ACA small nucleolar and telomerase RNPs”. In: RNA 15.6 (June 2009), p. 1188. doi: 10.1261/rna.1532109. url: https://pmc.ncbi.nlm.nih.gov/articles/PMC2685518/ (visited on 10/31/2024).

[29] Andrew S. Venteicher et al. “Identification of ATPases pontin and reptin as telomerase components essential for holoenzyme assembly”. In: Cell 132.6 (Mar. 21, 2008), p. 945. doi: 10.1016/j.cell.2008.01.019. url: https://pmc.ncbi.nlm.nih.gov/articles/PMC2291539/ (visited on 10/31/2024).

[30] Katarina Nordfjäll, et al. “Telomere length and heredity: Indications of paternal inheritance”. In: Proceedings of the National Academy of Sciences 102.45 (Nov. 8, 2005). Publisher: Proceedings of the National Academy of Sciences, pp. 16374–16378. doi: 10.1073/pnas.0501724102. url: https://www.pnas.org/doi/10.1073/pnas. 0501724102 (visited on 10/25/2024).

[31] Qiao Weng et al. “The known genetic loci for telomere length may be involved in the modification of telomeres length after birth”. In: Scientific Reports 6.1 (Dec. 8, 2016). Publisher: Nature Publishing Group, p. 38729. issn: 2045-2322. doi: 10.1038/srep38729. url: https://www.nature.com/articles/srep38729 (visited on 10/25/2024).

[32] Jessica L. Buxton et al. “Human leukocyte telomere length is associated with DNA methylation levels in multiple subtelomeric and imprinted loci”. In: Scientific Reports 4.1 (May 14, 2014). Publisher: Nature Publishing Group, p. 4954. issn: 2045-2322. doi: 10.1038/srep04954. url: https://www.nature.com/articles/srep04954 (visited on 10/25/2024).

[33] M. Kamiya et al. “The cell cycle control gene ZAC/PLAGL1 is imprinted–a strong candidate gene for transient neonatal diabetes”. In: Human Molecular Genetics 9.3 (Feb. 12, 2000), pp. 453–460. issn: 0964-6906. doi: 10.1093/hmg/9.3.453.

[34] Deborah Mackay et al. “Clinical utility gene card for: Transient Neonatal Diabetes Mellitus, 6q24-related”. In: European Journal of Human Genetics 22.9 (Sept. 2014). Publisher: Nature Publishing Group, pp. 1153–1153. issn: 1476-5438. doi: 10.1038/ejhg.2014.27. url: https://www.nature.com/articles/ejhg201427 (visited on 10/30/2024).

[35] Hao Hong et al. “Central IGF1 improves glucose tolerance and insulin sensitivity in mice”. In: Nutrition & Diabetes 7.12 (Dec. 19, 2017). Publisher: Nature Publishing Group, pp. 1–10. issn: 2044-4052. doi: 10.1038/s41387-017-0002-0. url: https://www.nature.com/articles/s41387-017-0002-0 (visited on 11/01/2024).

[36] D. S. Matassa et al. “Oxidative metabolism drives inflammation-induced platinum resistance in human ovarian cancer”. In: Cell Death & Differentiation 23.9 (Sept. 2016). Publisher: Nature Publishing Group, pp. 1542–1554. issn: 1476-5403. doi: 10.1038/cdd.2016.39. url: https://www.nature.com/articles/cdd201639 (visited on 10/25/2024).

[37] Damian Szklarczyk et al. “The STRING database in 2023: protein-protein association networks and functional enrichment analyses for any sequenced genome of interest”. In: Nucleic Acids Research 51 (D1 Jan. 6, 2023), pp. D638–D646. issn: 1362-4962. doi: 10.1093/nar/gkac1000.

[38] Wende Zhu et al. “Meg3-DMR, not the Meg3 gene, regulates imprinting of the Dlk1-Dio3 locus”. In: Developmental Biology 455.1 (Nov. 1, 2019), pp. 10–18. issn: 1095-564X. doi: 10.1016/j.ydbio.2019.07.005.

[39] Tomohiko Kayashima et al. “The novel imprinted carboxypeptidase A4 gene ( CPA4) in the 7q32 imprinting domain”. In: Human Genetics 112.3 (Mar. 2003), pp. 220–226. issn: 0340-6717. doi: 10.1007/s00439-002-0891-3.

[40] Bharati Jadhav et al. “RNA-Seq in 296 phased trios provides a high-resolution map of genomic imprinting”. In: BMC Biology 17.1 (June 24, 2019), p. 50. issn: 1741-7007. doi: 10.1186/s12915-019-0674-0. url: https://doi.org/10.1186/s12915-019-0674-0 (visited on 10/25/2024).

[41] Florian Zink et al. “Insights into imprinting from parent-of-origin phased methylomes and transcriptomes”. In: Nature Genetics 50.11 (Nov. 2018). Publisher: Nature Publishing Group, pp. 1542–1552. issn: 1546-1718. doi: 10.1038/s41588-018-0232-7. url: https://www.nature.com/articles/s41588-018-0232-7 (visited on 10/25/2024).

[42] Nicola Pirastu et al. “Genetic analyses identify widespread sex-differential participation bias”. In: Nature Genetics 53.5 (May 2021). Publisher: Nature Publishing Group, pp. 663–671. issn: 1546-1718. doi: 10.1038/s41588-021-00846-7. url: https://www.nature.com/articles/s41588-021-00846-7 (visited on 11/20/2024).

[43] Mitja I. Kurki et al. “FinnGen provides genetic insights from a well-phenotyped isolated population”. In: Nature 613.7944 (Jan. 2023). Publisher: Nature Publishing Group, pp. 508–518. issn: 1476-4687. doi: 10.1038/s41586-022-05473-8. url: https://www.nature.com/articles/s41586-022-05473-8 (visited on 12/05/2024).

[44] Ben M. Brumpton et al. “The HUNT study: A population-based cohort for genetic research”. In: Cell Genomics 2.10 (Oct. 12, 2022), p. 100193. issn: 2666-979X. doi: 10.1016/j.xgen.2022.100193. url: https://www.sciencedirect.com/science/article/pii/S2666979X22001422 (visited on 12/05/2024).

[45] Clare Bycroft et al. “The UK Biobank resource with deep phenotyping and genomic data”. In: Nature 562.7726 (Oct. 2018). Publisher: Nature Publishing Group, pp. 203–209. issn: 1476-4687. doi: 10.1038/s41586-018-0579-z. url: https://www.nature.com/articles/s41586-018-0579-z (visited on 08/08/2024).

[46] Shaun Purcell et al. “PLINK: A Tool Set for Whole-Genome Association and Population-Based Linkage Analyses”. In: American Journal of Human Genetics 81.3 (Sept. 2007), pp. 559–575. issn: 0002-9297. url: https://www.ncbi.nlm.nih.gov/pmc/articles/PMC1950838/ (visited on 08/08/2024).

[47] Diogo M Ribeiro et al. Phasing report. Google Docs. url: https://docs.google.com/document/d/1EJmh-JcR8HBvu3rjBtREw_50kDIW-sOW2EcC6zweuTc/edit?usp=embed_facebook (visited on 08/08/2024).

[48] Ani Manichaikul et al. “Robust relationship inference in genome-wide association studies”. In: Bioinformatics 26.22 (Nov. 15, 2010), pp. 2867–2873. issn: 1367-4811, 1367-4803. doi: 10.1093/bioinformatics/btq559. url: https://academic.oup.com/bioinformatics/article/26/22/2867/228512 (visited on 08/08/2024).

[49] Stephanie L Battle et al. “A bioinformatics pipeline for estimating mitochondrial DNA copy number and heteroplasmy levels from whole genome sequencing data”. In: NAR Genomics and Bioinformatics 4.2 (June 1, 2022), lqac034. issn: 2631-9268. doi: 10.1093/nargab/lqac034. url: https://doi.org/10.1093/nargab/lqac034 (visited on 08/08/2024).

[50] Simone Rubinacci, Olivier Delaneau, and Jonathan Marchini. “Genotype imputation using the Positional Burrows Wheeler Transform”. In: PLOS Genetics 16.11 (Nov. 16, 2020). Publisher: Public Library of Science, e1009049. issn: 1553-7404. doi: 10.1371/journal.pgen.1009049. url: https://journals.plos.org/plosgenetics/article?id=10.1371/journal.pgen.1009049 (visited on 08/08/2024).

[51] Joelle Mbatchou et al. “Computationally efficient whole-genome regression for quantitative and binary traits”. In: Nature Genetics 53.7 (July 2021). Publisher: Nature Publishing Group, pp. 1097–1103. issn: 1546-1718. doi: 10.1038/s41588-021-00870-7. url: https://www.nature.com/articles/s41588-021-00870-7 (visited on 10/25/2024).

[52] Brendan K. Bulik-Sullivan et al. “LD Score regression distinguishes confounding from polygenicity in genome-wide association studies”. In: Nature Genetics 47.3 (Mar. 2015). Publisher: Nature Publishing Group, pp. 291–295. issn: 1546-1718. doi: 10.1038/ng.3211. url: https://www.nature.com/articles/ng.3211 (visited on 10/28/2024).

[53] Ralf Tambets et al. Genome-wide association study for circulating metabolites in 619,372 individuals. Pages: 2024.10.15.24315557. Oct. 31, 2024. doi: 10.1101/2024.10.15.24315557. url: https://www.medrxiv.org/content/10.1101/2024.10.15.24315557v2 (visited on 11/26/2024).

